# Menopausal symptoms in peri- and postmenopausal women: systematic review and meta-analysis of prevalence, incidence, comorbidities, and clinical outcomes

**DOI:** 10.64898/2026.06.09.26355211

**Authors:** Zhen He, Yao Wang, Zekai Chen, Zhoude Zheng, Jiaqi Chen, Shufan Li, Yueyang Wu, Chris H.L. Thio, Harold Snieder, Qingying Zhang

**Author notes:** **Corresponding authors:** Qingying Zhang, No.22, Xinling Road, Shantou, Guangdong 515041, China, Harold Snieder, address: PO Box 30.001, 9700 RB Groningen, The Netherlands. contribute equally (Zhen He, Yao Wang).

## Abstract

**Introduction:** The global epidemiology of menopausal symptoms among middle-aged and elderly women remains unclear.

**Methods:** Data on prevalence, comorbidities, incidence and outcomes of menopausal symptoms published up until March 1^st^ 2019 were searched in PubMed, Embase and Cochrane databases. We used a random-effects model to compute point estimates of prevalence for 24 types of menopausal symptoms. We narratively summarized the patterns of the comorbidities, incidence and outcomes of menopausal symptoms due to limited data.

**Results:** A total of 239 studies (n≈2.5 million middle-aged and elderly women) from 56 countries and regions were included in the analysis. The global pooled prevalence analysis revealed that hot flashes (48%) and night sweats (30%) were highly prevalent, alongside psychological symptoms like insomnia (47%), irritability (46%), anxiety (39%), and depression (30%). Physical symptoms including joint aches/pain (50%), backache (47%), and tiredness (61%) were also commonly reported. Heat intolerance showed the highest prevalence (76%), while symptoms like urinary incontinence (24%) and poor appetite (8%) were less frequent. These findings highlight the diverse and widespread impact of menopause on women globally, with significant variations across symptom types. Africa showed the highest pooled prevalence across a series of symptoms, compared with other continents. We observed high prevalence in developing countries, especially for psychological and physical symptoms; significant intra-Asian variation in vasomotor symptoms; hypertension and obesity as the most common comorbidities; joint pain, urinary incontinence, and vasomotor symptoms as the most incident complaints; and positive associations with cardiovascular disease in the psychological (depression and insomnia) and physical (joint pain) domains.

**Conclusion:** This study highlights the global burden of menopausal symptoms, with significant differences across continents. The findings call for more inclusive research on underrepresented groups (particularly in Africa) and further investigation into drivers of this marked global heterogeneity in prevalence of menopausal symptoms and their comorbidities, incidence and outcomes.

## Introduction

Middle-aged and older women go through a perimenopausal stage in which their ovarian function gradually declines. It typically starts with the onset of irregular menstruation after 40 years of age and ends one year after menstruation has ceased. Subsequently, postmenopause starts 12 months after the final menstrual period and extends until the end of life^1^. Noteworthy, menopausal symptoms are specific symptoms experienced by both peri- and postmenopausal women.

Menopausal symptoms include hot flashes, night sweats, headaches, palpitations, and depression, among many others^2^. Of these, hot flashes and night sweats (collectively referred to as vasomotor symptoms) are the most characteristic. These symptoms often lead women to seek medical treatment^3^, and together with heightened irritability, can seriously reduce efficiency at work^4^ and quality of life^5^. Numerous studies have reported the prevalence of menopausal symptoms which varies widely in different populations^6-21^. For example, the prevalence of hot flashes among middle-aged women ranges from 17% to 70%, and night sweats range from 15% to 45%, depending on the region. These variations may be attributed to differences in ethnicity, survey methods, study designs, sample representativeness, and other factors. Also, previous studies have reported the incidence^22^ and outcomes^23^ of menopausal symptoms, however, our understanding remains incomplete.

There have been a number of narrative^24-28^ and systematic review^29, 30^ on menopausal symptoms, but so far a study systematically reviewing and meta-analyzing the global data on these symptoms has been lacking. Therefore, in the current study we did a meta-analysis on a comprehensive list of 24 types of menopausal symptoms in peri- and postmenopausal women (including but not limited to hot flashes and night sweats) by using global data. We also examined their associated comorbidities, incidence, and outcomes to comprehensively assess their impact and aimed to provide reference data for medical decision-making.

## Materials and Methods

### Search Strategy

We identified English-language studies with information on prevalence, incidence and outcomes of menopausal symptoms in peri- or postmenopausal women from PubMed, Embase and Cochrane databases. **Supplemental Table S1** shows the detailed search strategy based on the combination of subject and free terms. In brief, the search strategy included the following: (“perimenopause” OR “menopause” OR “postmenopause”) AND (“menopausal syndrome” OR “hot flashes” OR “night sweats” OR “depression” OR “anxiety” OR “arthralgia” OR “dyssomnias”) AND (“stroke” OR “coronary Disease” OR “breast Neoplasms” OR “uterine cervical neoplasms” OR “mortality” OR “prevalence” OR “incidence” OR “ovarian neoplasms” OR “uterine neoplasms” OR “endometrial neoplasms” OR “leiomyoma”). The protocol of this study was registered with the International Platform of Registered Systematic Review and Meta-Analysis Protocols (INPLASY, https://inplasy.com/wp-content/uploads/2020/12/INPLASY-Protocol-1170.pdf) on 12 December 2020 (registration number INPLASY2020120074).

The original protocol title was “Global Epidemiology of Menopausal Syndromes in Peri- and Postmenopausal Women: A Meta-Analytic Assessment of Prevalence, Incidence, and Outcomes.” However, it was revised to the current title based on suggestions from co-authors.

### Study Exclusion Criteria

Exclusion criteria of this meta-analysis were: (1) The study was a review article or abstract or an animal study; (2) the study did not identify prevalence or incidence or outcomes of menopausal symptoms of peri- or postmenopausal women; (3) the study was not written in the English language; (4) the women were younger than 40 years old or premenopausal with regular menstruation^31^; (5) more than 40% of the women were under treatment of hormones in the study; (6) proportion of non-target populations or events (including premenopausal women [<40 years] and/or surgical menopause and/or hormone therapy) exceeded 40%. We chose a cutoff of 40% as the proportion is often higher than 30% in developed countries^14, 32^ and a lot of useful data will be removed if the threshold is too low; (7) studies in which identified prevalence, incidence and/or outcomes of peri-/postmenopausal women were based on women with specific diseases rather than from the general population.

### Data Extraction

Two independent evaluators determined the inclusion of selected papers by reviewing the title and abstract of the selected studies. Six reviewers extracted data and each article was extracted by reading the full text by two reviewers independently. Included studies were cross-sectional and descriptive studies for prevalence of menopausal symptoms and longitudinal studies for incidence and outcomes of menopausal symptoms that were conducted in peri- or postmenopausal women and published before March 1^st^ 2019. Disagreements were resolved by discussion or by a third person. The following information of cross-sectional studies was extracted from each study: Classification of menopausal symptoms, first author, region/country, publication year, study design, random (yes or no, i.e. whether the observational study is designed with random sampling), mean age, status (peri- or postmenopause), data source, diagnostic tool, number of cases with menopausal symptoms, sample size, prevalence, and quality score (A/B/C by Crombie’s items^33^ for cross-sectional studies) (**Supplementary Table S2**). The information extracted regarding prevalence, comorbidities (including hypertension, diabetes, obesity, and hyperlipidemia), as well as the incidence and outcomes of menopausal symptoms, has been summarized in **Supplementary Table S2**.

### Prevalence of menopausal symptoms

The diagnosis of menopausal symptoms was based on questionnaires or scales; however, there were no uniform diagnostic tools. We categorized 24 types of menopausal symptoms according to their names and/or definitions in the literature, and conducted meta-analysis for each symptom separately. These symptoms were categorized into four domains: 1) vasomotor domain (including hot flashes, night sweats/sweating and vasomotor symptoms); 2) psychological domain (including insomnia, depression, anxiety, dizziness, palpitations, irritability, poor concentration, forgetfulness and headache); 3) physical domain (including joint aches/pain, backache, heat intolerance, tiredness, breast tenderness, feelings of suffocation, shortness of breath, numbness of extremities, poor appetite and upset stomach), and; 4) genitourinary domain (including dry/sore vagina and urinary incontinence). Some studies provided prevalence rates of ‘any menopausal symptoms’, and we have also extracted this data. However, a number of symptoms with low reporting frequency, such as change in sexual desire^34^, panic attacks^15^, nausea^35^, eye pain^35^ and so on, extracted from corresponding questionnaires, could not be classified into the aforementioned 24 categories. Therefore, we did not include these unclassifiable symptoms in our meta-analysis (**Supplementary Table S3**). Instead, all infrequently reported symptoms that did not fit into the 24 predefined categories due to their heterogeneity were consolidated under the "Other" classification in **Supplementary Table S3**.

As only a few studies classified menopausal symptoms into mild, moderate, and severe categories with prevalence data^36-39^, we did not extract prevalence for each category separately but calculated overall prevalence by summing individuals with symptoms across all degrees and dividing by the total participants.

Due to the potential influence of factors such as premenopausal status, surgical menopause, and hormone therapy on the prevalence of menopausal symptoms, we refer to these collectively as non-target populations or events. We excluded studies in which >40% of participants were either a) pre-menopausal, b) had iatrogenic menopause (i.e. surgically or medically induced), or c) on hormone replacement therapy. This is done to minimize the impact of non-target populations or events on the pooled results.

### Comorbidity of menopausal symptoms

Previous studies found that menopausal symptoms may be associated with hypertension^40^ and cardiovascular diseases^41^. Therefore, when extracting prevalence data of menopausal symptoms, we concurrently looked at their comorbidities, including hypertension, diabetes, obesity and hyperlipidemia. This represents an exploratory research direction. For instance, if a substantial proportion of women experiencing a particular menopausal symptom also present with hypertension, this may indicate a potential association between the symptom and elevated blood pressure.

### Incidence of menopausal symptoms

We derived the incidence data for menopausal symptoms from longitudinal studies using the formula: incidence (per 1000 person-years) = Number of new cases / [Mean follow-up time in years * Number of participants followed] * 1000. Two cohort studies^42, 43^ reported on a number of menopausal symptoms. However, these studies either did not report incidence directly or lacked data needed for calculation. Hence, we reached out to the corresponding authors via email and verified that our calculated incidence in these two papers was correct. Due to a lack of incidence data, non-target populations or events were retained despite their potential to confound the observed frequency of menopausal symptoms.

### Outcomes of menopausal symptoms

Previous studies reported that menopausal symptoms may be associated with coronary heart disease^44^, mortality^45^, cardiovascular disease^41^, and cancer^46^. Therefore, we predefined a broad spectrum of outcomes of menopausal symptoms, encompassing stroke, coronary disease, leiomyoma, breast, uterine cervical, ovarian, uterine, and endometrial neoplasms, and mortality. This represents an exploratory research direction. Due to lack of data on outcomes based on longitudinal studies, we did not exclude non-target populations or events in this case.

### Overlap in samples

In total, we extracted data from 239 papers, encompassing approximately 2.5 million (n=2,545,501) middle-aged and elderly women. However, several included studies were based on the same or overlapping samples. In these cases, we applied the following two additional inclusion rules:

1. If two studies that were based on an overlapping sample reported the same symptom, the prevalence estimate of this symptom from the study with the larger sample (**Supplementary Table S3**) was taken forward to meta-analysis.
2. If two studies based on an overlapping sample reported the prevalence of two distinct menopausal symptoms, estimates of both symptoms were taken forward to meta-analysis.

Eleven studies^47-57^ were excluded that reported on duplicate samples, but with smaller sample sizes.

### Statistical Analysis

A Venn diagram was used to visualize the included literature (**Figure S1, located in Appendix 1**). Individual study prevalence was calculated by logit transformation and weighted by the inverse of the variance; pooled prevalence was calculated by the restricted maximum-likelihood estimator using a random effects model. The I^2^ statistic was employed to assess the heterogeneity among the studies. Four models for each symptom were analyzed. In the strictest model (model 1), we removed studies with non-target populations or events and non-randomly sampled studies. In model 2, we removed studies with non-target populations or events but not non-randomly sampled studies. In model 3, we removed non-randomly sampled studies but not the studies with non-target populations or events. In the most lenient model (model 4), we did not remove any studies. We then conducted subgroup analysis according to menopausal status, i.e., for the perimenopausal, the postmenopausal and the total group (all women over 40 years). We further plotted the global distribution map of pooled prevalence rates for the 24 types of menopausal symptoms by country, and constructed bar charts to illustrate the pooled prevalence ranking of menopausal symptoms across different countries (**Appendix 1**). A series of line graphs were used to visualize the trend changes for each country over the publication years (**Appendix 2**). Additionally, a forest plot was created to illustrate the distribution of prevalence rates for these symptoms across different continents. Furthermore, we conducted univariable and multivariable meta-regression analyses for model 4 to elucidate the heterogeneity. In these analyses, the logit-transformed proportion of symptoms was used as the dependent variable. Specifically, in the multivariable analysis, we utilized the forced entry method^58^, adjusting for continents (Africa, Asia, Europe, Oceania, South America and North America [reference]), publication years (as a continuous variable), non-target populations or events (no vs. yes), status (perimenopause, postmenopause and peri/postmenopause/>=40yrs [reference]), quality of study (Grade A [reference], B and C), and sample size (as a continuous variable). We performed the Begg-Mazumdar Test^59^ and Egger Test^60^ to assess publication bias in model 4. If publication bias was detected, we used the Trim and Fill method^61^ to adjust the meta-analysis estimate. All analyses were performed using the *meta and metafor* packages in R version 3.6.2. The reporting of the current systematic review adheres to the PRISMA 2020 statement^62^ (**Supplementary Table S4**).

## Results

### Study Characteristics

A total of 239 papers (∼2.5 million middle-aged and elderly women) were included in this meta-analysis, where 216 had prevalence data, 22 had comorbidities data, 10 had incidence data, and 12 had outcomes data (**Figure 1** for flowchart and **Figure S1** for a Venn diagram [See **Appendix 1**]).

**Figure 1.**
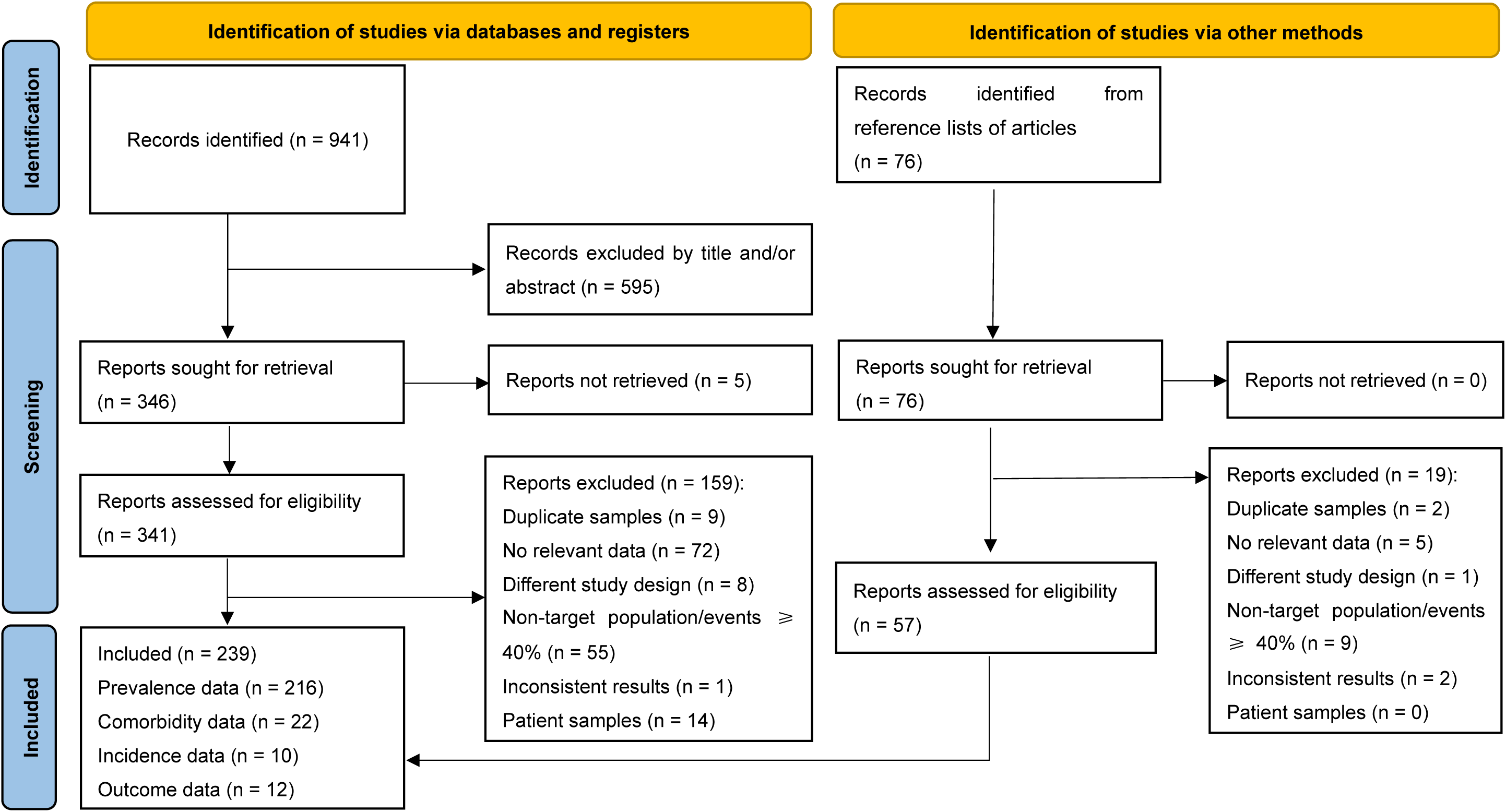
Flowchart of study selection according to the PRISMA guidelines^62^. Non-target population/events include premenopausal women (<40 years) and/or surgical menopause and/or hormone therapy. “Inconsistent results” refer to studies where the reported data and accompanying descriptions appear to lack alignment, making them unsuitable for inclusion in this analysis. “Duplicate samples” refers to studies that reported on duplicate samples, but were excluded because they had smaller sample sizes.

### Pooled prevalence of menopausal symptoms

Figure 2 gives an overview of pooled prevalences of menopausal symptoms globally (n≈2.5 million women) for the perimenopausal, the postmenopausal and the total group (all women over 40 years) for both the strictest model (model 1) and the most lenient model (model 4). A total of 24 types of menopausal symptoms were meta-analyzed. For the most lenient model 4 the pooled prevalence rates [95% CI] in the total group were: hot flashes (0.48 [0.44; 0.52]), night sweats/sweating (0.30 [0.26; 0.34]), vasomotor symptoms (0.51 [0.46; 0.55]), insomnia (0.47 [0.44; 0.49]), depression (0.30 [0.26; 0.34]), anxiety (0.39 [0.33; 0.45]), dizziness (0.35 [0.30; 0.40]), palpitations (0.34 [0.29; 0.39]), irritability (0.46 [0.41; 0.51]), poor concentration (0.44 [0.36; 0.51]), forgetfulness (0.60 [0.54; 0.67]), headache (0.42 [0.38; 0.45]), joint aches/pain (0.50 [0.43; 0.57]), backache (0.47 [0.41; 0.53]), heat intolerance (0.76 [0.70; 0.82]), tiredness (0.61 [0.55; 0.67]), breast tenderness (0.30 [0.19; 0.44]), feelings of suffocation (0.34 [0.28; 0.40]), shortness of breath (0.16 [0.07; 0.33]), numbness of extremities (0.49 [0.44; 0.54]), poor appetite (0.08 [0.05; 0.12]), upset stomach (0.17 [0.10; 0.26]), dry/sore vagina (0.30 [0.26; 0.34]), and urinary incontinence (0.24 [0.20; 0.29]). Prevalence of any menopausal symptom was 0.85 [0.73; 0.92].

**Figure 2.**
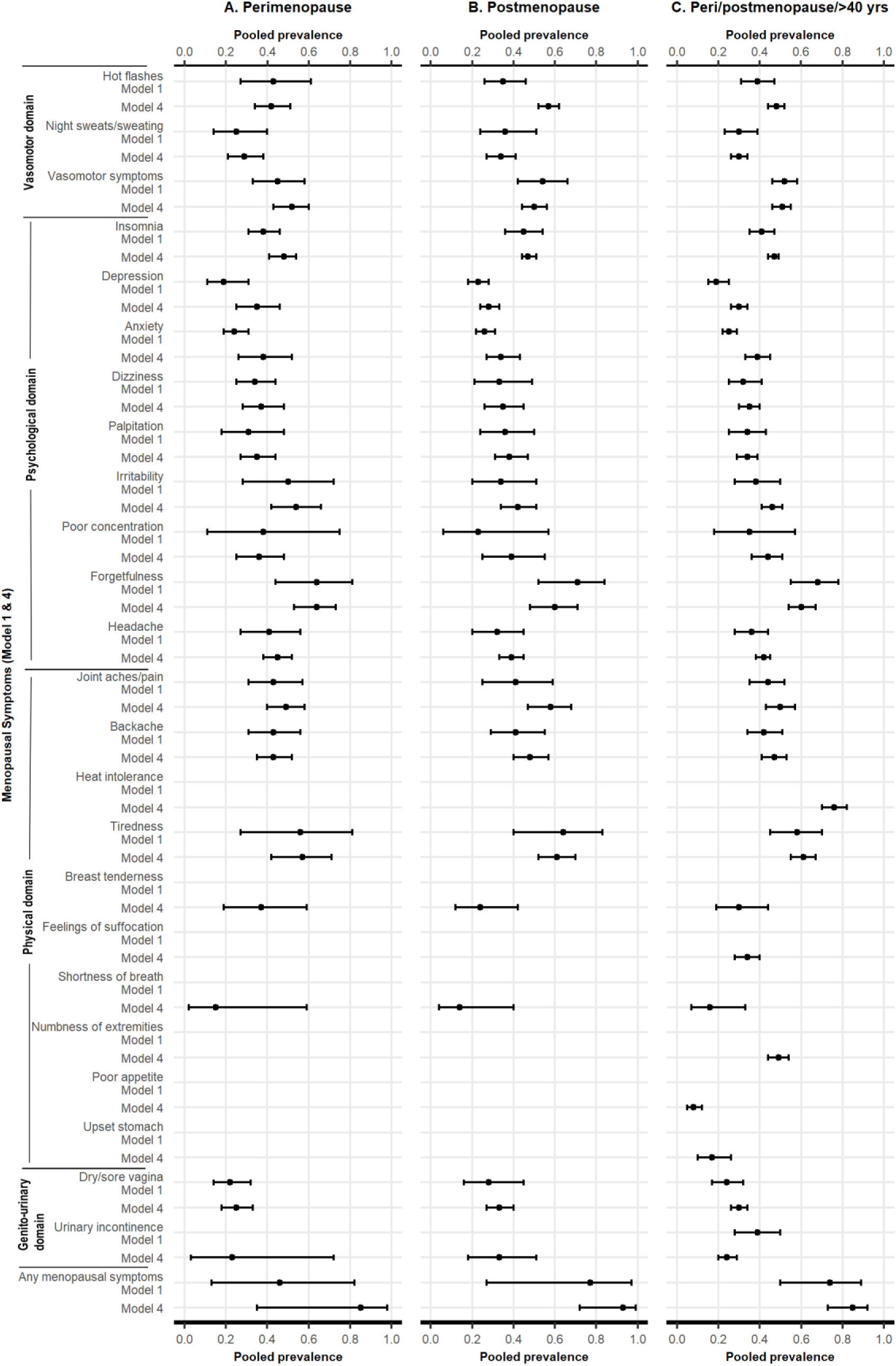
Overview of pooled prevalence of menopausal symptoms globally (n≈2.5 million women) for the perimenopausal, the postmenopausal and the total group (all women over 40 years) for both the strictest model (model 1) and the most lenient model (model 4). Model 1 removed studies with non-target population/events >40% and non-randomly sampled studies, model 4, did not remove any studies; Non-target population/events, included premenopausal women (<40 years) and/or surgical menopause and/or hormone therapy. Vasomotor symptoms included both hot flashes and night sweats, which we could not extract separately.

Women in peri- and post-menopausal stages exhibited comparable pooled prevalence rates between peri- and post-menopausal women across the 24 menopausal symptoms (Figure 2 **and Supplementary Table S5**).

Pooled prevalences, number of studies included, sample sizes and heterogeneity between studies for all models, including intermediate models 2 and 3, in the perimenopausal, the postmenopausal and the total group are shown in **Supplemental Table S5**.

**Global distribution of menopausal symptoms by country and six continents** Figure 3 **and Appendix 3** display the global distribution of prevalence rates of the 24 types of menopausal symptoms across different countries using world maps. In summary, the global distribution of menopausal symptoms shows the following patterns:

1. **High prevalence in developing countries**: Symptoms are more severe in South Asia, Africa, and Latin America, especially psychological and physical symptoms.
2. **Significant variation in Asia**: women from East Asia (e.g., Japan, China) experience mild symptoms, while those from South and Southeast Asia (e.g., Nepal, Thailand) experience more severe symptoms, particularly vasomotor symptoms.
3. **Regional variation in vasomotor symptoms**: Higher prevalences in Europe and Africa, lowest in East Asia.
4. **Data gaps**: There are notable data gaps in South America, the Middle East, and parts of Africa.

**Figure 3.**
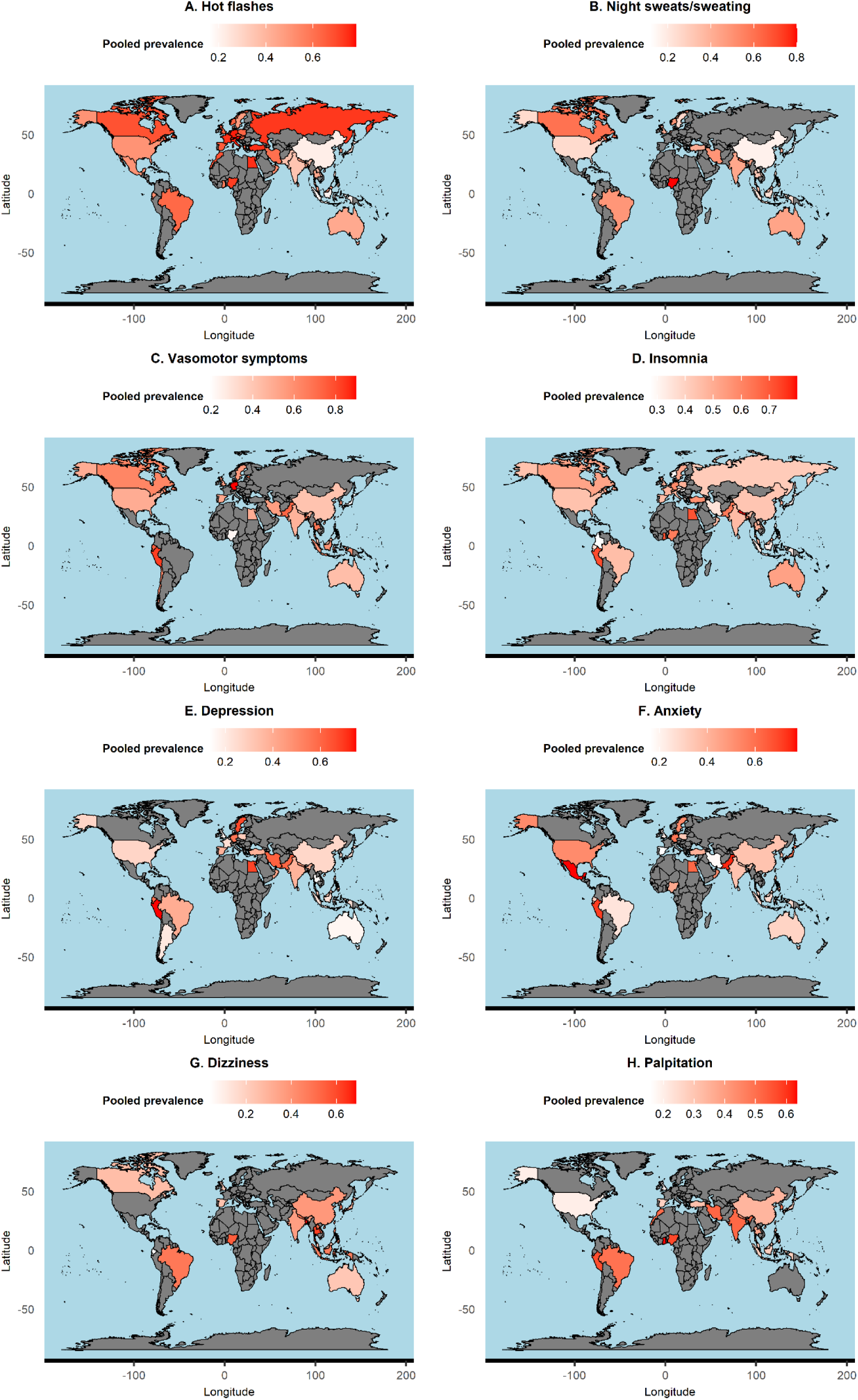
Global distribution of 8 types of menopausal symptoms by countries. The remaining 16 types of symptoms can be found in Appendix 3.

The patterns of each menopausal symptom across countries over publication years were summarized in **Table 1 and Appendix 2**.

**Table 1.**
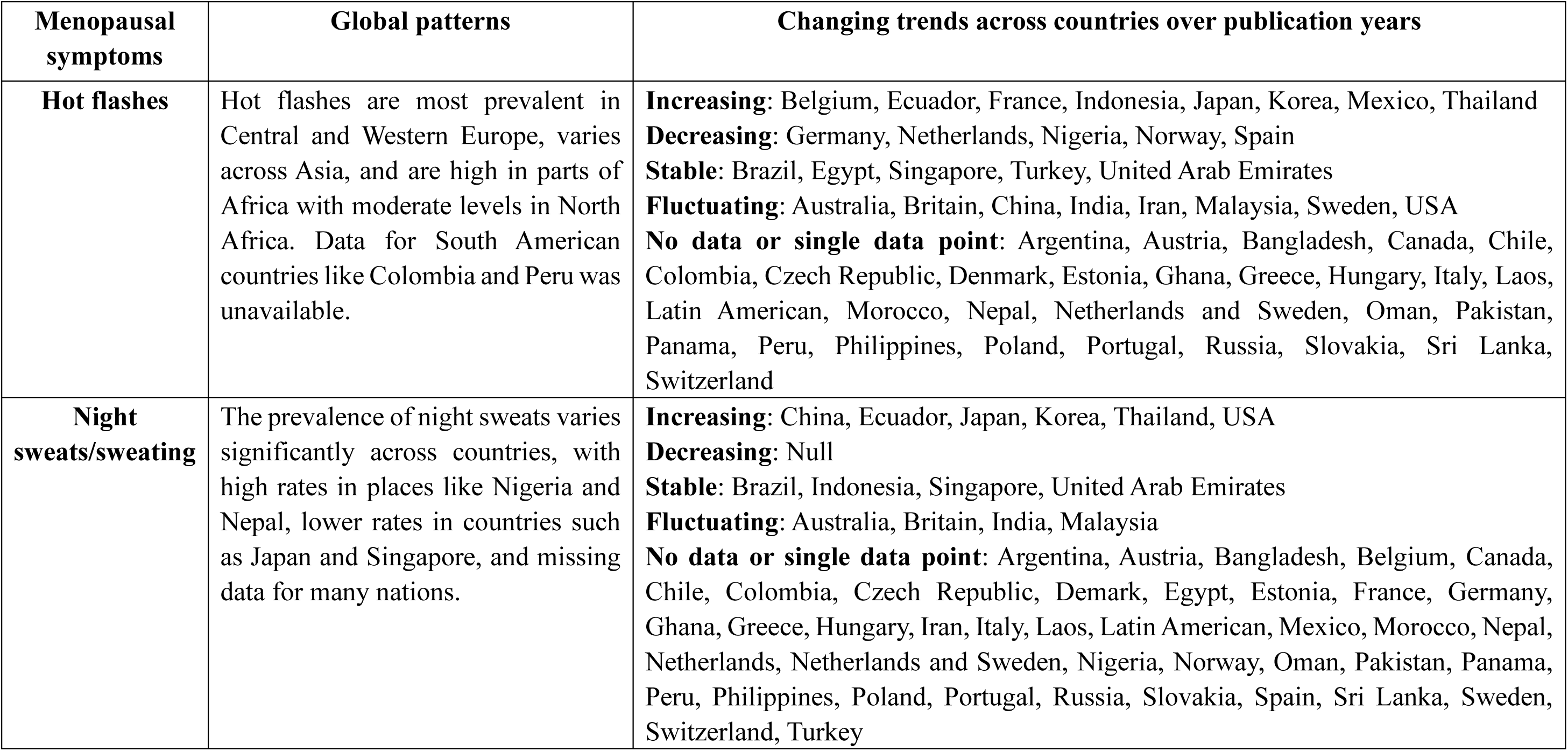
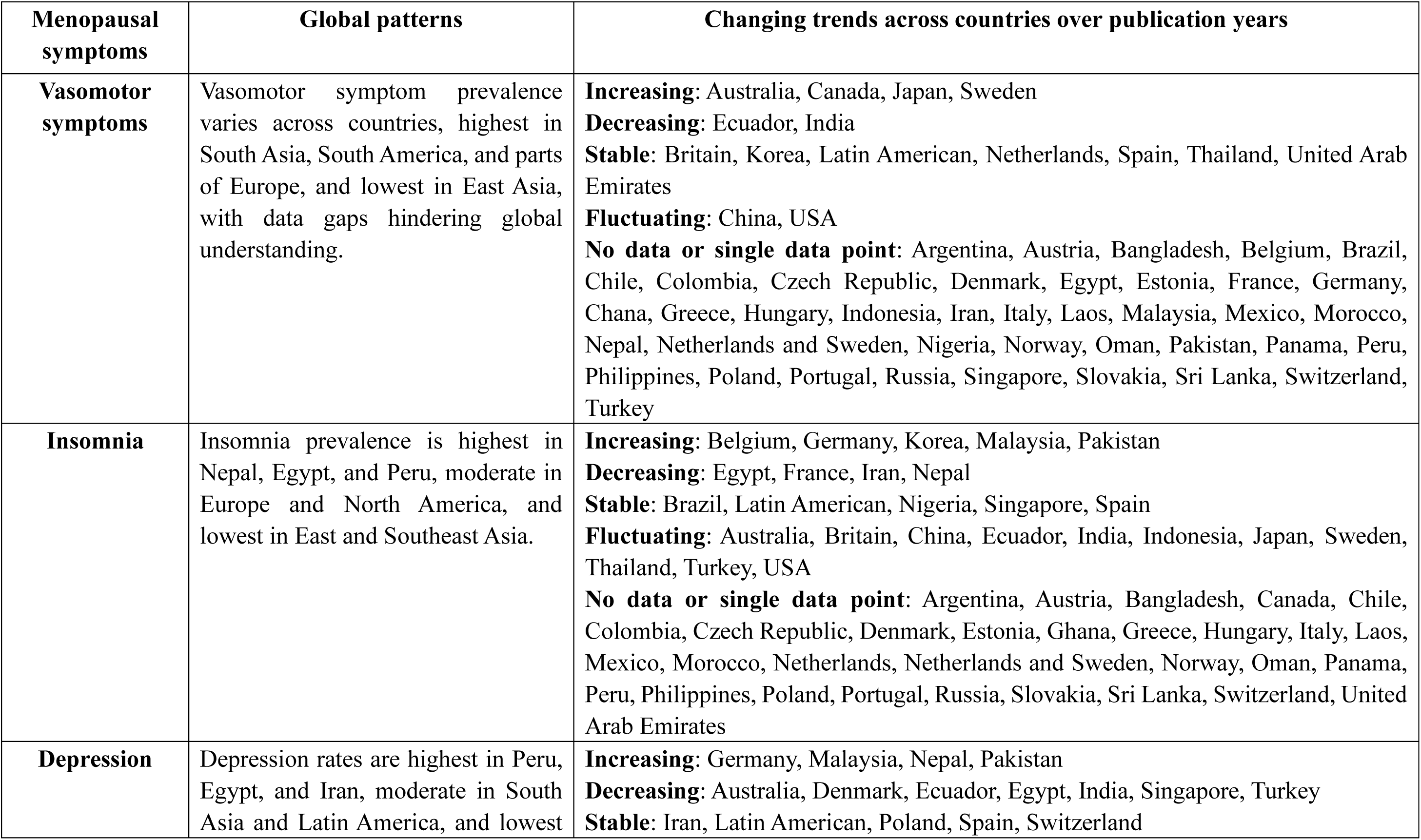
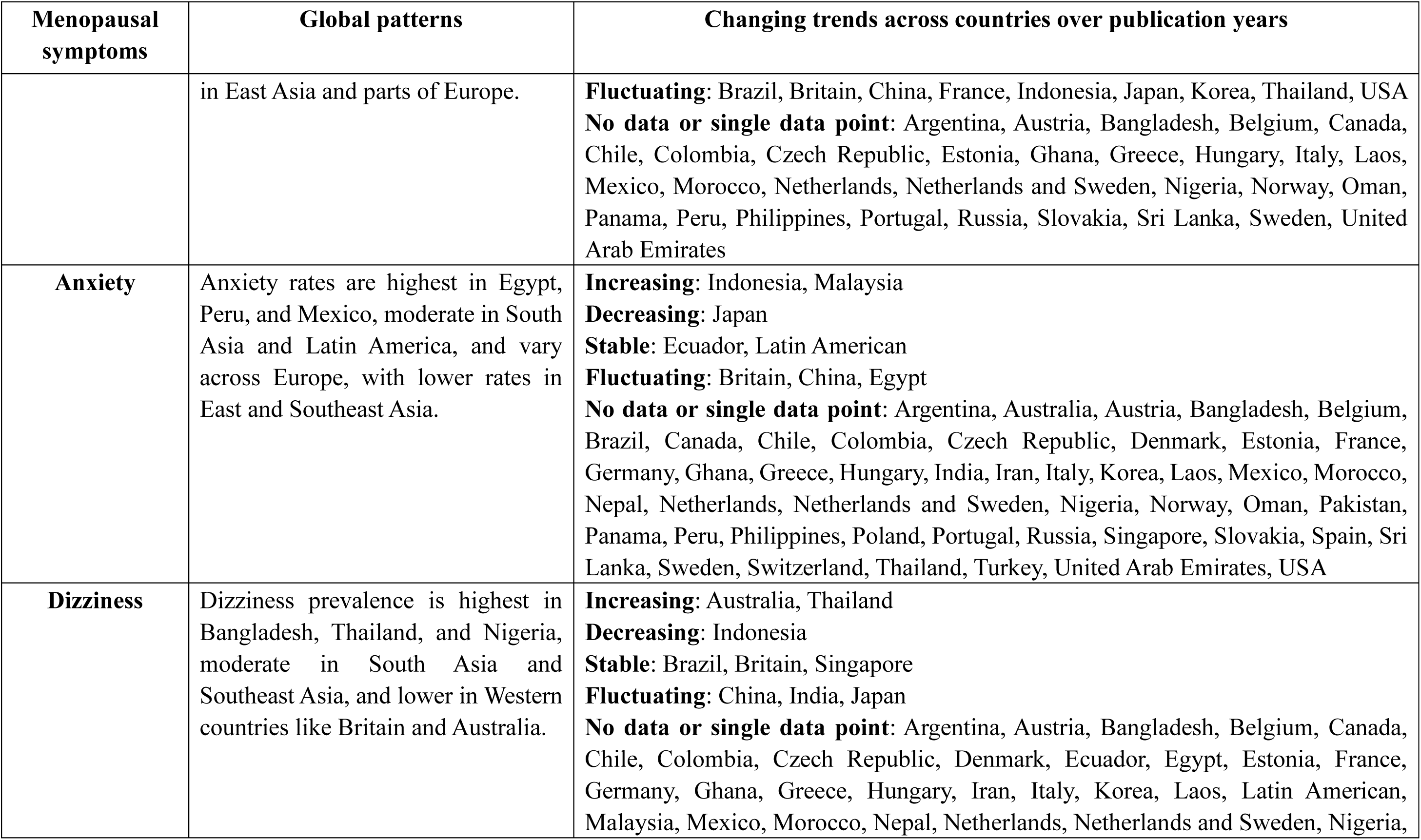
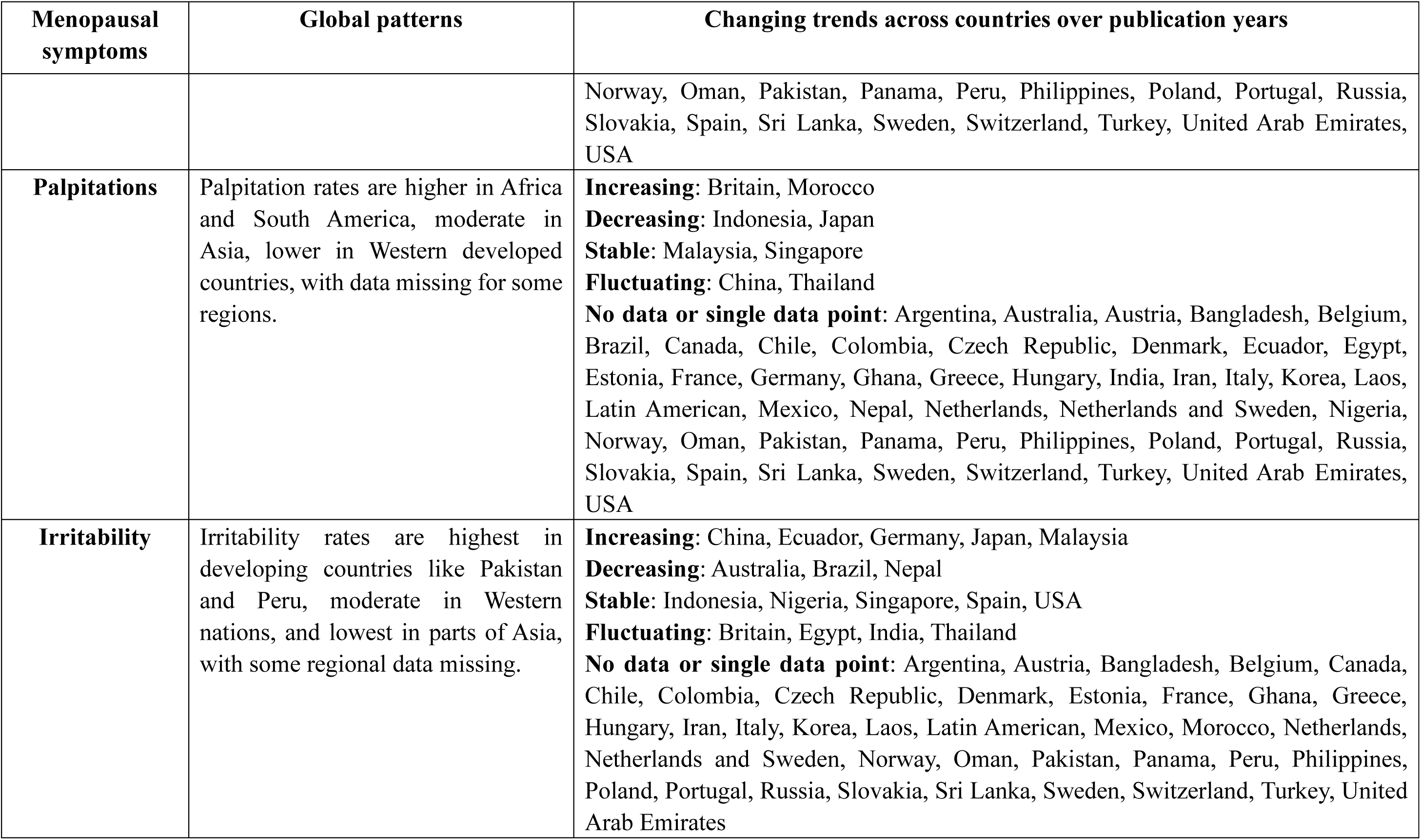
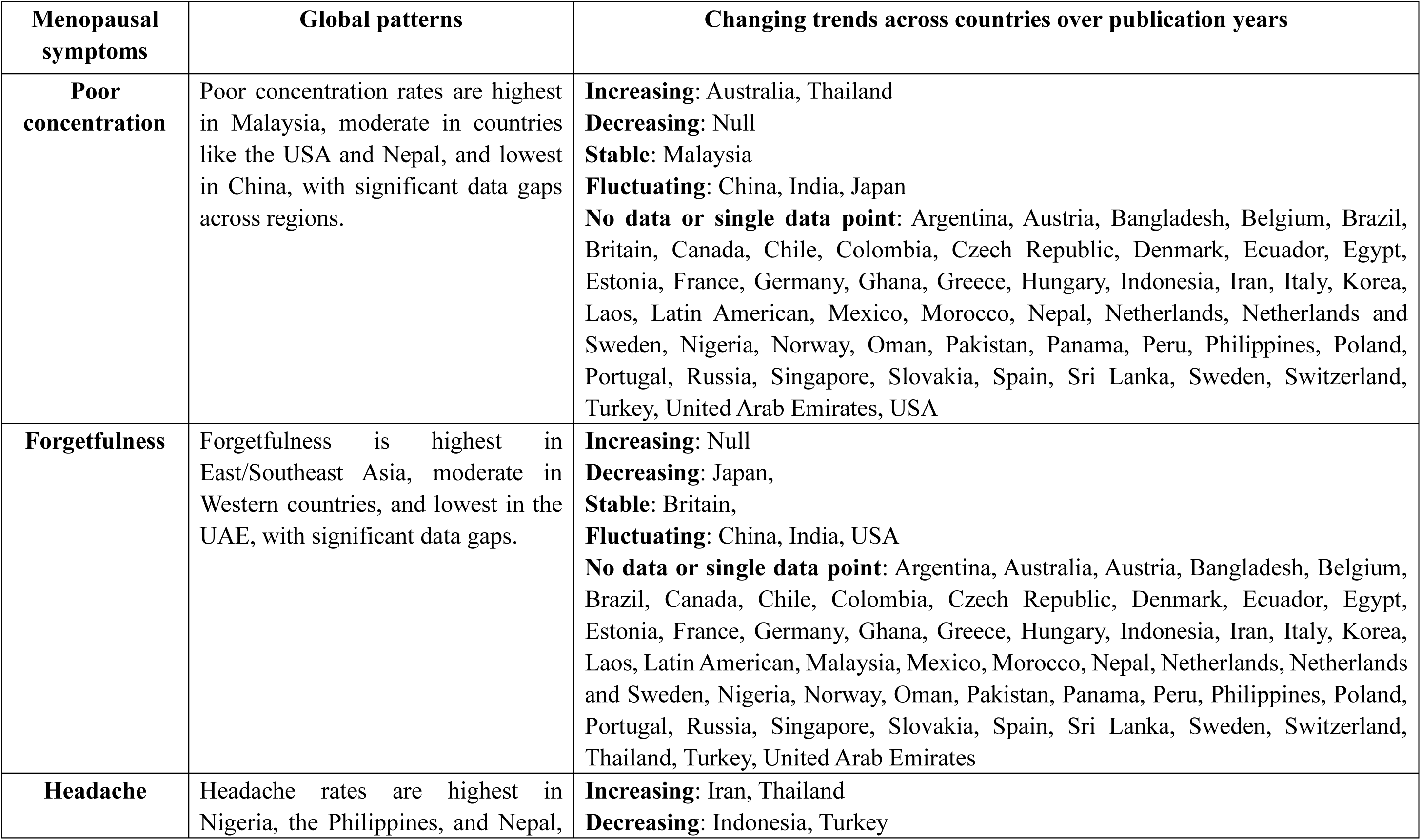
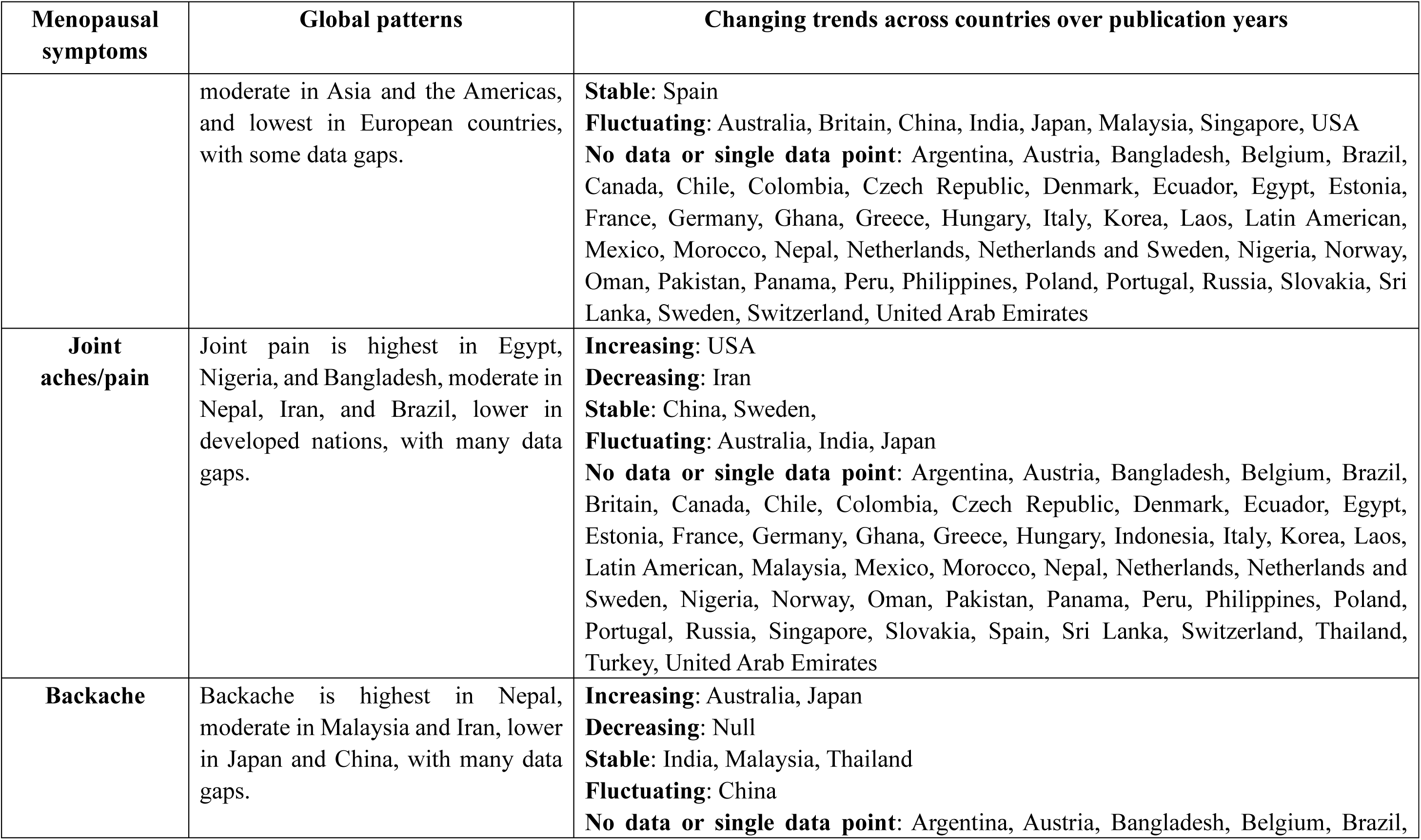
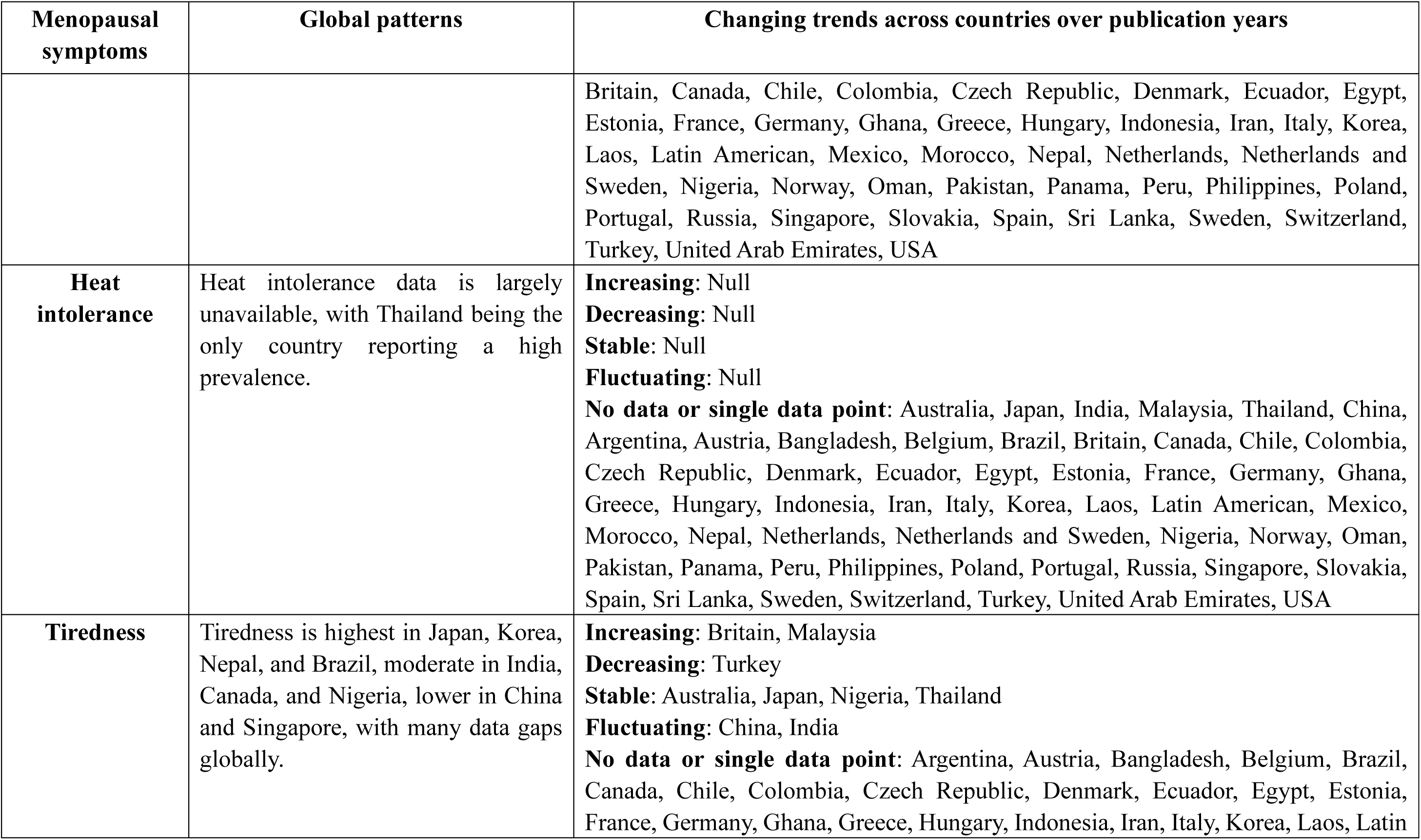
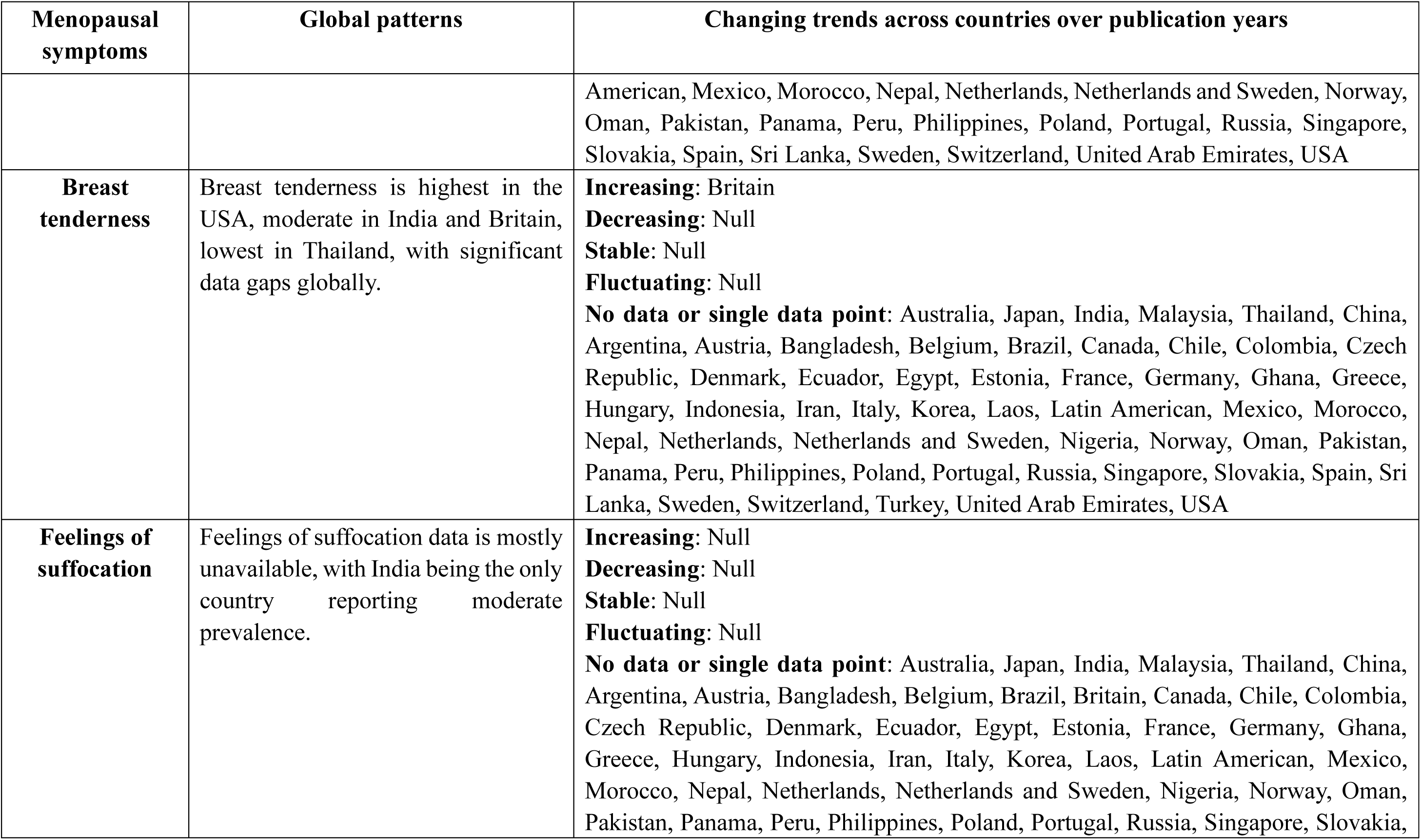
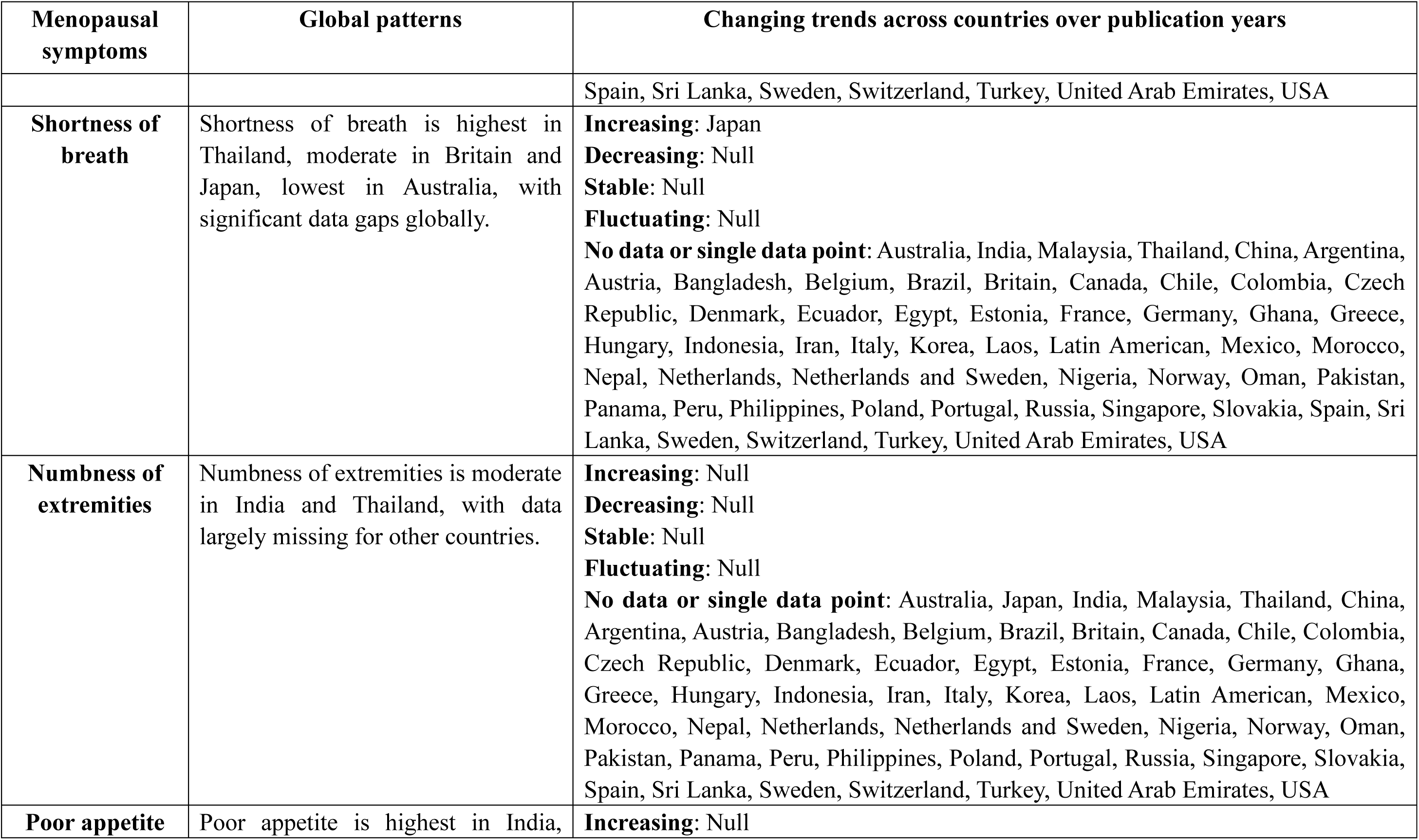
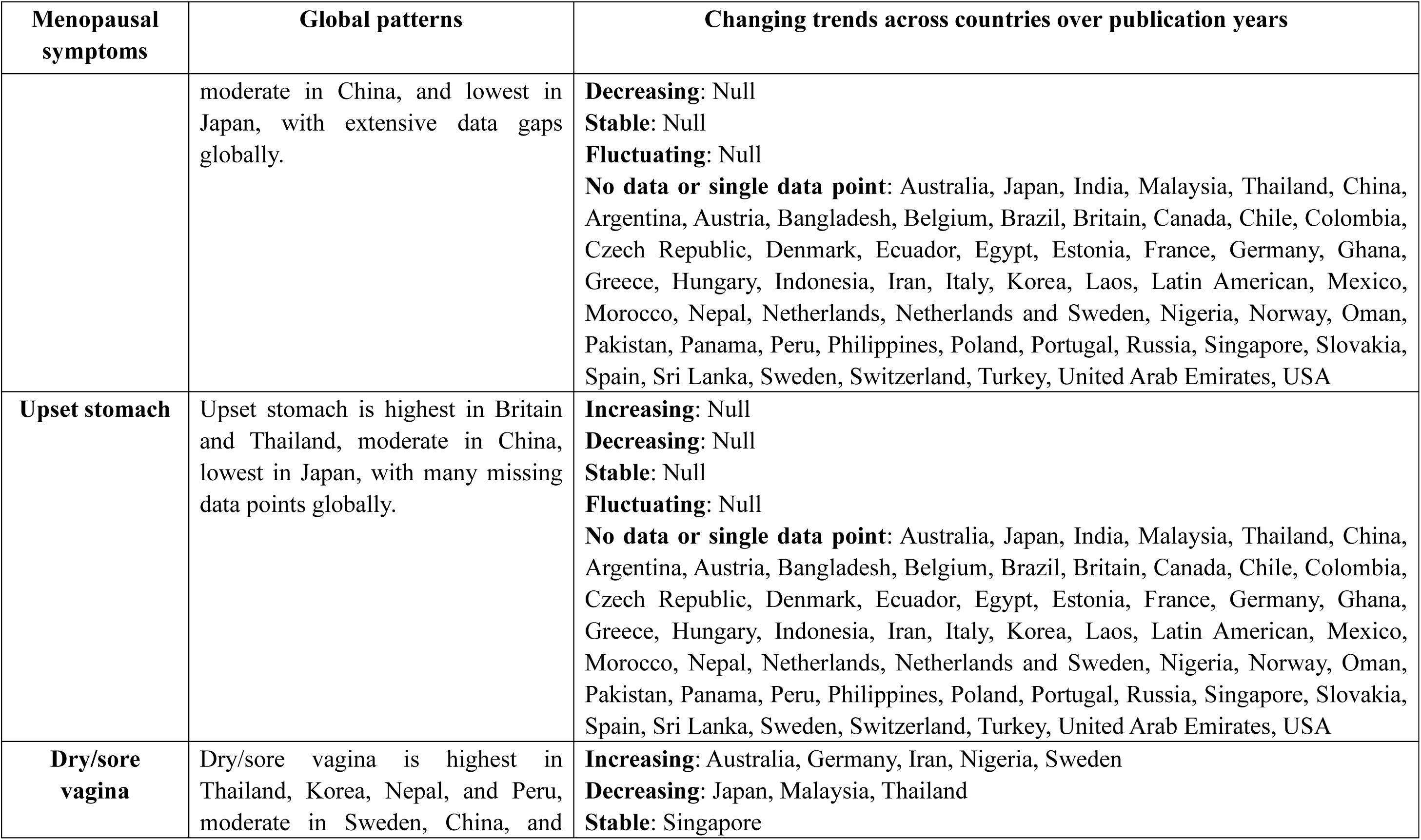
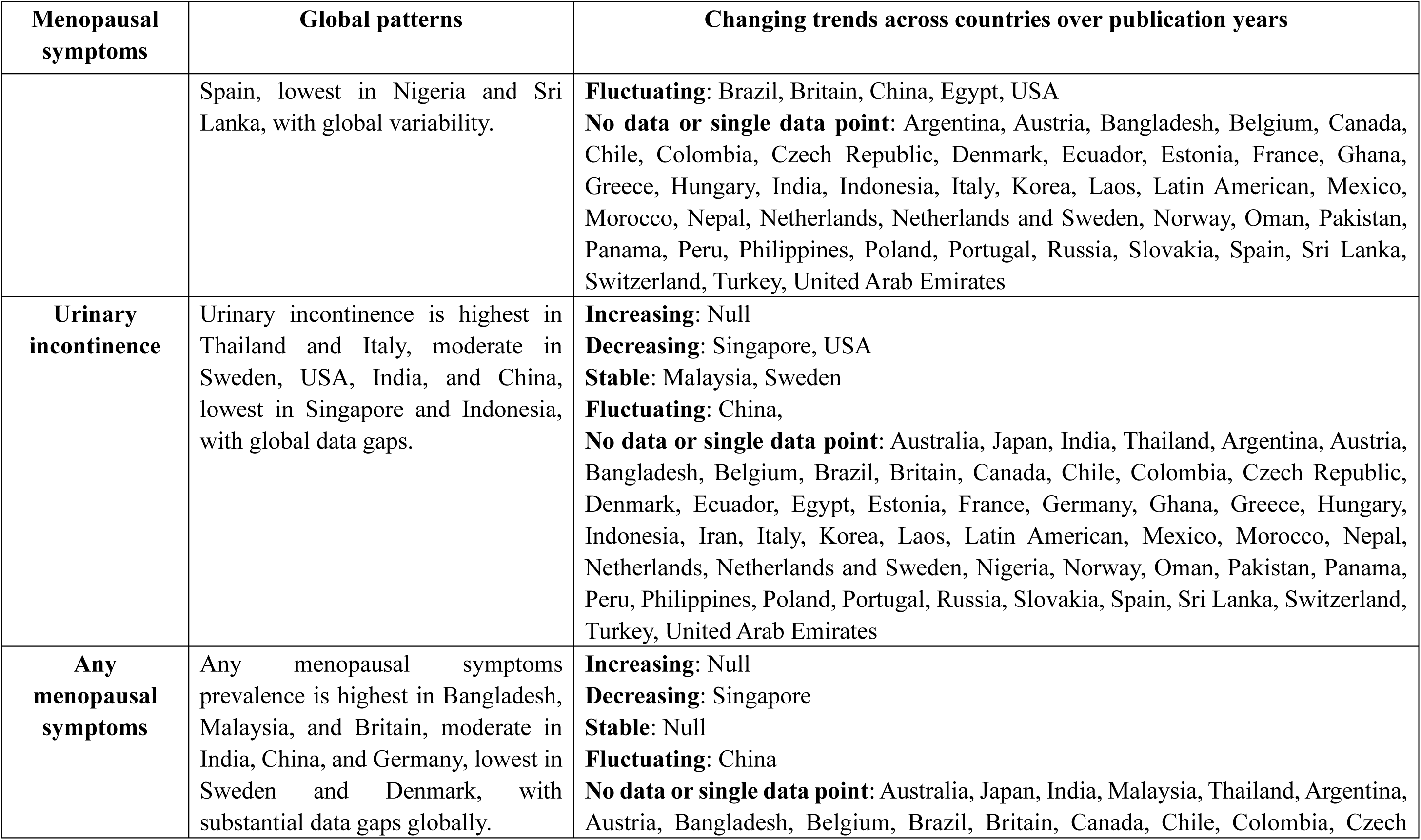
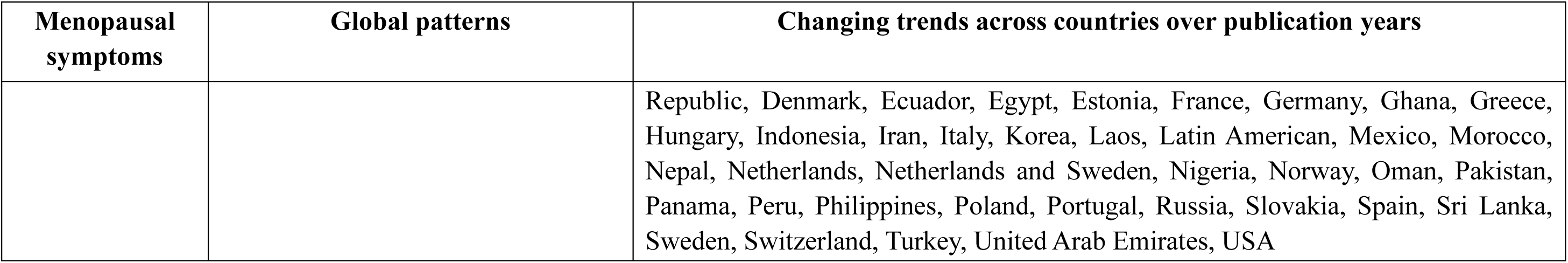
Global patterns and trends in menopausal symptoms among middle-aged and older women across countries and publication years.

| Menopausal symptoms | Global patterns | Changing trends across countries over publication years |
| --- | --- | --- |
| **Hot flashes** | Hot flashes are most prevalent in Central and Western Europe, varies across Asia, and are high in parts of Africa with moderate levels in North Africa. Data for South American countries like Colombia and Peru was unavailable. | **Increasing:** Belgium, Ecuador, France, Indonesia, Japan, Korea, Mexico, Thailand<br>**Decreasing:** Germany, Netherlands, Nigeria, Norway, Spain<br>**Stable:** Brazil, Egypt, Singapore, Turkey, United Arab Emirates<br>**Fluctuating:** Australia, Britain, China, India, Iran, Malaysia, Sweden, USA<br>**No data or single data point:** Argentina, Austria, Bangladesh, Canada, Chile, Colombia, Czech Republic, Denmark, Estonia, Ghana, Greece, Hungary, Italy, Laos, Latin American, Morocco, Nepal, Netherlands and Sweden, Oman, Pakistan, Panama, Peru, Philippines, Poland, Portugal, Russia, Slovakia, Sri Lanka, Switzerland |
| **Night sweats/sweating** | The prevalence of night sweats varies significantly across countries, with high rates in places like Nigeria and Nepal, lower rates in countries such as Japan and Singapore, and missing data for many nations. | **Increasing:** China, Ecuador, Japan, Korea, Thailand, USA<br>**Decreasing:** Null<br>**Stable:** Brazil, Indonesia, Singapore, United Arab Emirates<br>**Fluctuating:** Australia, Britain, India, Malaysia<br>**No data or single data point:** Argentina, Austria, Bangladesh, Belgium, Canada, Chile, Colombia, Czech Republic, Demark, Egypt, Estonia, France, Germany, Ghana, Greece, Hungary, Iran, Italy, Laos, Latin American, Mexico, Morocco, Nepal, Netherlands, Netherlands and Sweden, Nigeria, Norway, Oman, Pakistan, Panama, Peru, Philippines, Poland, Portugal, Russia, Slovakia, Spain, Sri Lanka, Sweden, Switzerland, Turkey |
| **Vasomotor symptoms** | Vasomotor symptom prevalence varies across countries, highest in South Asia, South America, and parts of Europe, and lowest in East Asia, with data gaps hindering global understanding. | **Increasing:** Australia, Canada, Japan, Sweden<br>**Decreasing:** Ecuador, India<br>**Stable:** Britain, Korea, Latin American, Netherlands, Spain, Thailand, United Arab Emirates<br>**Fluctuating:** China, USA<br>**No data or single data point:** Argentina, Austria, Bangladesh, Belgium, Brazil, Chile, Colombia, Czech Republic, Denmark, Egypt, Estonia, France, Germany, Ghana, Greece, Hungary, Indonesia, Iran, Italy, Laos, Malaysia, Mexico, Morocco, Nepal, Netherlands and Sweden, Nigeria, Norway, Oman, Pakistan, Panama, Peru, Philippines, Poland, Portugal, Russia, Singapore, Slovakia, Sri Lanka, Switzerland, Turkey |
| **Insomnia** | Insomnia prevalence is highest in Nepal, Egypt, and Peru, moderate in Europe and North America, and lowest in East and Southeast Asia. | **Increasing:** Belgium, Germany, Korea, Malaysia, Pakistan<br>**Decreasing:** Egypt, France, Iran, Nepal<br>**Stable:** Brazil, Latin American, Nigeria, Singapore, Spain<br>**Fluctuating:** Australia, Britain, China, Ecuador, India, Indonesia, Japan, Sweden, Thailand, Turkey, USA<br>**No data or single data point:** Argentina, Austria, Bangladesh, Canada, Chile, Colombia, Czech Republic, Denmark, Estonia, Ghana, Greece, Hungary, Italy, Laos, Mexico, Morocco, Netherlands, Netherlands and Sweden, Norway, Oman, Panama, Peru, Philippines, Poland, Portugal, Russia, Slovakia, Sri Lanka, Switzerland, United Arab Emirates |
| **Depression** | Depression rates are highest in Peru, Egypt, and Iran, moderate in South Asia and Latin America, and lowest | **Increasing:** Germany, Malaysia, Nepal, Pakistan<br>**Decreasing:** Australia, Denmark, Ecuador, Egypt, India, Singapore, Turkey<br>**Stable:** Iran, Latin American, Poland, Spain, Switzerland |
|  | in East Asia and parts of Europe. | **Fluctuating:** Brazil, Britain, China, France, Indonesia, Japan, Korea, Thailand, USA<br>**No data or single data point:** Argentina, Austria, Bangladesh, Belgium, Canada, Chile, Colombia, Czech Republic, Estonia, Ghana, Greece, Hungary, Italy, Laos, Mexico, Morocco, Netherlands, Netherlands and Sweden, Nigeria, Norway, Oman, Panama, Peru, Philippines, Portugal, Russia, Slovakia, Sri Lanka, Sweden, United Arab Emirates |
| **Anxiety** | Anxiety rates are highest in Egypt, Peru, and Mexico, moderate in South Asia and Latin America, and vary across Europe, with lower rates in East and Southeast Asia. | **Increasing:** Indonesia, Malaysia<br>**Decreasing:** Japan<br>**Stable:** Ecuador, Latin American<br>**Fluctuating:** Britain, China, Egypt<br>**No data or single data point:** Argentina, Australia, Austria, Bangladesh, Belgium, Brazil, Canada, Chile, Colombia, Czech Republic, Denmark, Estonia, France, Germany, Ghana, Greece, Hungary, India, Iran, Italy, Korea, Laos, Mexico, Morocco, Nepal, Netherlands, Netherlands and Sweden, Nigeria, Norway, Oman, Pakistan, Panama, Peru, Philippines, Poland, Portugal, Russia, Singapore, Slovakia, Spain, Sri Lanka, Sweden, Switzerland, Thailand, Turkey, United Arab Emirates, USA |
| **Dizziness** | Dizziness prevalence is highest in Bangladesh, Thailand, and Nigeria, moderate in South Asia and Southeast Asia, and lower in Western countries like Britain and Australia. | **Increasing:** Australia, Thailand<br>**Decreasing:** Indonesia<br>**Stable:** Brazil, Britain, Singapore<br>**Fluctuating:** China, India, Japan<br>**No data or single data point:** Argentina, Austria, Bangladesh, Belgium, Canada, Chile, Colombia, Czech Republic, Denmark, Ecuador, Egypt, Estonia, France, Germany, Ghana, Greece, Hungary, Iran, Italy, Korea, Laos, Latin American, Malaysia, Mexico, Morocco, Nepal, Netherlands, Netherlands and Sweden, Nigeria, |
|  |  | Norway, Oman, Pakistan, Panama, Peru, Philippines, Poland, Portugal, Russia, Slovakia, Spain, Sri Lanka, Sweden, Switzerland, Turkey, United Arab Emirates, USA |
| **Palpitations** | Palpitation rates are higher in Africa and South America, moderate in Asia, lower in Western developed countries, with data missing for some regions. | **Increasing:** Britain, Morocco<br>**Decreasing:** Indonesia, Japan<br>**Stable:** Malaysia, Singapore<br>**Fluctuating:** China, Thailand<br>**No data or single data point:** Argentina, Australia, Austria, Bangladesh, Belgium, Brazil, Canada, Chile, Colombia, Czech Republic, Denmark, Ecuador, Egypt, Estonia, France, Germany, Ghana, Greece, Hungary, India, Iran, Italy, Korea, Laos, Latin American, Mexico, Nepal, Netherlands, Netherlands and Sweden, Nigeria, Norway, Oman, Pakistan, Panama, Peru, Philippines, Poland, Portugal, Russia, Slovakia, Spain, Sri Lanka, Sweden, Switzerland, Turkey, United Arab Emirates, USA |
| **Irritability** | Irritability rates are highest in developing countries like Pakistan and Peru, moderate in Western nations, and lowest in parts of Asia, with some regional data missing. | **Increasing:** China, Ecuador, Germany, Japan, Malaysia<br>**Decreasing:** Australia, Brazil, Nepal<br>**Stable:** Indonesia, Nigeria, Singapore, Spain, USA<br>**Fluctuating:** Britain, Egypt, India, Thailand<br>**No data or single data point:** Argentina, Austria, Bangladesh, Belgium, Canada, Chile, Colombia, Czech Republic, Denmark, Estonia, France, Ghana, Greece, Hungary, Iran, Italy, Korea, Laos, Latin American, Mexico, Morocco, Netherlands, Netherlands and Sweden, Norway, Oman, Pakistan, Panama, Peru, Philippines, Poland, Portugal, Russia, Slovakia, Sri Lanka, Sweden, Switzerland, Turkey, United Arab Emirates |

| **Menopausal symptoms** | **Global patterns** | **Changing trends across countries over publication years** |
| --- | --- | --- |
| **Poor concentration** | Poor concentration rates are highest in Malaysia, moderate in countries like the USA and Nepal, and lowest in China, with significant data gaps across regions. | **Increasing:** Australia, Thailand<br>**Decreasing:** Null<br>**Stable:** Malaysia<br>**Fluctuating:** China, India, Japan<br>**No data or single data point:** Argentina, Austria, Bangladesh, Belgium, Brazil, Britain, Canada, Chile, Colombia, Czech Republic, Denmark, Ecuador, Egypt, Estonia, France, Germany, Ghana, Greece, Hungary, Indonesia, Iran, Italy, Korea, Laos, Latin American, Mexico, Morocco, Nepal, Netherlands, Netherlands and Sweden, Nigeria, Norway, Oman, Pakistan, Panama, Peru, Philippines, Poland, Portugal, Russia, Singapore, Slovakia, Spain, Sri Lanka, Sweden, Switzerland, Turkey, United Arab Emirates, USA |
| **Forgetfulness** | Forgetfulness is highest in East/Southeast Asia, moderate in Western countries, and lowest in the UAE, with significant data gaps. | **Increasing:** Null<br>**Decreasing:** Japan,<br>**Stable:** Britain,<br>**Fluctuating:** China, India, USA<br>**No data or single data point:** Argentina, Australia, Austria, Bangladesh, Belgium, Brazil, Canada, Chile, Colombia, Czech Republic, Denmark, Ecuador, Egypt, Estonia, France, Germany, Ghana, Greece, Hungary, Indonesia, Iran, Italy, Korea, Laos, Latin American, Malaysia, Mexico, Morocco, Nepal, Netherlands, Netherlands and Sweden, Nigeria, Norway, Oman, Pakistan, Panama, Peru, Philippines, Poland, Portugal, Russia, Singapore, Slovakia, Spain, Sri Lanka, Sweden, Switzerland, Thailand, Turkey, United Arab Emirates |
| **Headache** | Headache rates are highest in Nigeria, the Philippines, and Nepal, | **Increasing:** Iran, Thailand<br>**Decreasing:** Indonesia, Turkey |
|  | moderate in Asia and the Americas, and lowest in European countries, with some data gaps. | **Stable:** Spain<br>**Fluctuating:** Australia, Britain, China, India, Japan, Malaysia, Singapore, USA<br>**No data or single data point:** Argentina, Austria, Bangladesh, Belgium, Brazil, Canada, Chile, Colombia, Czech Republic, Denmark, Ecuador, Egypt, Estonia, France, Germany, Ghana, Greece, Hungary, Italy, Korea, Laos, Latin American, Mexico, Morocco, Nepal, Netherlands, Netherlands and Sweden, Nigeria, Norway, Oman, Pakistan, Panama, Peru, Philippines, Poland, Portugal, Russia, Slovakia, Sri Lanka, Sweden, Switzerland, United Arab Emirates |
| **Joint aches/pain** | Joint pain is highest in Egypt, Nigeria, and Bangladesh, moderate in Nepal, Iran, and Brazil, lower in developed nations, with many data gaps. | **Increasing:** USA<br>**Decreasing:** Iran<br>**Stable:** China, Sweden,<br>**Fluctuating:** Australia, India, Japan<br>**No data or single data point:** Argentina, Austria, Bangladesh, Belgium, Brazil, Britain, Canada, Chile, Colombia, Czech Republic, Denmark, Ecuador, Egypt, Estonia, France, Germany, Ghana, Greece, Hungary, Indonesia, Italy, Korea, Laos, Latin American, Malaysia, Mexico, Morocco, Nepal, Netherlands, Netherlands and Sweden, Nigeria, Norway, Oman, Pakistan, Panama, Peru, Philippines, Poland, Portugal, Russia, Singapore, Slovakia, Spain, Sri Lanka, Switzerland, Thailand, Turkey, United Arab Emirates |
| **Backache** | Backache is highest in Nepal, moderate in Malaysia and Iran, lower in Japan and China, with many data gaps. | **Increasing:** Australia, Japan<br>**Decreasing:** Null<br>**Stable:** India, Malaysia, Thailand<br>**Fluctuating:** China<br>**No data or single data point:** Argentina, Austria, Bangladesh, Belgium, Brazil, |
|  |  | Britain, Canada, Chile, Colombia, Czech Republic, Denmark, Ecuador, Egypt, Estonia, France, Germany, Ghana, Greece, Hungary, Indonesia, Iran, Italy, Korea, Laos, Latin American, Mexico, Morocco, Nepal, Netherlands, Netherlands and Sweden, Nigeria, Norway, Oman, Pakistan, Panama, Peru, Philippines, Poland, Portugal, Russia, Singapore, Slovakia, Spain, Sri Lanka, Sweden, Switzerland, Turkey, United Arab Emirates, USA |
| **Heat intolerance** | Heat intolerance data is largely unavailable, with Thailand being the only country reporting a high prevalence. | **Increasing:** Null<br>**Decreasing:** Null<br>**Stable:** Null<br>**Fluctuating:** Null<br>**No data or single data point:** Australia, Japan, India, Malaysia, Thailand, China, Argentina, Austria, Bangladesh, Belgium, Brazil, Britain, Canada, Chile, Colombia, Czech Republic, Denmark, Ecuador, Egypt, Estonia, France, Germany, Ghana, Greece, Hungary, Indonesia, Iran, Italy, Korea, Laos, Latin American, Mexico, Morocco, Nepal, Netherlands, Netherlands and Sweden, Nigeria, Norway, Oman, Pakistan, Panama, Peru, Philippines, Poland, Portugal, Russia, Singapore, Slovakia, Spain, Sri Lanka, Sweden, Switzerland, Turkey, United Arab Emirates, USA |
| **Tiredness** | Tiredness is highest in Japan, Korea, Nepal, and Brazil, moderate in India, Canada, and Nigeria, lower in China and Singapore, with many data gaps globally. | **Increasing:** Britain, Malaysia<br>**Decreasing:** Turkey<br>**Stable:** Australia, Japan, Nigeria, Thailand<br>**Fluctuating:** China, India<br>**No data or single data point:** Argentina, Austria, Bangladesh, Belgium, Brazil, Canada, Chile, Colombia, Czech Republic, Denmark, Ecuador, Egypt, Estonia, France, Germany, Ghana, Greece, Hungary, Indonesia, Iran, Italy, Korea, Laos, Latin |
|  |  | American, Mexico, Morocco, Nepal, Netherlands, Netherlands and Sweden, Norway, Oman, Pakistan, Panama, Peru, Philippines, Poland, Portugal, Russia, Singapore, Slovakia, Spain, Sri Lanka, Sweden, Switzerland, United Arab Emirates, USA |
| **Breast tenderness** | Breast tenderness is highest in the USA, moderate in India and Britain, lowest in Thailand, with significant data gaps globally. | **Increasing:** Britain<br>**Decreasing:** Null<br>**Stable:** Null<br>**Fluctuating:** Null<br>**No data or single data point:** Australia, Japan, India, Malaysia, Thailand, China, Argentina, Austria, Bangladesh, Belgium, Brazil, Canada, Chile, Colombia, Czech Republic, Denmark, Ecuador, Egypt, Estonia, France, Germany, Ghana, Greece, Hungary, Indonesia, Iran, Italy, Korea, Laos, Latin American, Mexico, Morocco, Nepal, Netherlands, Netherlands and Sweden, Nigeria, Norway, Oman, Pakistan, Panama, Peru, Philippines, Poland, Portugal, Russia, Singapore, Slovakia, Spain, Sri Lanka, Sweden, Switzerland, Turkey, United Arab Emirates, USA |
| **Feelings of suffocation** | Feelings of suffocation data is mostly unavailable, with India being the only country reporting moderate prevalence. | **Increasing:** Null<br>**Decreasing:** Null<br>**Stable:** Null<br>**Fluctuating:** Null<br>**No data or single data point:** Australia, Japan, India, Malaysia, Thailand, China, Argentina, Austria, Bangladesh, Belgium, Brazil, Britain, Canada, Chile, Colombia, Czech Republic, Denmark, Ecuador, Egypt, Estonia, France, Germany, Ghana, Greece, Hungary, Indonesia, Iran, Italy, Korea, Laos, Latin American, Mexico, Morocco, Nepal, Netherlands, Netherlands and Sweden, Nigeria, Norway, Oman, Pakistan, Panama, Peru, Philippines, Poland, Portugal, Russia, Singapore, Slovakia, |
|  |  | Spain, Sri Lanka, Sweden, Switzerland, Turkey, United Arab Emirates, USA |
| **Shortness of breath** | Shortness of breath is highest in Thailand, moderate in Britain and Japan, lowest in Australia, with significant data gaps globally. | **Increasing:** Japan<br>**Decreasing:** Null<br>**Stable:** Null<br>**Fluctuating:** Null<br>**No data or single data point:** Australia, India, Malaysia, Thailand, China, Argentina, Austria, Bangladesh, Belgium, Brazil, Britain, Canada, Chile, Colombia, Czech Republic, Denmark, Ecuador, Egypt, Estonia, France, Germany, Ghana, Greece, Hungary, Indonesia, Iran, Italy, Korea, Laos, Latin American, Mexico, Morocco, Nepal, Netherlands, Netherlands and Sweden, Nigeria, Norway, Oman, Pakistan, Panama, Peru, Philippines, Poland, Portugal, Russia, Singapore, Slovakia, Spain, Sri Lanka, Sweden, Switzerland, Turkey, United Arab Emirates, USA |
| **Numbness of extremities** | Numbness of extremities is moderate in India and Thailand, with data largely missing for other countries. | **Increasing:** Null<br>**Decreasing:** Null<br>**Stable:** Null<br>**Fluctuating:** Null<br>**No data or single data point:** Australia, Japan, India, Malaysia, Thailand, China, Argentina, Austria, Bangladesh, Belgium, Brazil, Britain, Canada, Chile, Colombia, Czech Republic, Denmark, Ecuador, Egypt, Estonia, France, Germany, Ghana, Greece, Hungary, Indonesia, Iran, Italy, Korea, Laos, Latin American, Mexico, Morocco, Nepal, Netherlands, Netherlands and Sweden, Nigeria, Norway, Oman, Pakistan, Panama, Peru, Philippines, Poland, Portugal, Russia, Singapore, Slovakia, Spain, Sri Lanka, Sweden, Switzerland, Turkey, United Arab Emirates, USA |
| **Poor appetite** | Poor appetite is highest in India, | **Increasing:** Null |

| **Menopausal symptoms** | **Global patterns** | **Changing trends across countries over publication years** |
| --- | --- | --- |
|  | moderate in China, and lowest in Japan, with extensive data gaps globally. | **Decreasing:** Null<br>**Stable:** Null<br>**Fluctuating:** Null<br>**No data or single data point:** Australia, Japan, India, Malaysia, Thailand, China, Argentina, Austria, Bangladesh, Belgium, Brazil, Britain, Canada, Chile, Colombia, Czech Republic, Denmark, Ecuador, Egypt, Estonia, France, Germany, Ghana, Greece, Hungary, Indonesia, Iran, Italy, Korea, Laos, Latin American, Mexico, Morocco, Nepal, Netherlands, Netherlands and Sweden, Nigeria, Norway, Oman, Pakistan, Panama, Peru, Philippines, Poland, Portugal, Russia, Singapore, Slovakia, Spain, Sri Lanka, Sweden, Switzerland, Turkey, United Arab Emirates, USA |
| **Upset stomach** | Upset stomach is highest in Britain and Thailand, moderate in China, lowest in Japan, with many missing data points globally. | **Increasing:** Null<br>**Decreasing:** Null<br>**Stable:** Null<br>**Fluctuating:** Null<br>**No data or single data point:** Australia, Japan, India, Malaysia, Thailand, China, Argentina, Austria, Bangladesh, Belgium, Brazil, Britain, Canada, Chile, Colombia, Czech Republic, Denmark, Ecuador, Egypt, Estonia, France, Germany, Ghana, Greece, Hungary, Indonesia, Iran, Italy, Korea, Laos, Latin American, Mexico, Morocco, Nepal, Netherlands, Netherlands and Sweden, Nigeria, Norway, Oman, Pakistan, Panama, Peru, Philippines, Poland, Portugal, Russia, Singapore, Slovakia, Spain, Sri Lanka, Sweden, Switzerland, Turkey, United Arab Emirates, USA |
| **Dry/sore vagina** | Dry/sore vagina is highest in Thailand, Korea, Nepal, and Peru, moderate in Sweden, China, and | **Increasing:** Australia, Germany, Iran, Nigeria, Sweden<br>**Decreasing:** Japan, Malaysia, Thailand<br>**Stable:** Singapore |
|  | Spain, lowest in Nigeria and Sri Lanka, with global variability. | **Fluctuating:** Brazil, Britain, China, Egypt, USA<br>**No data or single data point:** Argentina, Austria, Bangladesh, Belgium, Canada, Chile, Colombia, Czech Republic, Denmark, Ecuador, Estonia, France, Ghana, Greece, Hungary, India, Indonesia, Italy, Korea, Laos, Latin American, Mexico, Morocco, Nepal, Netherlands, Netherlands and Sweden, Norway, Oman, Pakistan, Panama, Peru, Philippines, Poland, Portugal, Russia, Slovakia, Spain, Sri Lanka, Switzerland, Turkey, United Arab Emirates |
| **Urinary incontinence** | Urinary incontinence is highest in Thailand and Italy, moderate in Sweden, USA, India, and China, lowest in Singapore and Indonesia, with global data gaps. | **Increasing:** Null<br>**Decreasing:** Singapore, USA<br>**Stable:** Malaysia, Sweden<br>**Fluctuating:** China,<br>**No data or single data point:** Australia, Japan, India, Thailand, Argentina, Austria, Bangladesh, Belgium, Brazil, Britain, Canada, Chile, Colombia, Czech Republic, Denmark, Ecuador, Egypt, Estonia, France, Germany, Ghana, Greece, Hungary, Indonesia, Iran, Italy, Korea, Laos, Latin American, Mexico, Morocco, Nepal, Netherlands, Netherlands and Sweden, Nigeria, Norway, Oman, Pakistan, Panama, Peru, Philippines, Poland, Portugal, Russia, Slovakia, Spain, Sri Lanka, Switzerland, Turkey, United Arab Emirates |
| **Any menopausal symptoms** | Any menopausal symptoms prevalence is highest in Bangladesh, Malaysia, and Britain, moderate in India, China, and Germany, lowest in Sweden and Denmark, with substantial data gaps globally. | **Increasing:** Null<br>**Decreasing:** Singapore<br>**Stable:** Null<br>**Fluctuating:** China<br>**No data or single data point:** Australia, Japan, India, Malaysia, Thailand, Argentina, Austria, Bangladesh, Belgium, Brazil, Britain, Canada, Chile, Colombia, Czech |
|  |  | Republic, Denmark, Ecuador, Egypt, Estonia, France, Germany, Ghana, Greece, Hungary, Indonesia, Iran, Italy, Korea, Laos, Latin American, Mexico, Morocco, Nepal, Netherlands, Netherlands and Sweden, Nigeria, Norway, Oman, Pakistan, Panama, Peru, Philippines, Poland, Portugal, Russia, Slovakia, Spain, Sri Lanka, Sweden, Switzerland, Turkey, United Arab Emirates, USA |

For a more detailed analysis of the pooled prevalence ranking of other menopausal symptoms in different countries, please refer to **Figure A-Y** in **Appendix 1**. In addition, please see Figure 4 for further details regarding the pooled prevalence of these symptoms across continents. Africa, albeit with limited data, demonstrated the highest pooled prevalence of hot flashes, night sweats, insomnia, depression, dizziness, palpitations, irritability, headache, joint pain, and any menopausal symptoms, compared with other continents.

**Figure 4.**
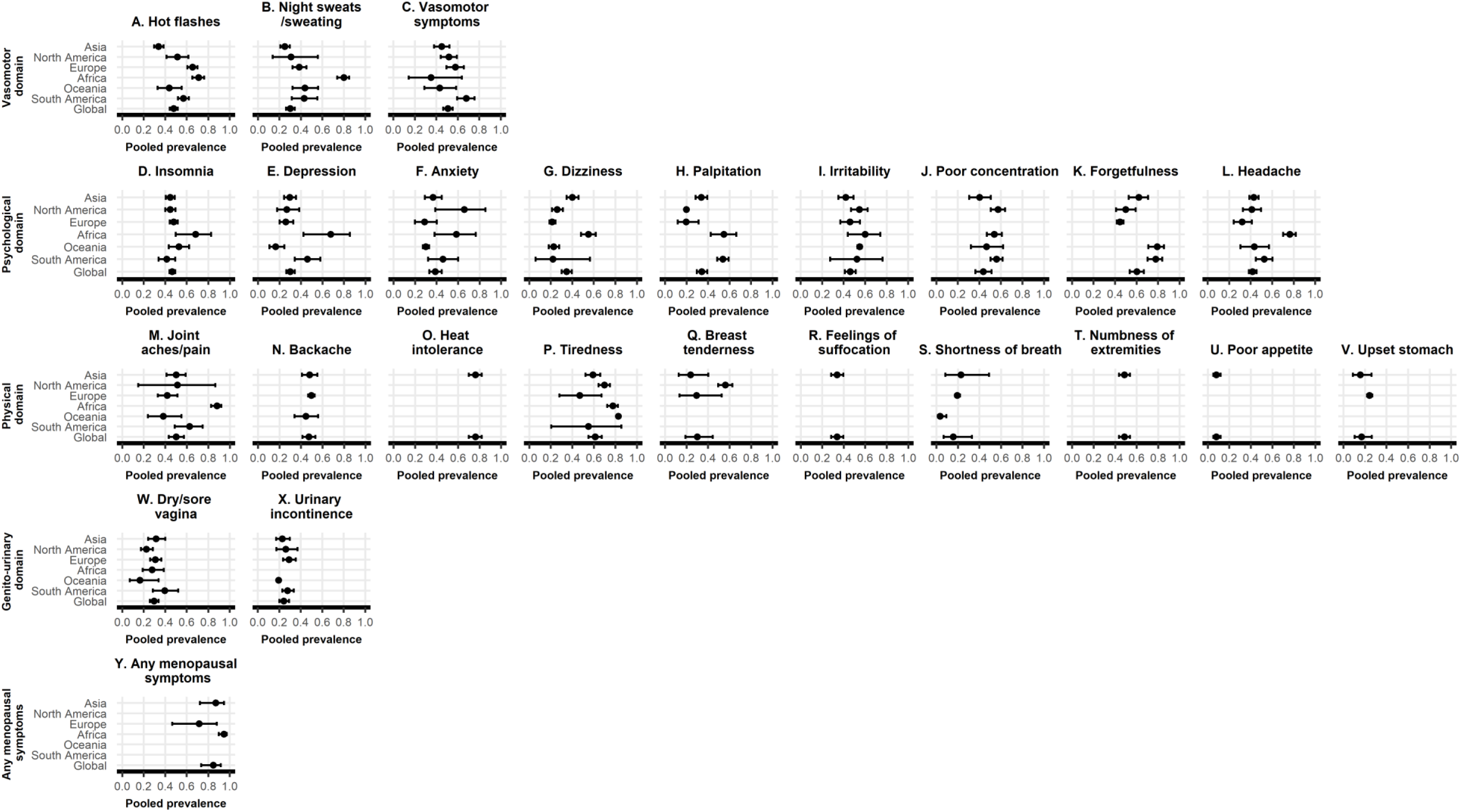
Pooled prevalences of 24 types of menopausal symptoms by continents.

### Prevalence of comorbidities in women with menopausal symptoms

Given the insufficient data on comorbidities in women experiencing menopausal symptoms, we opted to summarize the patterns descriptively in **Supplementary Table S6** rather than perform a pooled analysis. Key findings by 4 domains are as follows: **Vasomotor domain**: Hypertension prevalence ranged from 0.16 to 0.53; diabetes, 0.06 -0.09; obesity, 0.15-0.53; and hyperlipidemia, 0.34-0.45.

**Psychological domain**: Hypertension prevalence ranged from 0.18 to 0.50; diabetes, 0.03-0.14; obesity, 0.02-0.45; and hyperlipidemia, 0.13-0.50.

**Physical domain**: Limited data; obesity prevalence for stiffness/soreness in joints was 0.46 (BMI ≥27 kg/m²).

**Genito-urinary domain**: Hypertension prevalence was 0.26; diabetes 0.06; obesity 0.54 (BMI ≥27 kg/m²).

Interestingly, among women aged 40-55 years, the estimated prevalence of obesity (BMI ≥ 27 kg/m²) among those with and without vasomotor symptoms, urinary incontinence, dry/sore vagina, insomnia, and forgetfulness was as follows: 53.1% vs. 35.0%, 54.4% vs. 37.9%, 45.9% vs. 40.0%, 44.0% vs. 38.9%, and 45.1% vs. 38.0%, respectively. However, we were unable to exclude those non-target populations or events in this case^63^.

### Incidence and outcomes of menopausal symptoms

Due to the limited availability of data on the incidence and outcomes of menopausal symptoms, we refrained from conducting a meta-analysis and instead described their potential patterns. With respect to the incidence of menopausal symptoms (per 1000 person-years), joint pain (137.0), urinary incontinence (111.2), and vasomotor symptoms (105.9) demonstrated the highest rates, followed by urinary incontinence and insomnia, which exhibited high heterogeneity and warrant further investigations. In contrast, breast tenderness displayed the lowest incidence (23.1) (**Supplementary Table S7**). Regarding outcomes (**Supplementary Table S8**), we summarized the patterns across four domains:

**Vasomotor domain**: Associations with coronary heart disease and all-cause death were inconsistent; a marginal negative association with breast cancer was observed.

**Psychological domain** (depression and insomnia data): Consistent positive associations were noted with all-cause mortality and cardiovascular disease, but not with cancer.

**Physical domain** (joint pain data): positive associations were observed with coronary heart disease and fatal cardiovascular disease.

**Genito-urinary domain**: No outcome data available.

### Heterogeneity and publication bias

Almost all prevalences of the 24 types of menopausal symptoms exhibited substantial heterogeneity (I^2^ statistics > 90%, **Supplementary Table S5**). Univariate and multivariable meta-regression analyses indicated that continents and publication years significantly explained the heterogeneity in prevalences (*P* < 0.05) of most of the menopausal symptoms. However, a large portion of the heterogeneity remained unexplained (**Supplementary Table S9**). Stratified analysis by menopausal status (i.e., peri- and postmenopausal periods) did not significantly alter the level of heterogeneity (**Supplementary Table S5**).

The results of the Begg-Mazumdar Test and/or Egger Test suggested the possible existence of publication bias. Compared with the trim and fill method, the original analyses appeared to underestimate the prevalences of night sweats/sweating, anxiety, and joint aches/pain by 8-17%, while overestimating the prevalence of vasomotor symptoms, insomnia, depression, palpitations, forgetfulness, headache, tiredness, and any menopausal symptoms by 9-22% (**Supplementary Table S10**).

## Discussion

In this study, we extracted data from 239 papers regarding menopausal symptoms, encompassing over 2.5 million women. This is thus far the largest systematic review and meta-analysis of prevalence, comorbidities, incidence and outcomes of menopausal symptoms in peri- and postmenopausal women worldwide. We found that the majority (85%) of global peri- and postmenopausal women experienced menopausal symptoms, thus consolidating earlier research^64^. Additionally, around 50% of women suffered from vasomotor symptoms. Prevalences of most menopausal symptoms, were comparable in peri-and postmenopausal women. We summarized the patterns of prevalence, comorbidities, incidence and outcomes of menopausal symptoms.

Previous reviews have summarized the prevalence of menopausal symptoms, reporting widely varied estimates. However, most of them were narrative reviews^24-28^ or systematic reviews with a narrow scope, limited numbers of studies, high heterogeneity, and a high risk of bias^29, 30^. In summary, these reviews reported that the prevalence of vasomotor symptoms ranged from 13% to 75%, psychological symptoms from single digits to over 80%, and physical symptoms from 5% to over 90%, with marked variation by geography and menopausal stage. However, these reviews are either constrained by narrative synthesis or limited in scope and areas, which makes it difficult to capture the global patterns of menopausal symptoms. Our systematic review and meta-analysis differ in several aspects. First, we included 239 papers encompassing approximately 2.5 million women and calculated the pooled prevalences of 24 types of menopausal symptoms. Second, we described the trends in the prevalences of these menopausal symptoms by publication years and country, adding this evidence, to the best of our knowledge, for the first time. Third, the prevalences of 24 types of menopausal symptoms was pooled by continents and menopausal stage, revealing the comprehensive characteristics of these symptoms. Fourth, we ranked the pooled prevalence estimates of 24 types of menopausal symptoms using global data, which we consider comparable given the similarities in sex and age across populations. Fifth, we extended the synthesis beyond prevalence to include comprehensive data on comorbidities, incidence and outcomes of menopausal symptoms and summarized their patterns, although the data remain limited. This aspect was largely unaddressed in previous reviews. Overall, despite a substantial heterogeneity, acknowledged by prior reviews as well, our study provided the global landscape of the full spectrum of menopausal symptoms, highlighting the burden of specific symptoms, which may thereby help to define more concrete clinical and public health priorities.

Our findings showed that the distribution of different menopausal symptoms varied across the six continents (Figure 4). We found that women from Africa have the highest pooled prevalence of hot flashes, night sweats, insomnia, depression, dizziness, headache, joint pain and any menopausal symptoms, compared to other continents. However, we believe that more studies from Africa are necessary to confirm our results, as data from Africa is limited.

An important aspect to highlight is the heterogeneity in the diagnostic tools for menopausal symptoms across different studies. Even when the measurement instruments share the same menopausal symptom labels (**Supplementary Table S3**), the content of the questionnaires may vary^65, 66^. Furthermore, the variety of diagnostic tools is extensive. Hence, we could not investigate the heterogeneity of prevalence rates attributed to diagnostic tools in the meta-regression (**Supplementary Table S9**). The heterogeneity of diagnostic tools may stem from variations in the years in which the studies were conducted and from cultural contexts. New tools for measuring menopausal symptoms continue to be developed. As a result, studies conducted at different time points have used different tools. In addition, investigation on sexual issues is more difficult in countries or regions with a relatively closed culture compared to developed countries, yielding differences in questionnaires. To mitigate the influence of heterogeneity of diagnostic tools, we only focused on the presence or absence of menopausal symptoms, rather than the distribution of mild, moderate, and severe cases. In the future, it would be beneficial to develop and promote a standardized diagnostic tool for measuring menopausal symptoms that is applicable to a wide range of countries and regions.

We found that the proportions of comorbidities, including hypertension, obesity and hyperlipidemia, reached up to 50% in women experiencing menopausal symptoms. Prior studies supported these findings. Our previous cross-sectional study, based on 196 Chinese perimenopausal women, found some evidence for a stronger inverse association between total bilirubin and hypertension in women with fewer menopausal symptoms (OR = 0.90 per μmol/L total bilirubin; 95% CI, 0.81-0.99) compared to those with more symptom (OR = 0.96 per μmol/L total bilirubin; 95% CI, 0.87-1.04)^40^, indicating menopausal symptoms may play a role in the development of hypertension.

Additionally, Sylvio et al.^67^ conducted a cross-sectional study in 749 Brazilian women and found a positive association between hot flashes and BMI (B = 0.03 kg/m^2^, standard deviation = 0.01, *P* < 0.05). A cross-sectional study encompassing 5,523 Dutch women revealed that women with hot flashes/flushing had higher cholesterol level (B = 0.27 mmol/L, 95% CI: 0.15-0.39), higher BMI (B = 0.60 kg/m^2^, 95% CI: 0.35-0.84), higher odds of hypercholesterolemia (OR = 1.52, 95% CI: 1.25-1.84) and higher odd of hypertension (OR = 1.20, 95% CI: 1.07-1.34). Results for night sweats were similar^21^. Interestingly, we found that the prevalence of diabetes among women experiencing menopausal symptoms was up to 15%. Despite this, Gast et al. reported that Australian women (n=4,895) with severe vasomotor symptoms had a higher risk of diabetes (OR = 1.55, 95% CI: 1.11-2.17)^68^.

We found that joint pain in the US, vasomotor symptoms in the Netherlands, urinary incontinence in the US and insomnia in the US are the symptoms with the highest incidence rates (all > 90 per 1000 person-years, **Table S7**). Future research should investigate the incidence of other menopausal symptoms within cohort studies. Prior studies have focused on the impact of menopausal symptoms on quality of life^69^ and work ability^4^, which may be deemed less important when compared to the mortality and years of life lost caused by cancer^70^ and cardiovascular diseases^71^. Despite some inconsistent findings regarding the relationship of vasomotor symptoms with coronary heart disease and all-cause mortality, our study did find association with coronary heart disease of some other menopausal symptoms such as depression and joint pain. This highlights the importance of menopausal symptoms and warrants prioritizing their management, for example, by promoting guidelines^27, 28^. Of note, hormone therapy has been the traditional treatment for vasomotor symptoms; however, its use is associated with an increased risk of breast cancer, venous thromboembolism and stroke. Although alternative options, such as herbal therapies, are available in this case, their clinical efficacy is limited^72^. In recent years, the FDA approved Veozah (Fezolinetant) as a nonhormone treatment for vasomotor symptoms. As the first nonhormonal therapy capable of crossing the blood–brain barrier, fezolinetant has been mentioned as representing a breakthrough in the management of menopausal symptoms, despite its side effects that include abdominal pain, diarrhea, insomnia, backache, hot flashes, and increased hepatic transaminases^72^. Understanding of the underlying mechanisms linking menopausal symptoms to outcomes remains limited warranting further studies.

In meta-analysis, publication bias can lead to an underestimation or overestimation of the pooled effect size. However, it is difficult to employ reliable methods for assessing publication bias when there is high heterogeneity^58^. Given the significant heterogeneity observed in almost all menopausal symptoms in this study, there are currently no acceptable methods to assess publication bias. Despite this, we proceeded to utilize traditional methods, including the Begg-Mazumdar Test and Egger Test, to evaluate potential publication bias. The results of these tests only provide an indication, because heterogeneity can impair the performance of both tests and bias the estimates^73^. Of note, there was large heterogeneity in prevalence estimates between studies. This heterogeneity might be partly attributable to different ethnic, cultural, geographic and climate features and also differences between instruments to measure menopausal symptoms. Although we conducted stratified analysis and meta-regression to explore the heterogeneity, substantial unexplained heterogeneity remained.

The current study has a number of strengths. For example, we systematically collected data from a considerable number of studies with a combined sample size of over 2.5 million, thereby enabling us to provide a comprehensive assessment on the global data of 24 types of menopausal symptoms. In addition, we distinguished between peri- and postmenopausal women, which has important implications for exploring variation in menopausal symptoms between these two distinct phases.

There are also several limitations in the present study. First, the prevalence of certain menopausal symptoms, such as dry/sore vagina, may be underestimated in some countries due to cultural factors. Women from more conservative countries are more likely to decline participation in such surveys. Second, the data on prevalence, incidence and outcomes of menopausal symptoms from Africa is limited, which may pose a challenge to accurately assess the pooled prevalence of menopausal symptoms globally. Third, the majority of the included studies involved non-target populations or events, because hormone therapy is widely used in women worldwide and premenopausal women were not excluded in most of studies. However, we limited the proportion of non-target populations or events to below 40% in the studies included. Fourth, we categorized hypertension, diabetes, obesity, and hyperlipidemia as comorbidities of menopausal symptoms whereas stroke, coronary heart disease, leiomyoma, breast, cervical, ovarian, uterine, and endometrial neoplasms, and all-cause mortality were considered as outcomes. However, these distinctions between comorbidities and outcomes are not absolute and should be considered exploratory. Fifth, we did not extract data to investigate whether women with menopausal symptoms experience more or fewer comorbidities compared to those without. Sixth, we did not assess the impact of menopausal symptoms on quality of life or work ability. Finally, in spite of the comprehensive body of evidence this systematic review and meta-analysis brought together, we recognize that we have not been able to incorporate the most recent literature as our literature search ended on March 1^st^ 2019. As such, confirming our findings in the latest literature should be a major priority of future work. Our study draws a panoramic picture of 24 types of menopausal symptoms according to menopausal status on a global scale, providing key information to support better management of menopausal symptoms for these women. Specifically, policymakers could prioritize addressing symptoms with higher prevalence among these women.

In conclusion, menopausal symptoms are ubiquitous in peri- and postmenopausal women, with the pooled prevalence of most symptoms being comparable between these two groups. The pooled prevalence rates of menopausal symptoms varied across the six continents, with higher prevalence in developing countries, especially for psychological and physical symptoms; significant variation in Asia, particularly in vasomotor symptoms; and notable regional differences in vasomotor symptoms. The findings highlight the need for broader research inclusion, particularly to address gaps in Africa, and for further study of comorbidities, incidence, outcomes, and underlying factors behind these disparities (i.e., variations in prevalence across continents and countries). Elucidating these factors will enhance the management of menopausal symptoms. It is also vital to prioritize high-burden symptoms in health policy and clinical planning. For example, African women should be given more attention regarding the burden of menopausal symptoms; women from Europe, Africa, South and Southeast Asia (e.g., Nepal, Thailand) should be carefully assessed for vasomotor symptoms; in developing countries, psychological and physical symptoms should be a focus during physician consultations.

## Author Approval

All authors have seen and approved the manuscript.

## Acknowledgement

We thank Dr. Fan Zhang and Dr. Rujia Wang for their kind assistance with this study. The protocol of this study was registered with the International Platform of Registered Systematic Review and Meta-Analysis Protocols (INPLASY, https://inplasy.com/wp-content/uploads/2020/12/INPLASY-Protocol-1170.pdf) on 12 December 2020 (registration number INPLASY2020120074).

## Data Availability Statement

All data are available in the supplementary tables and figures.

## Appendix 1. Pooled prevalence ranking of menopausal symptoms in different countries

**Figure S1.**
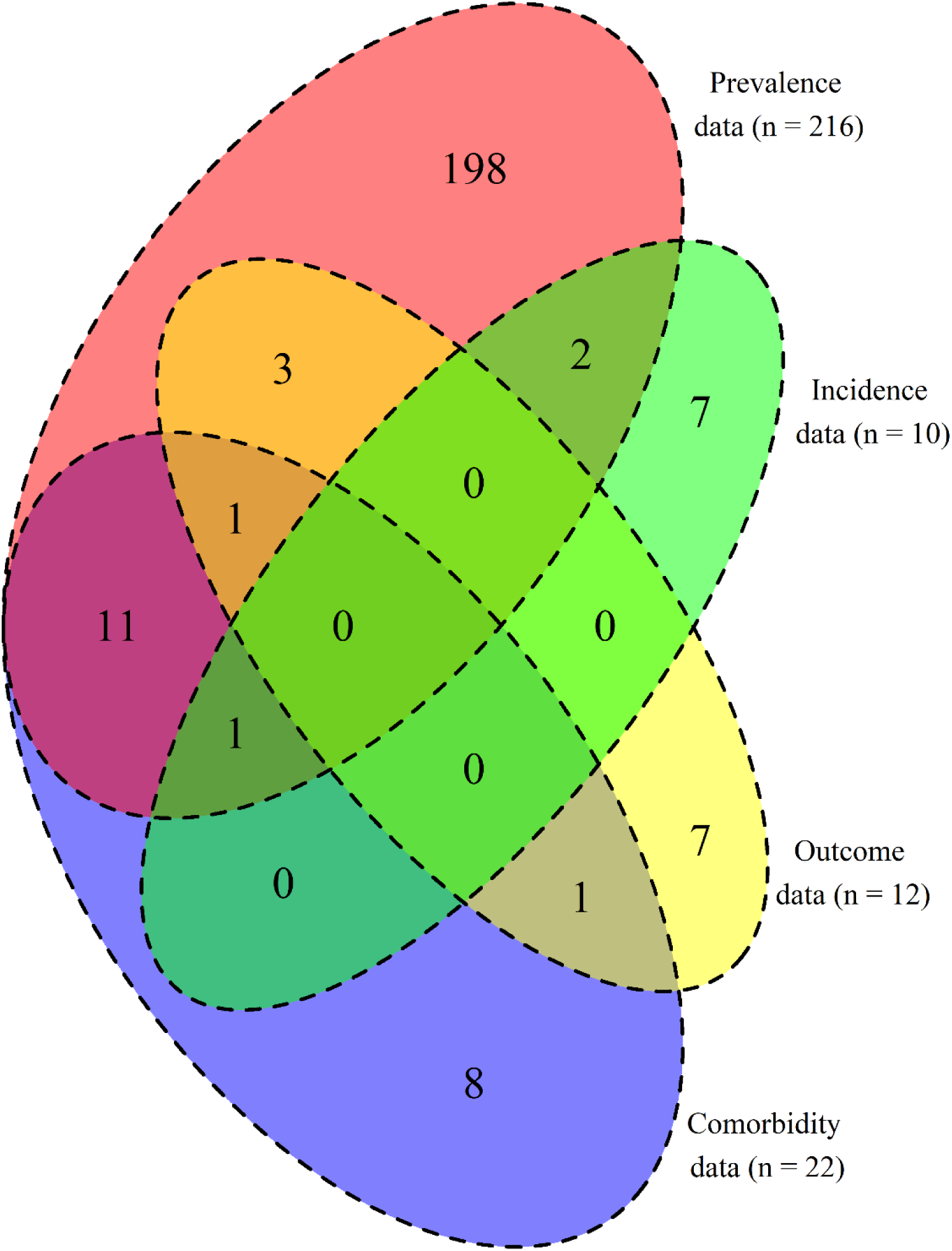
Venn diagram illustrating the results of this study.

**Figure A-Y:**
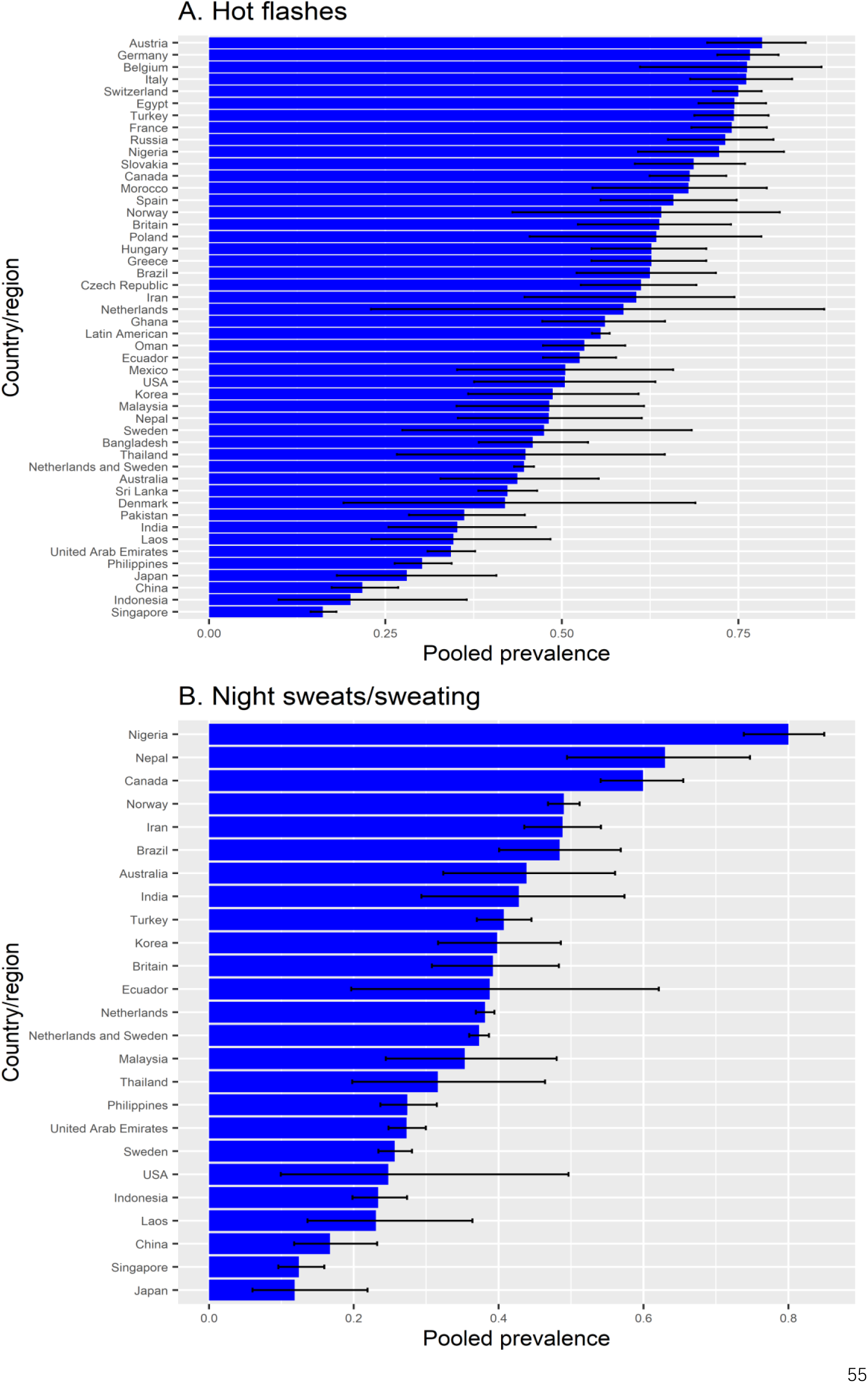
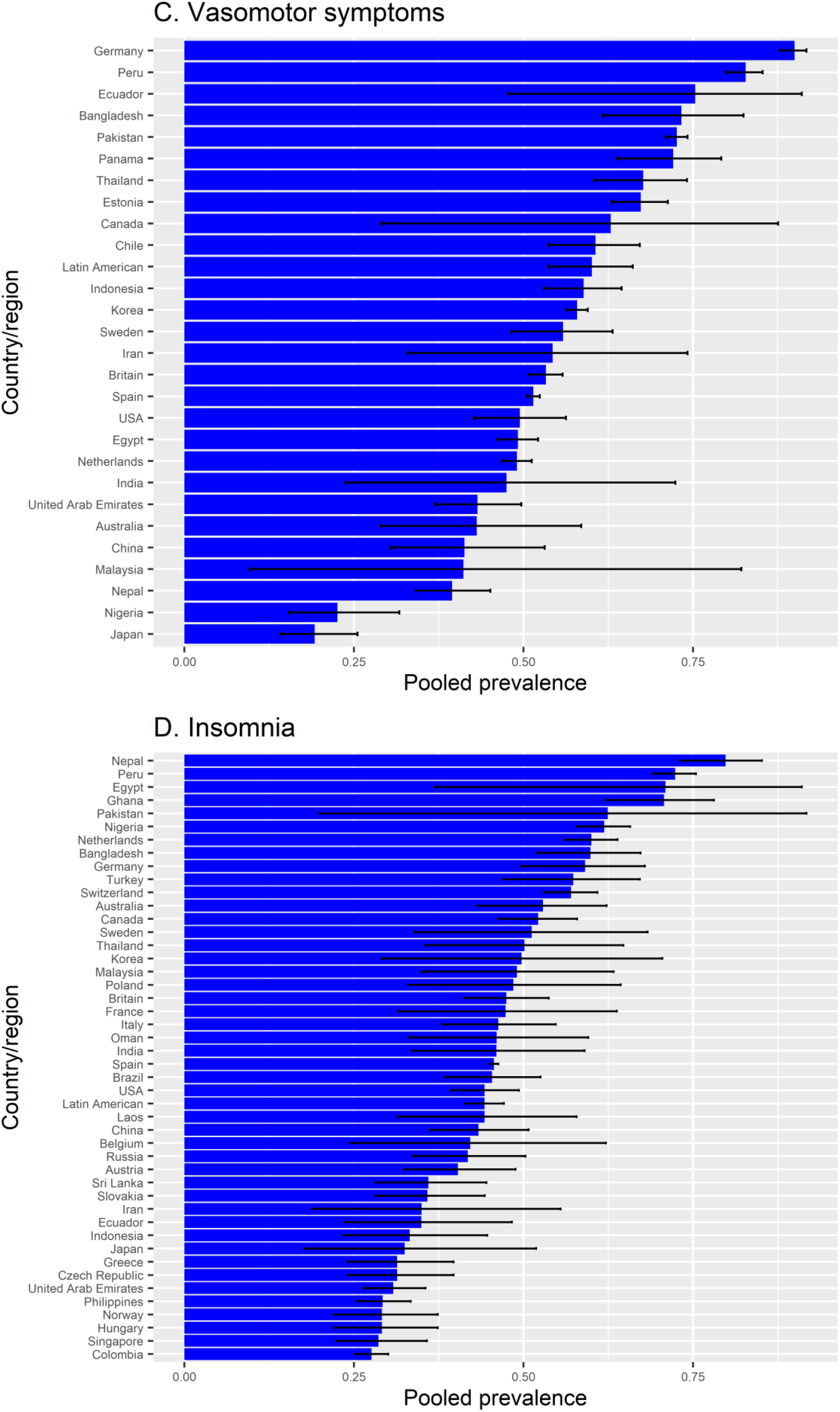
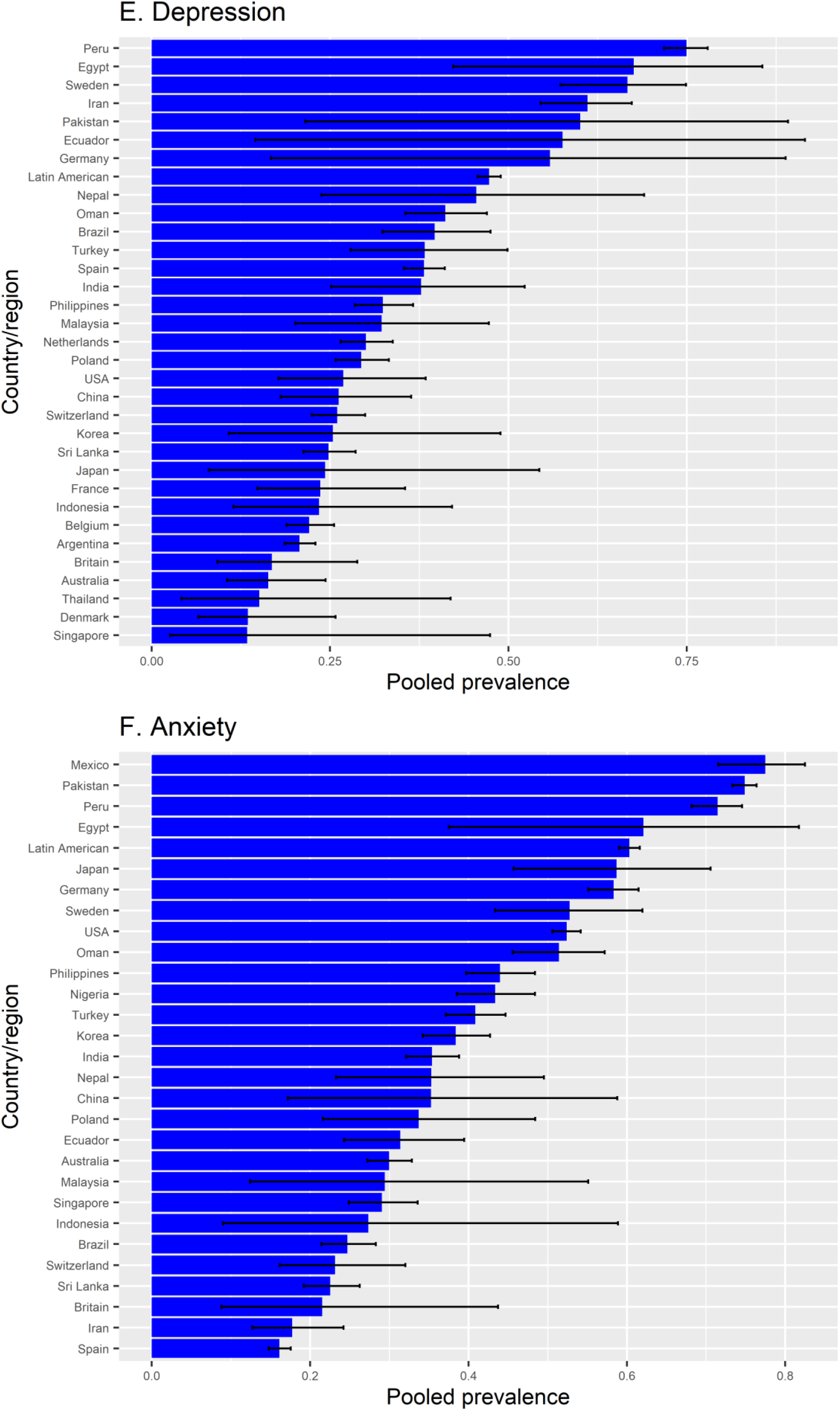
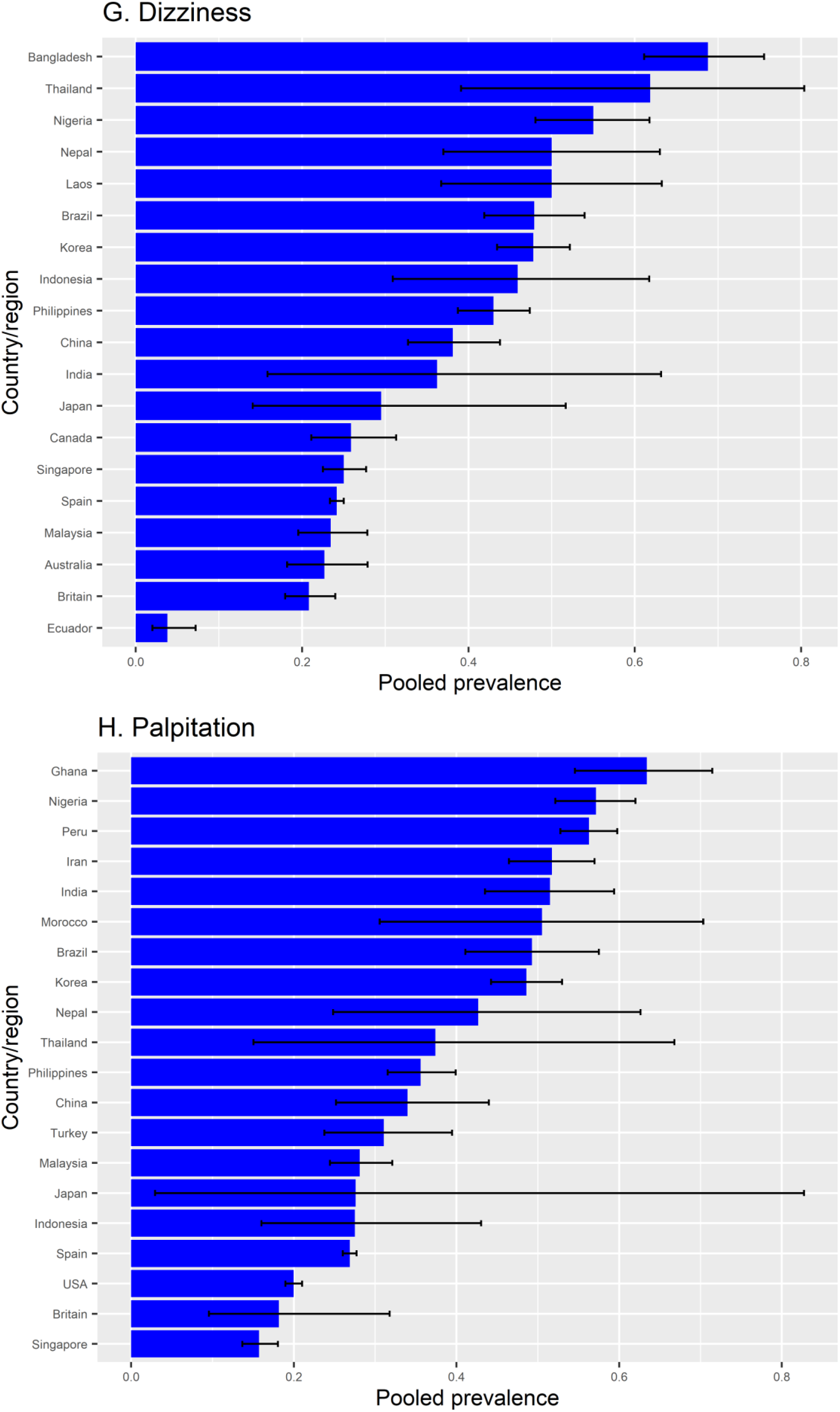
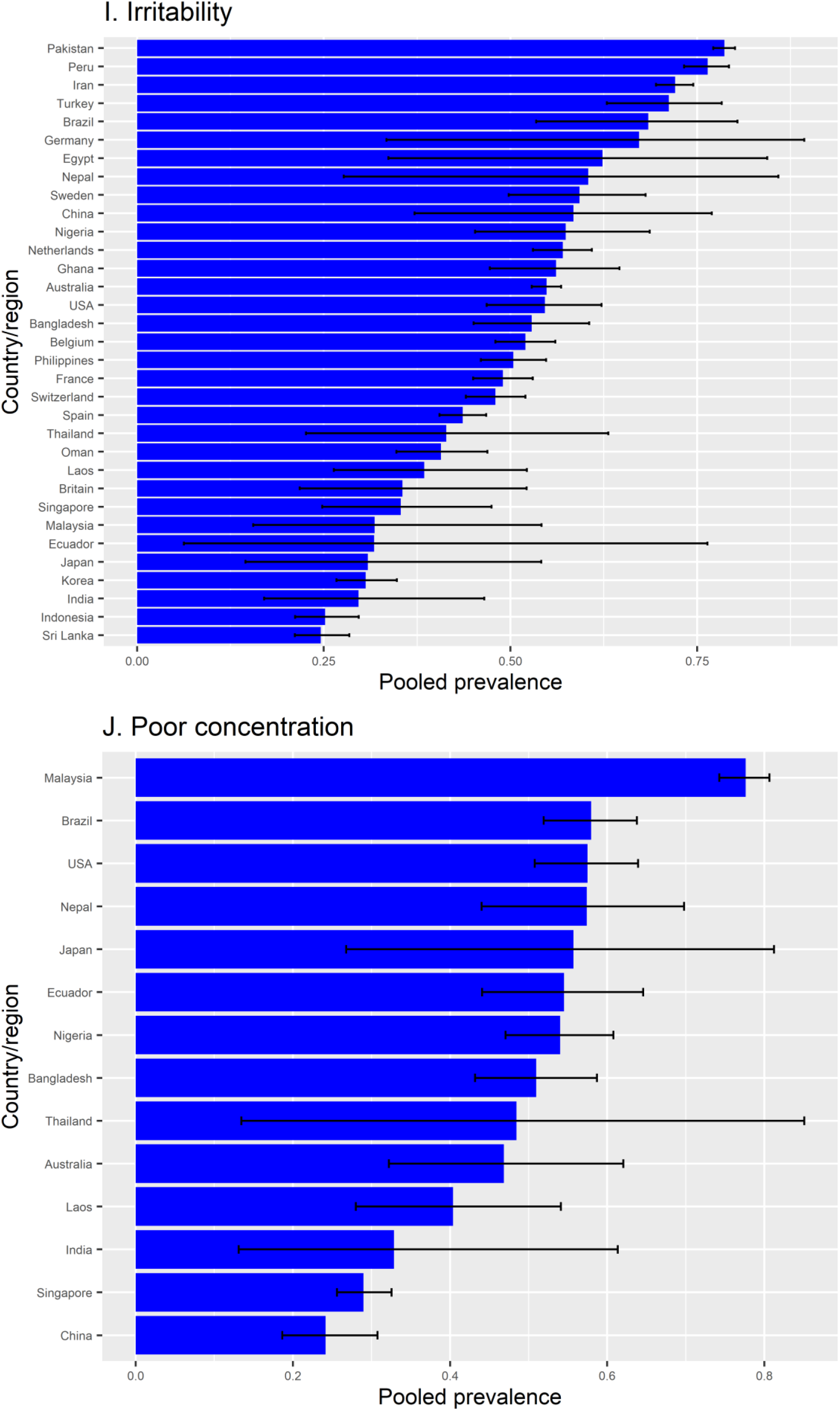
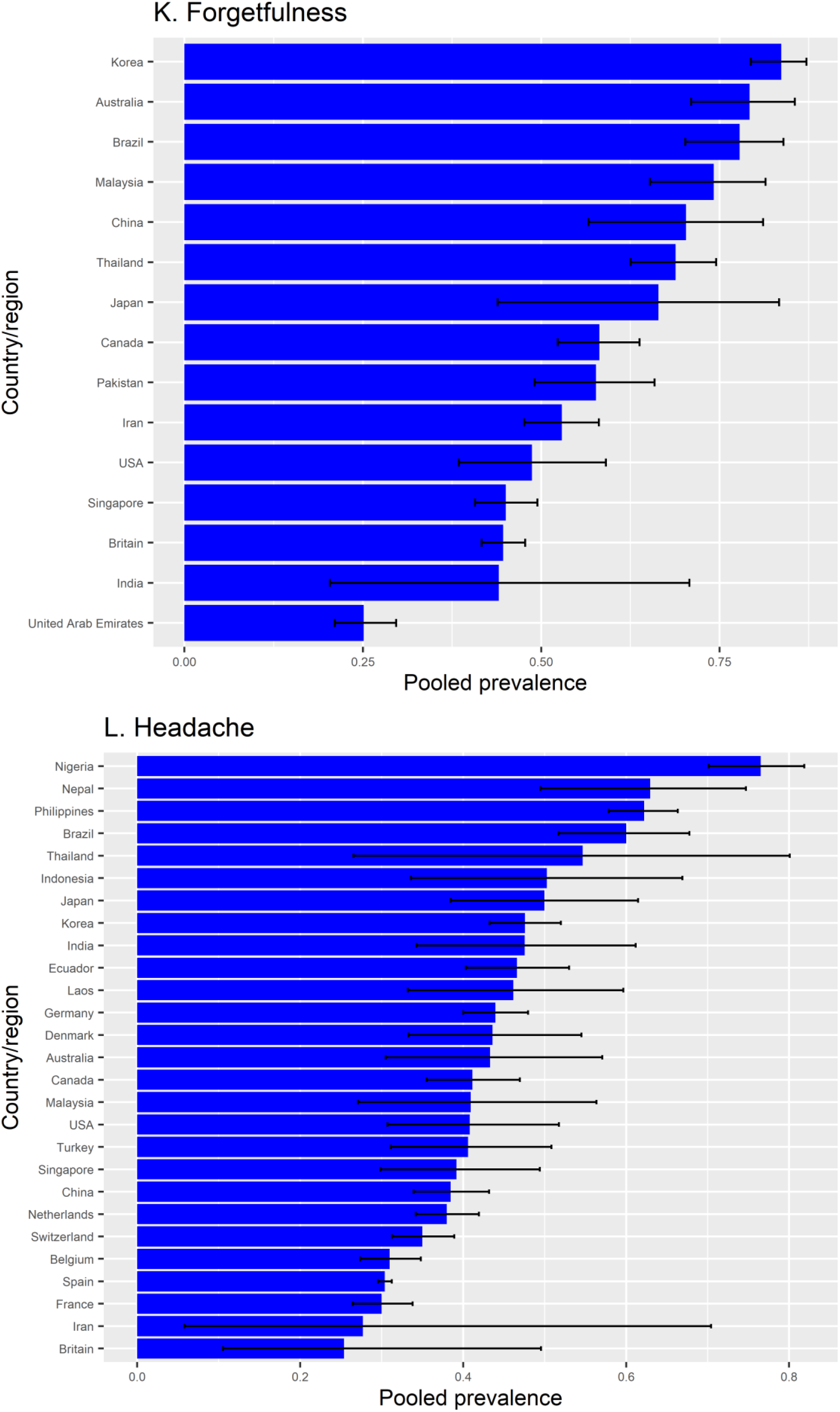
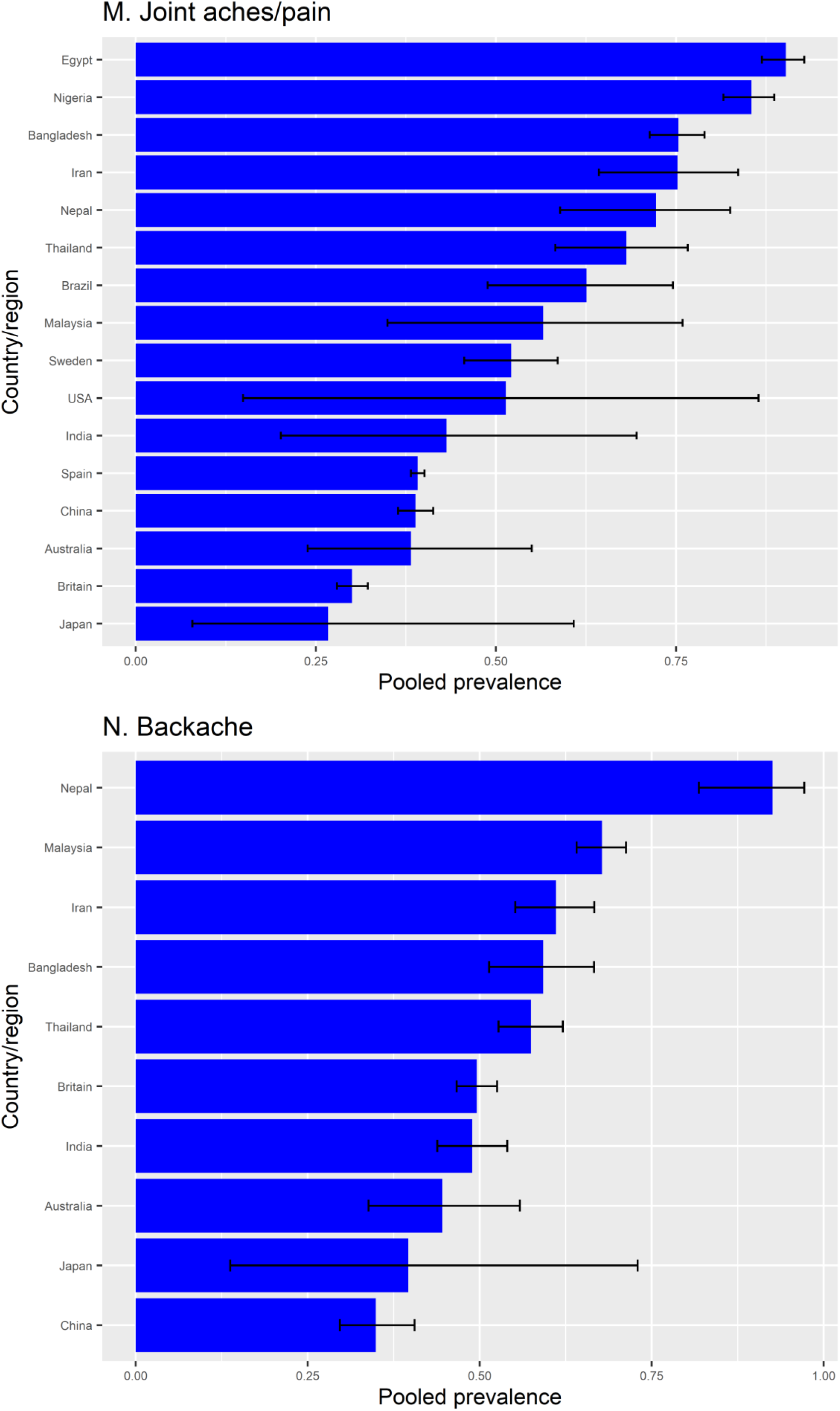
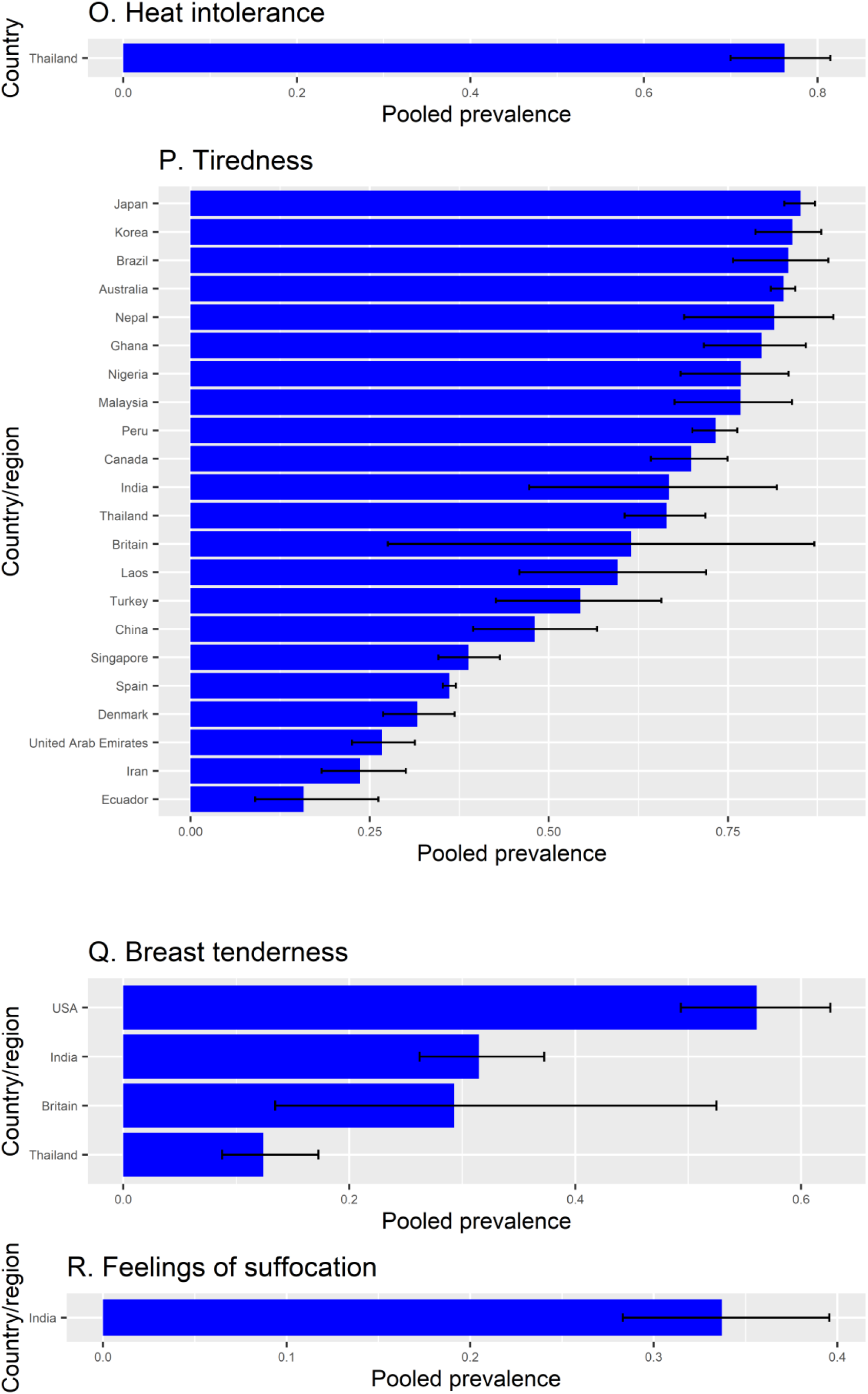
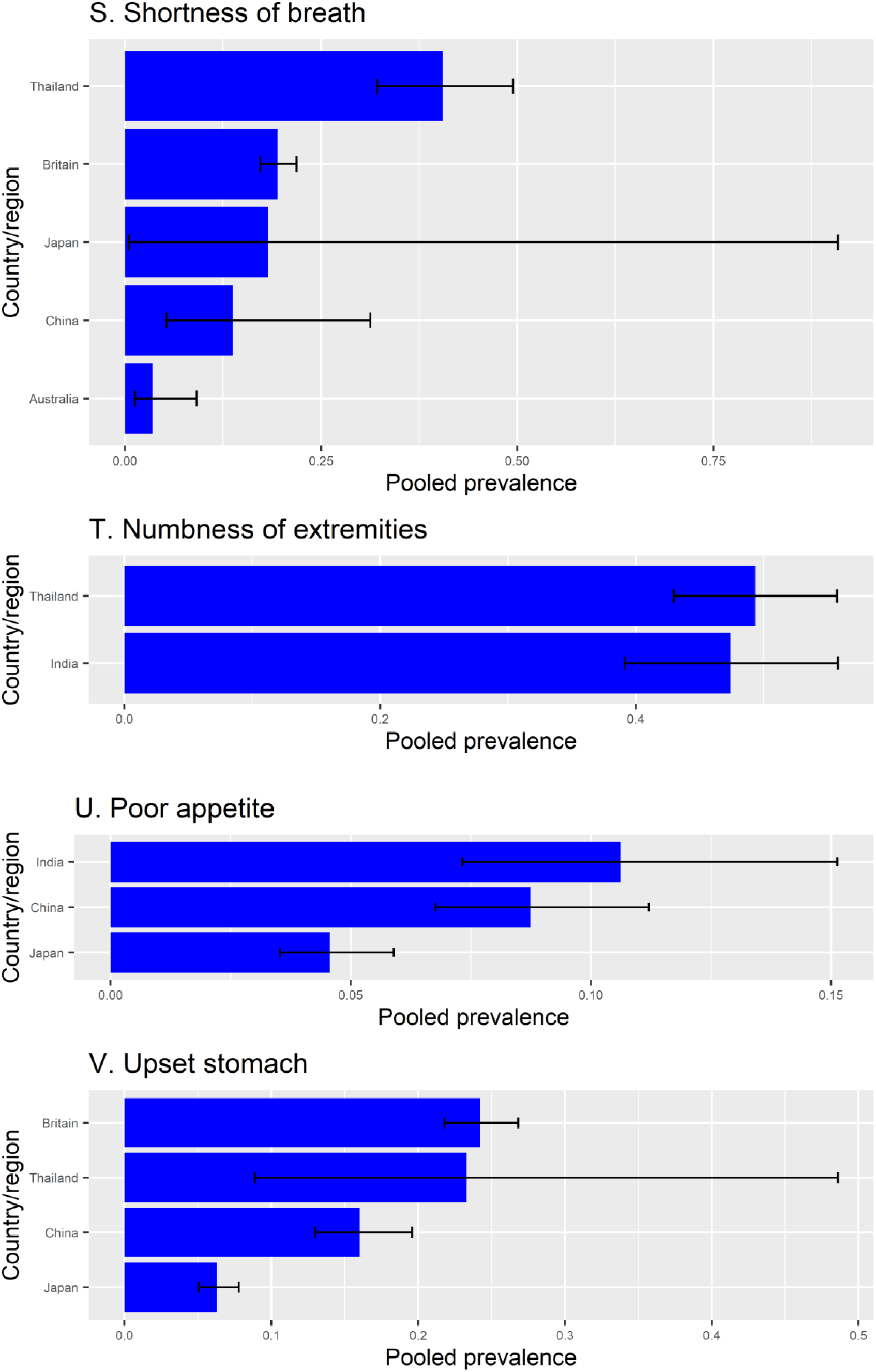
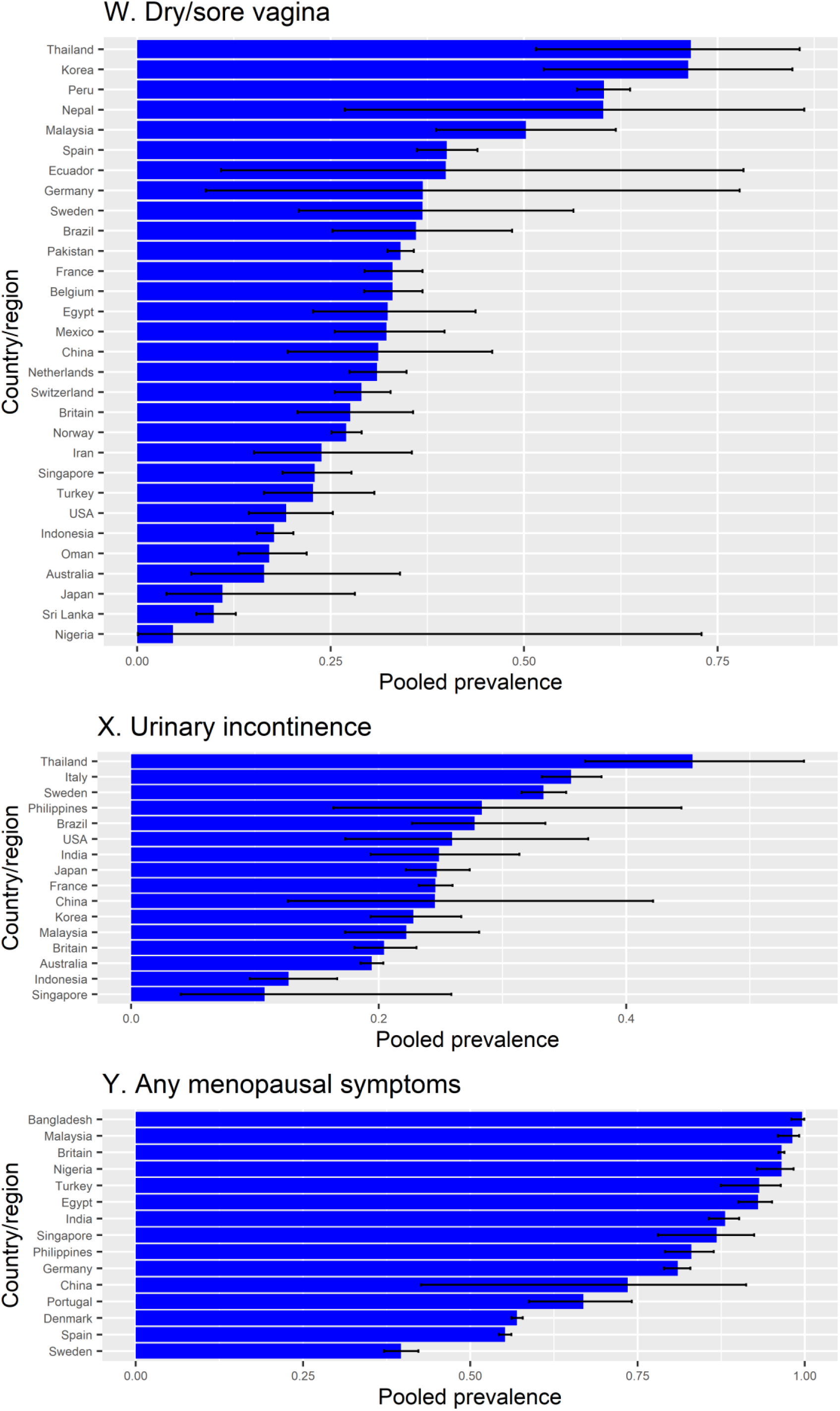
Bar chart of 24 menopausal symptoms by country. Error bars indicate 95% confidence intervals.

## Appendix 2. Trends in Menopausal Symptoms Across Countries Over Publication Years

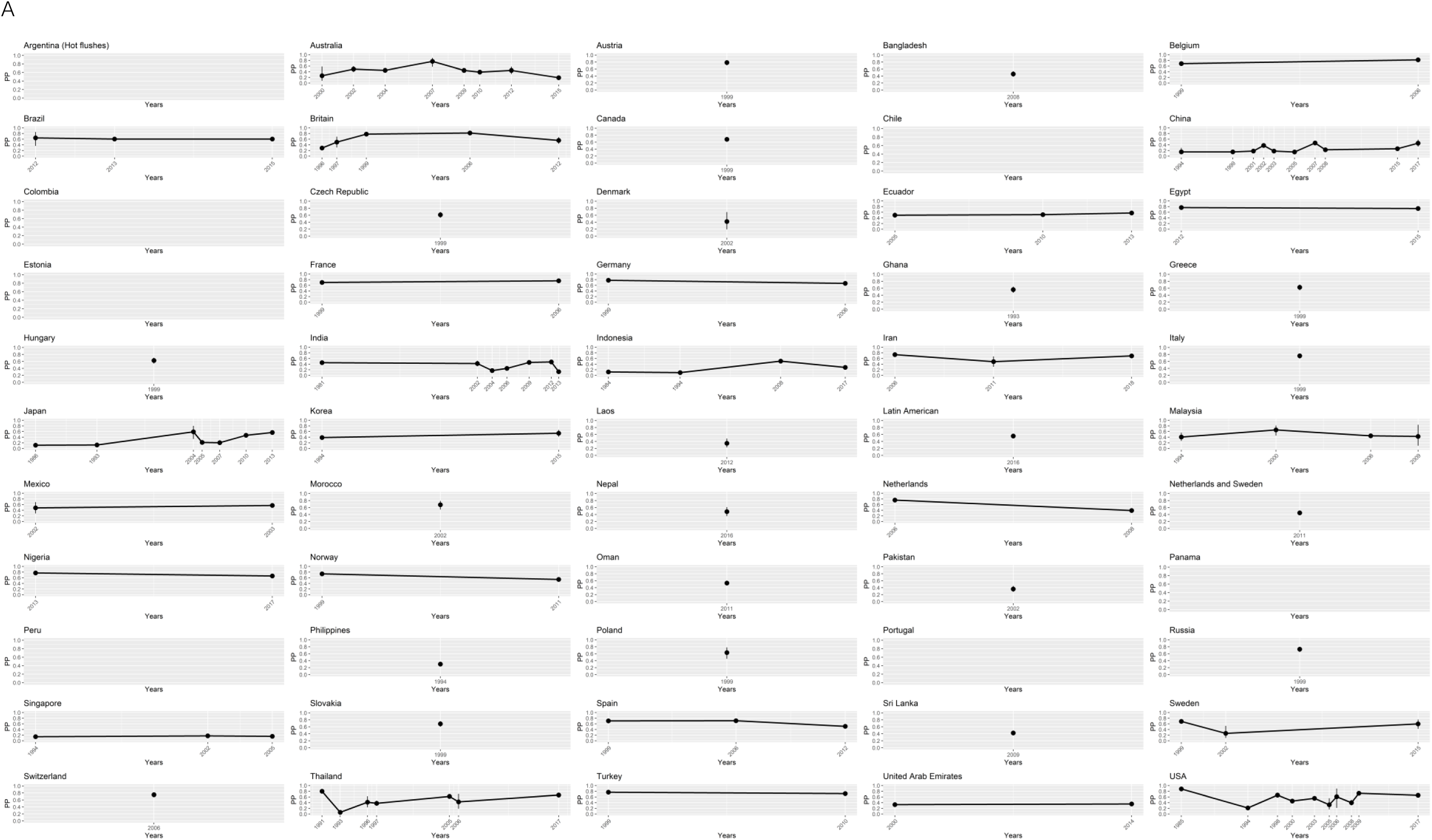
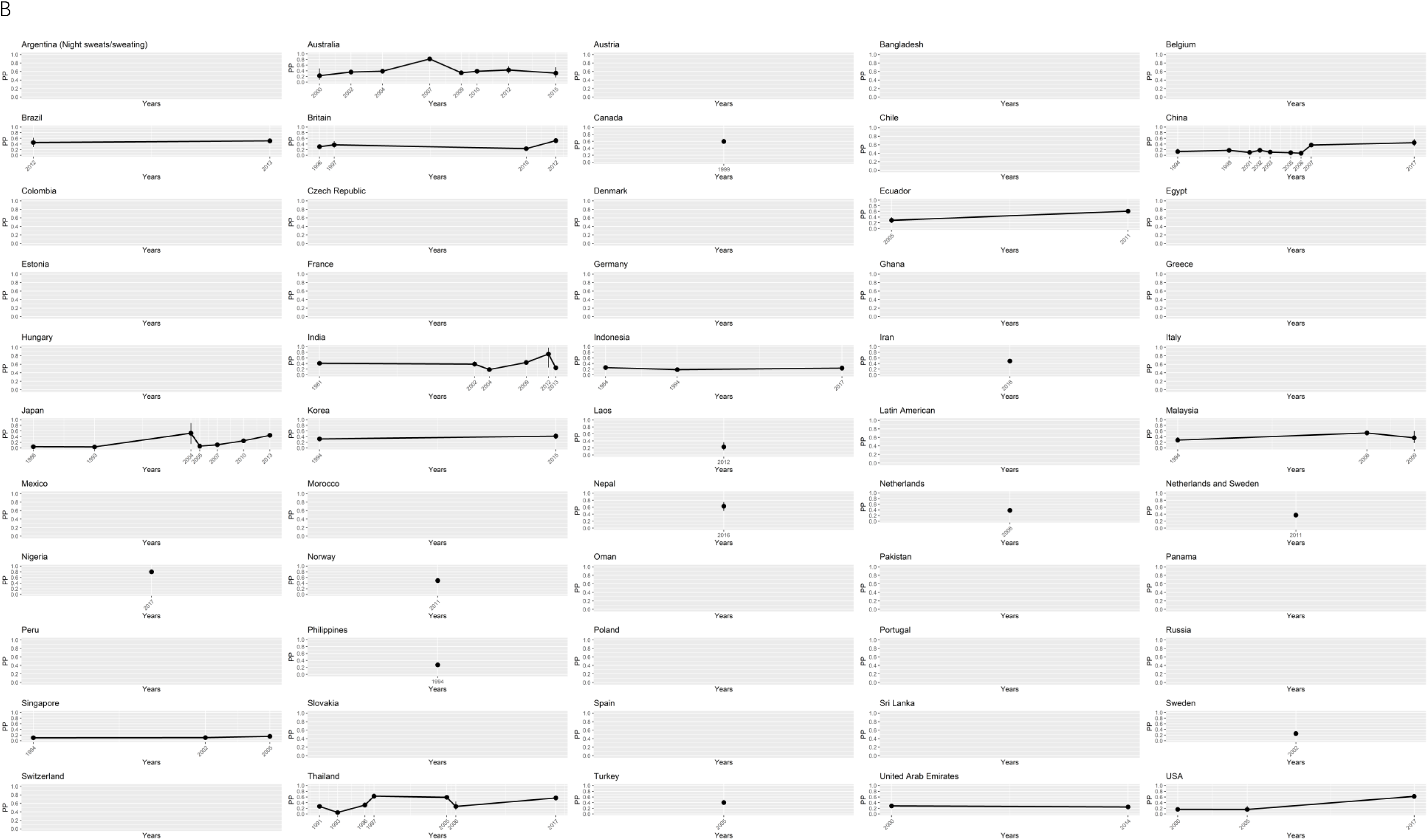
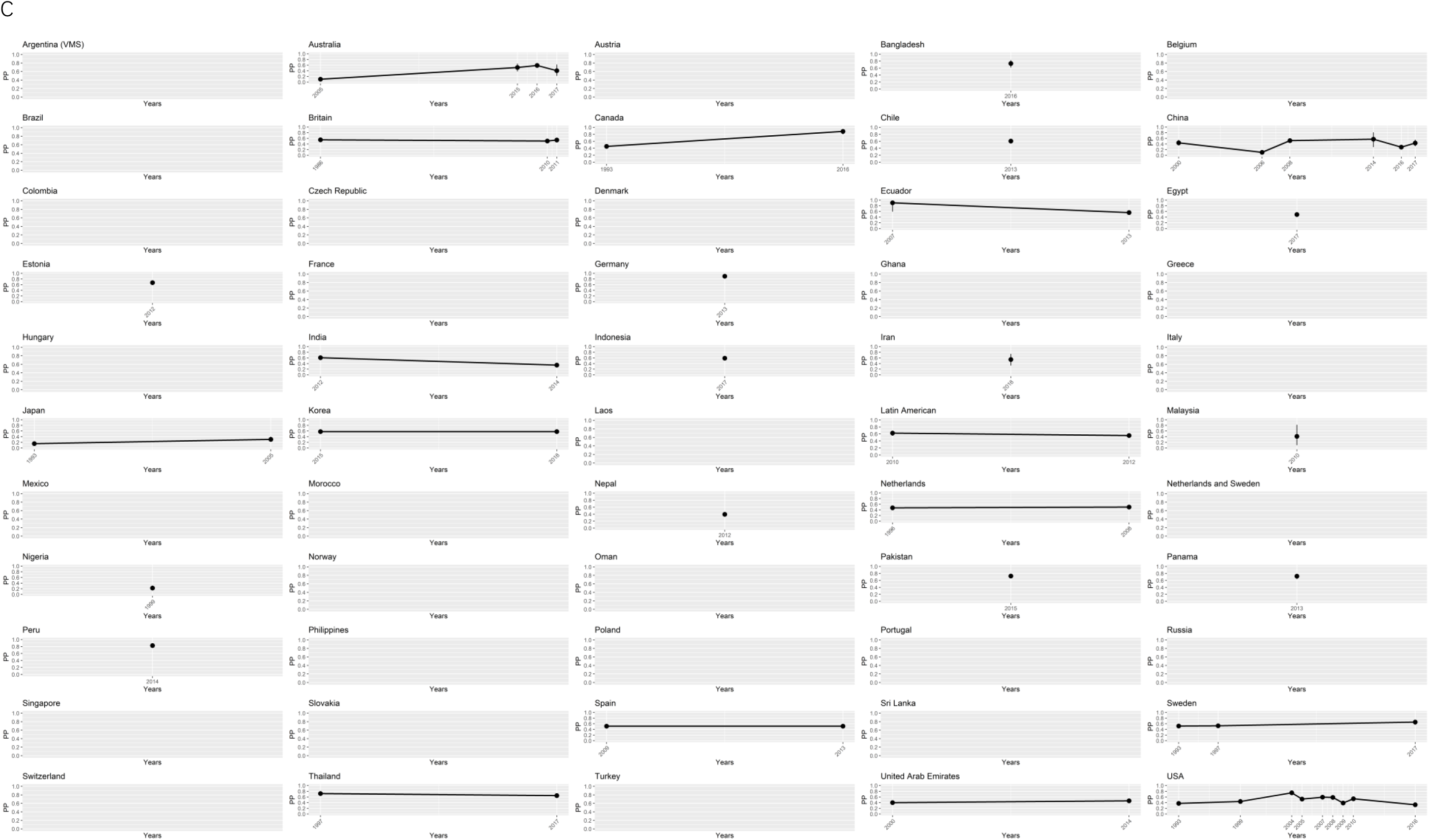
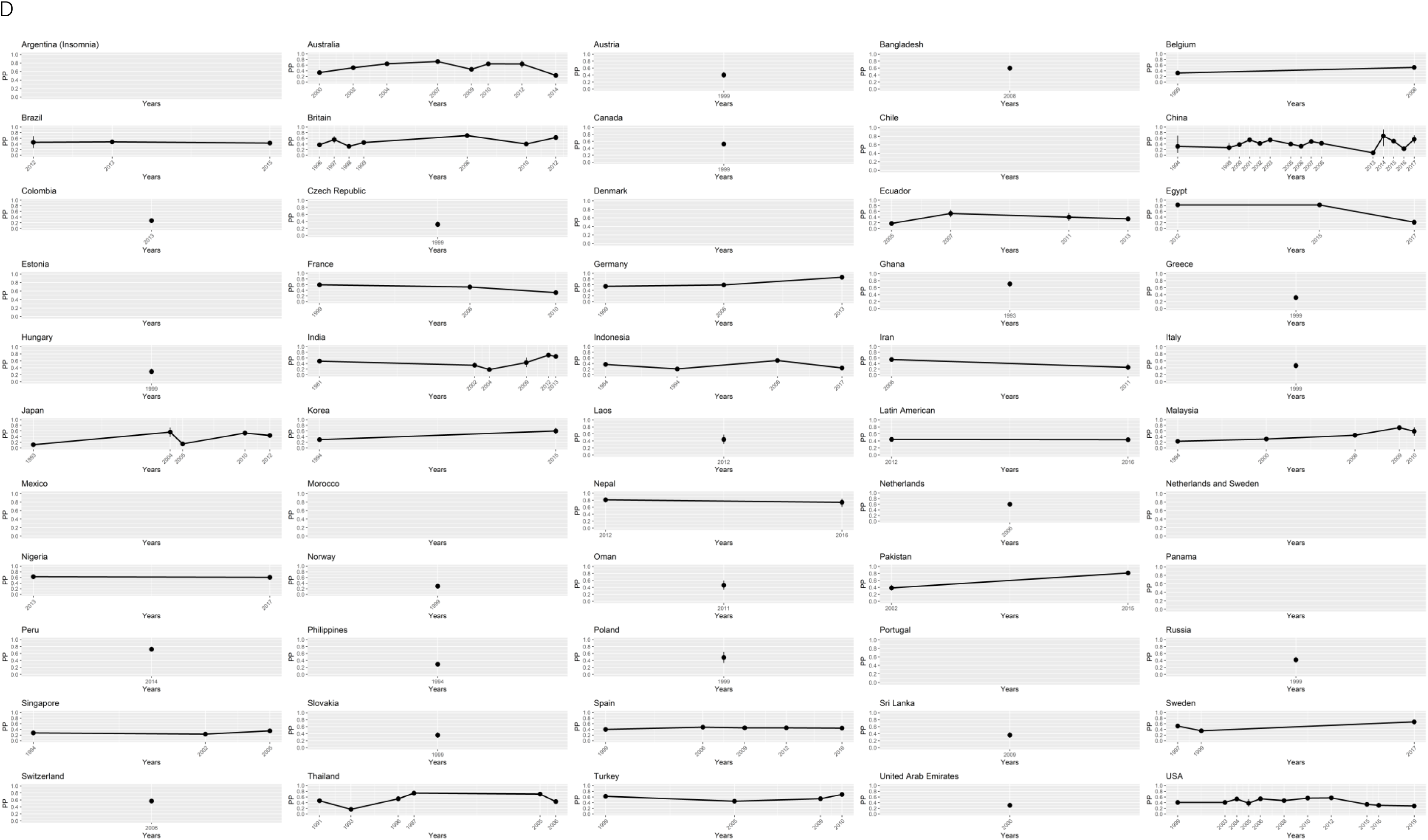
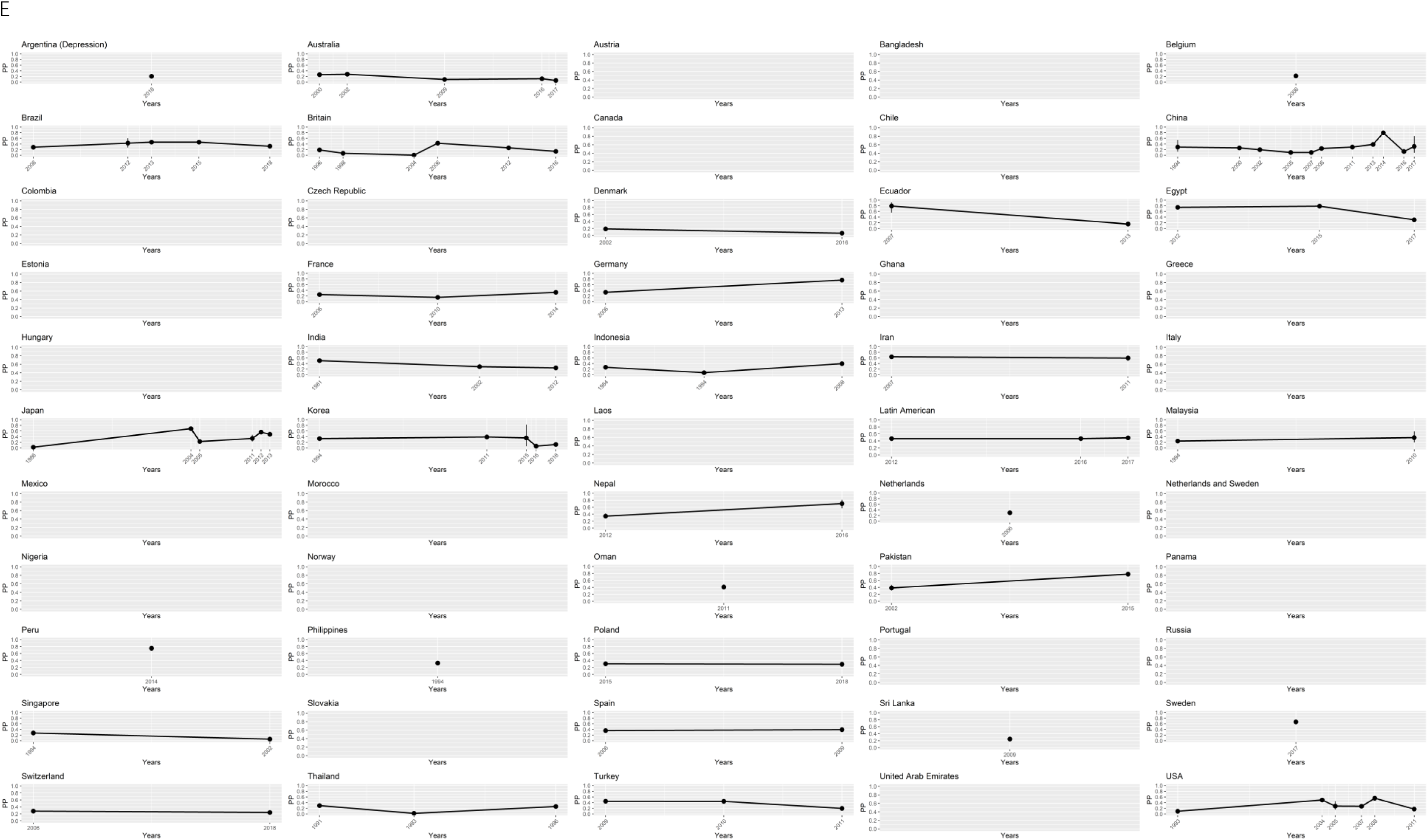
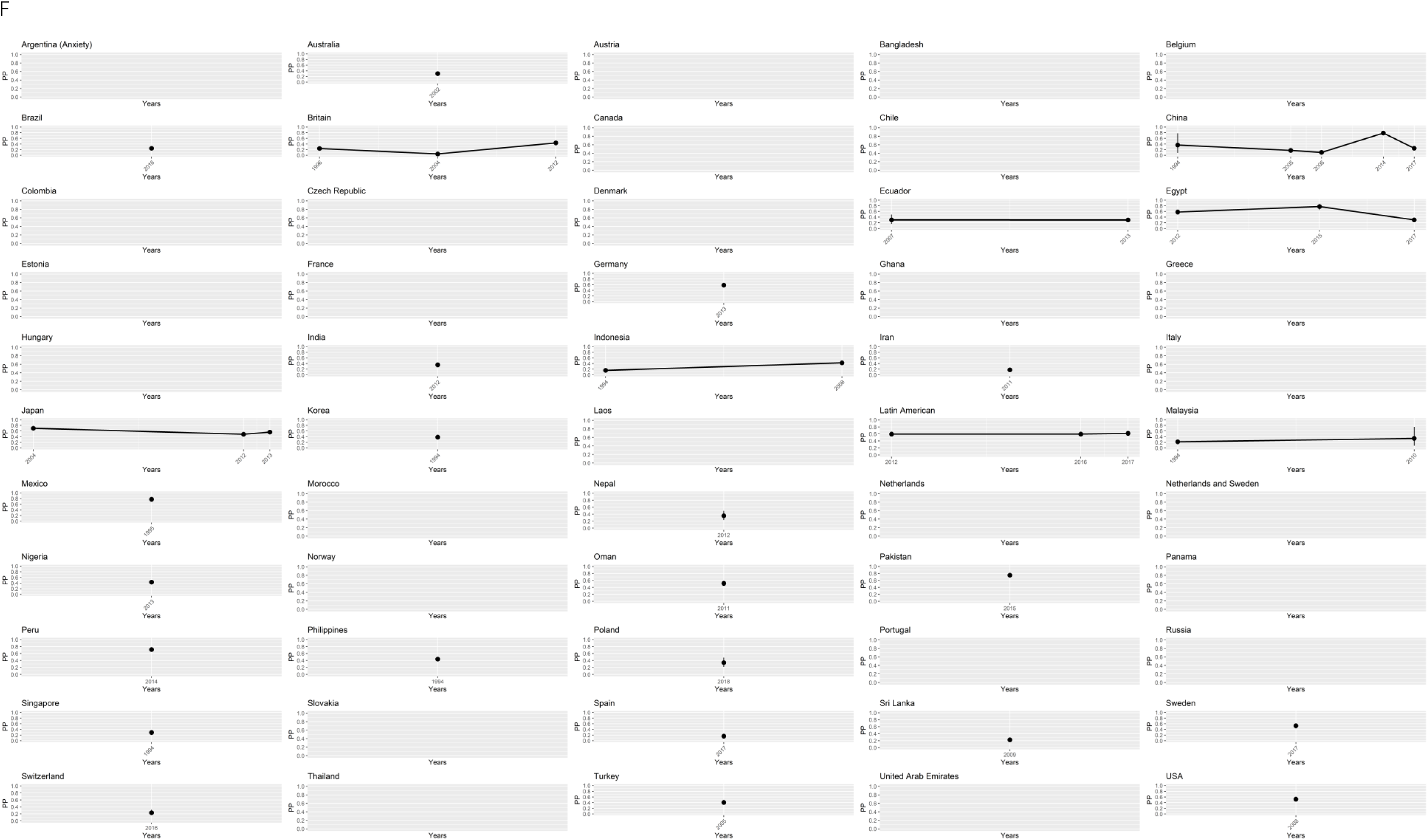
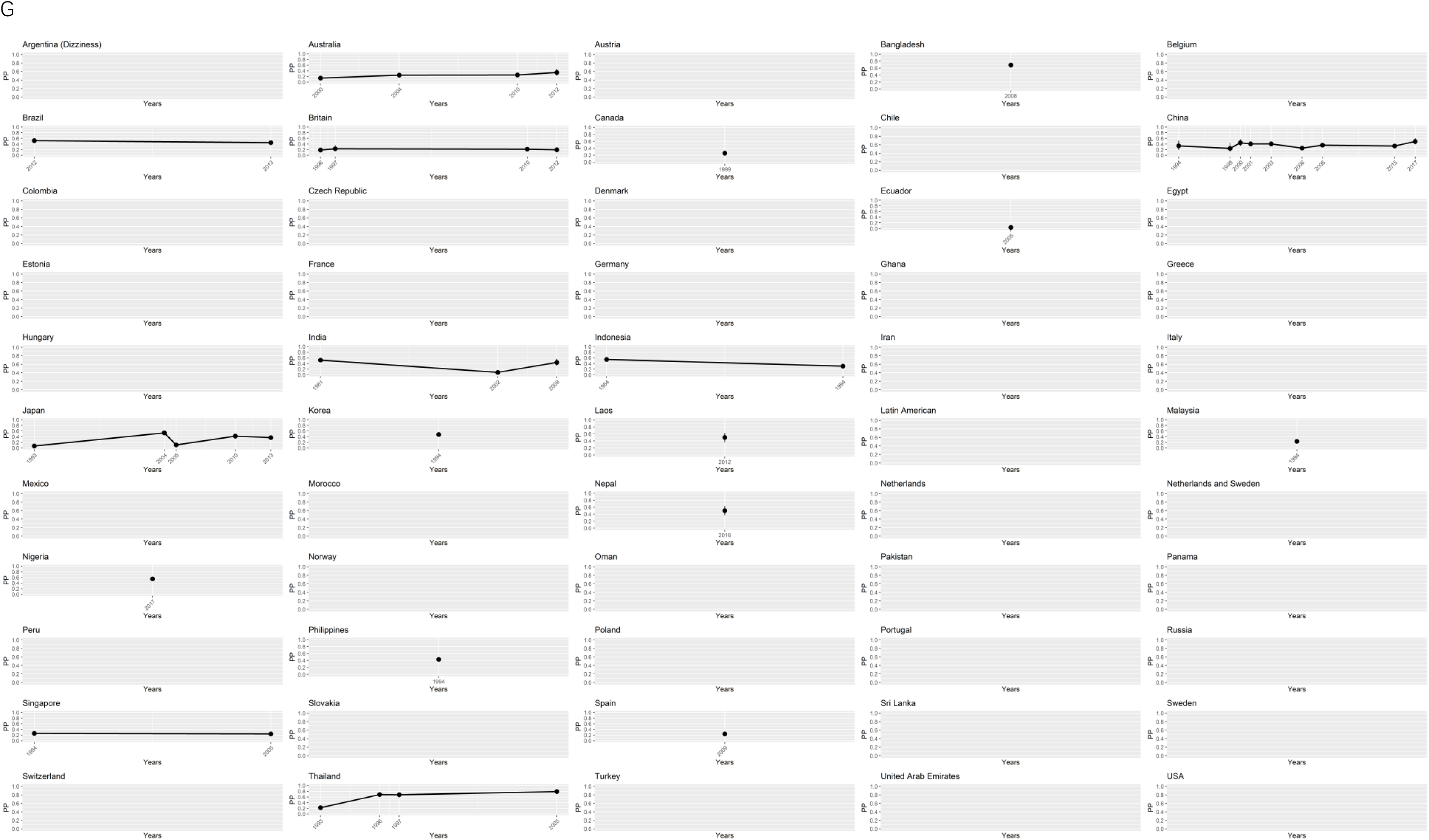
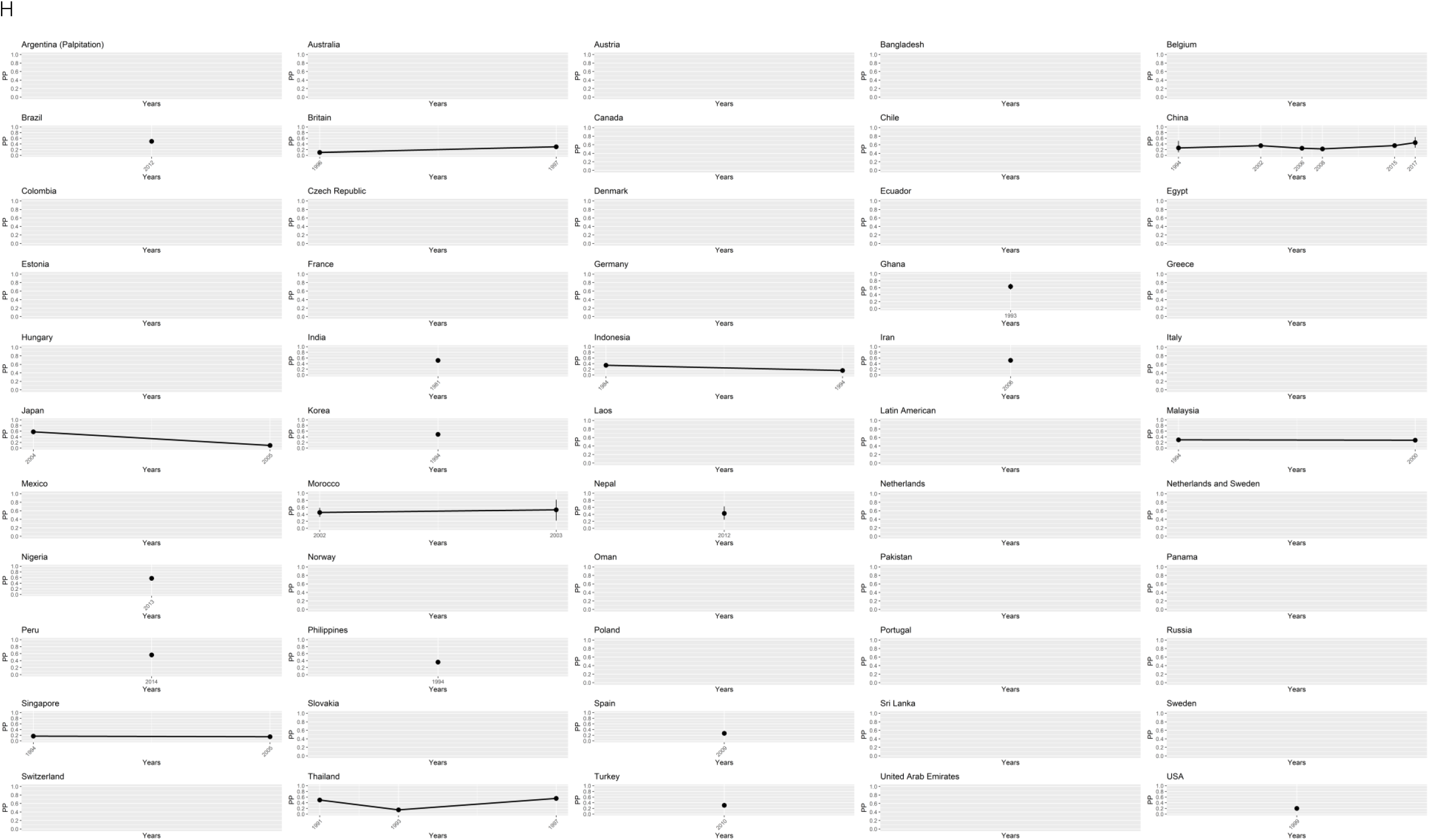
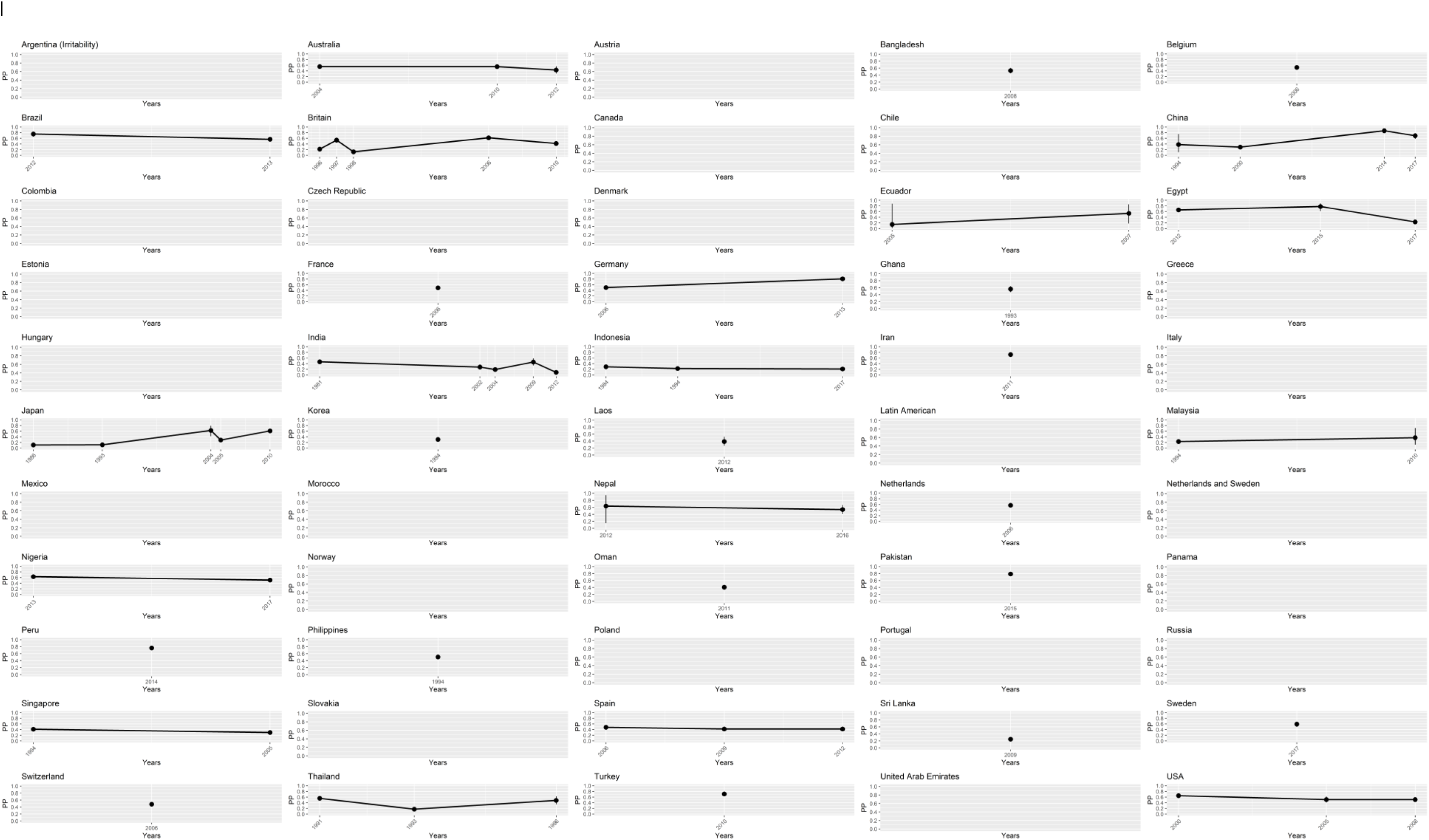
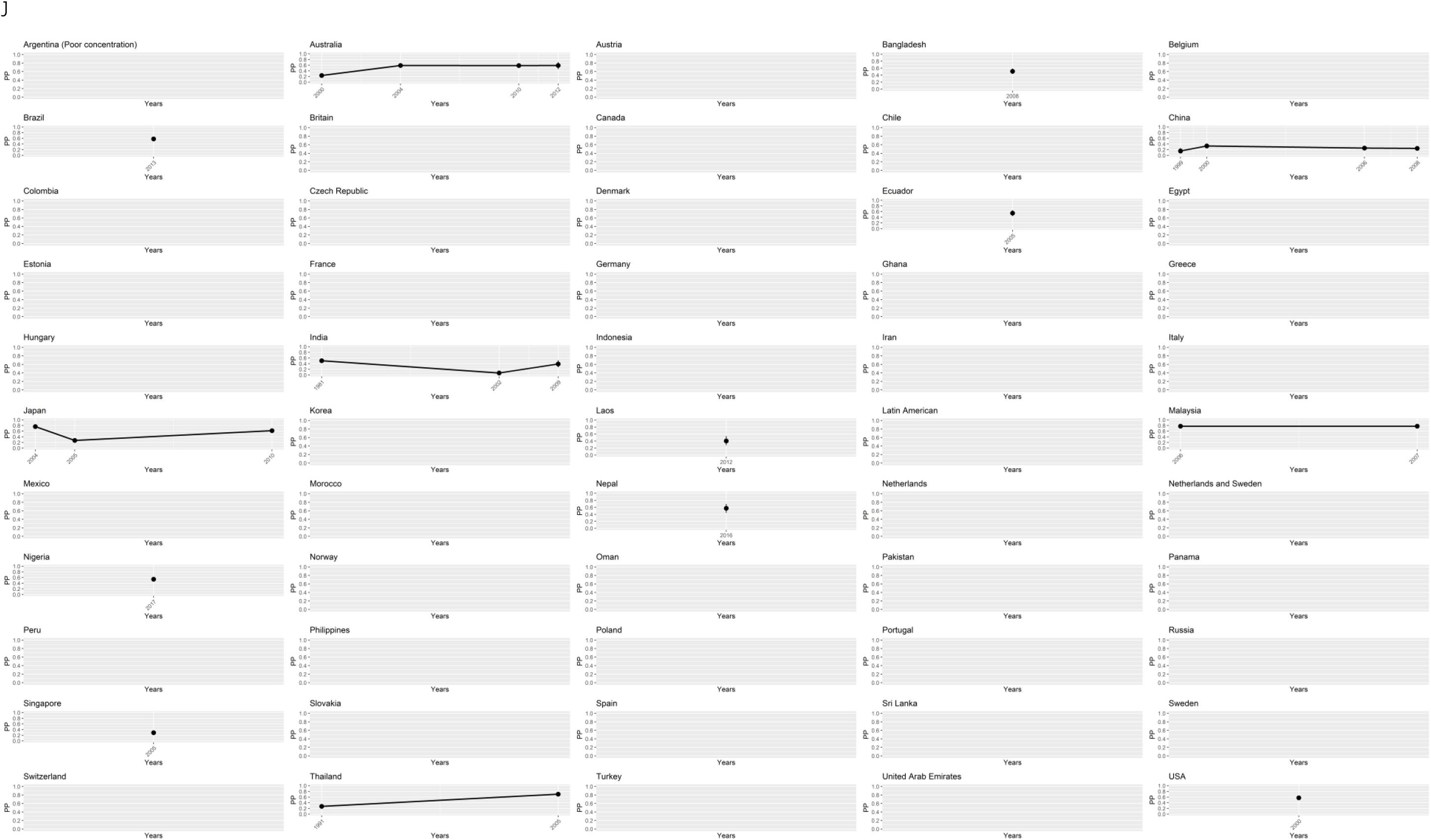
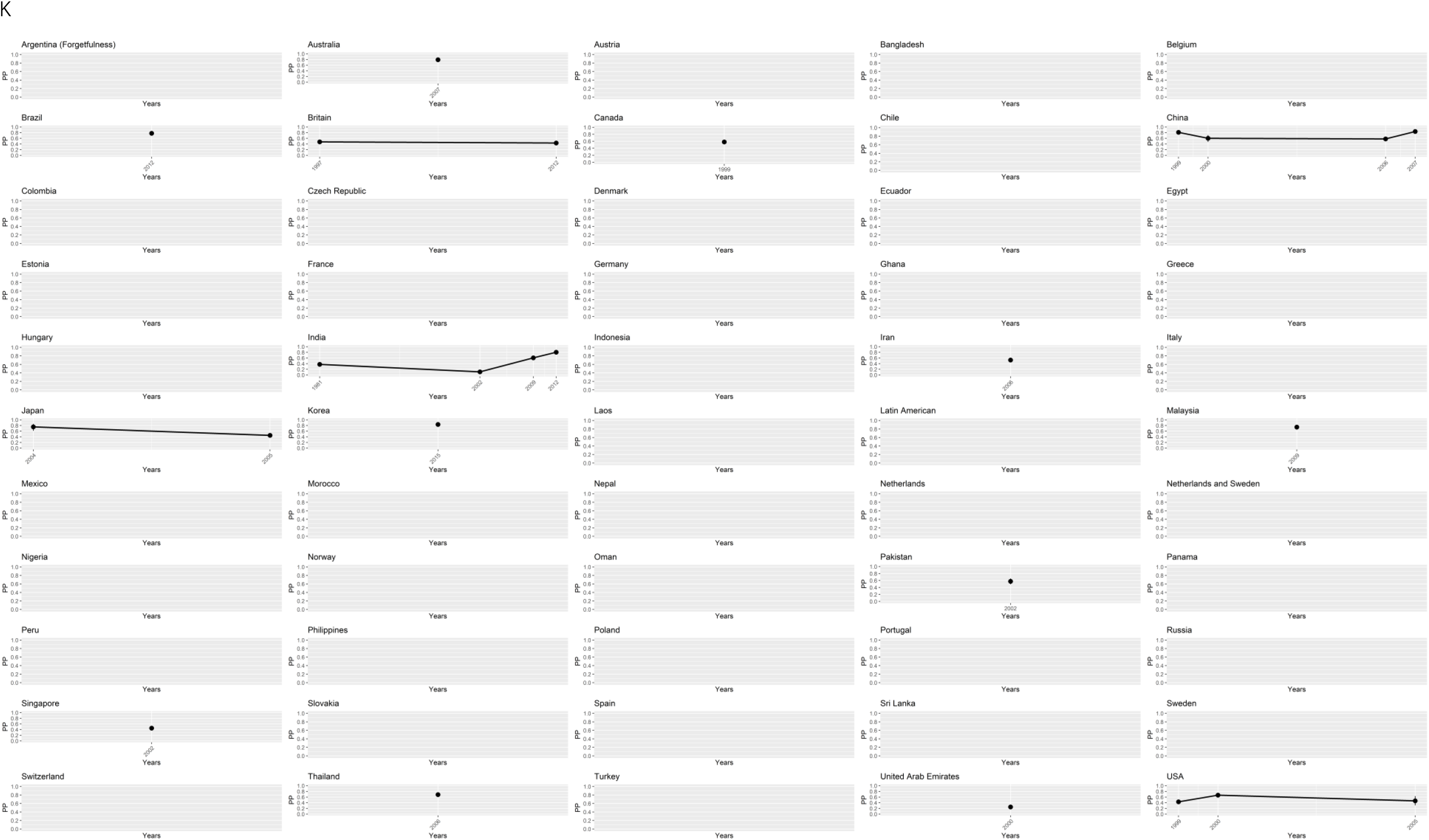
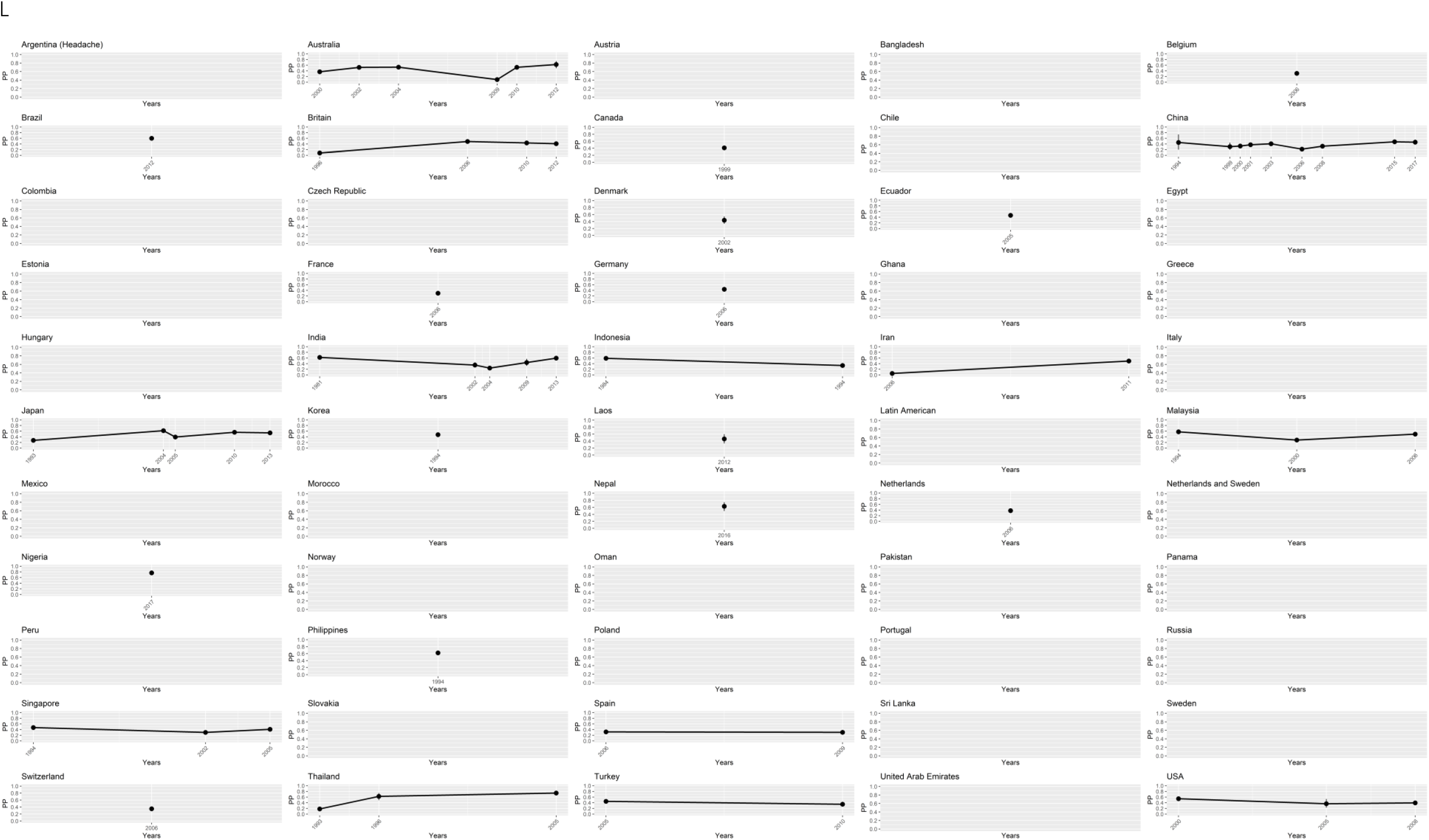
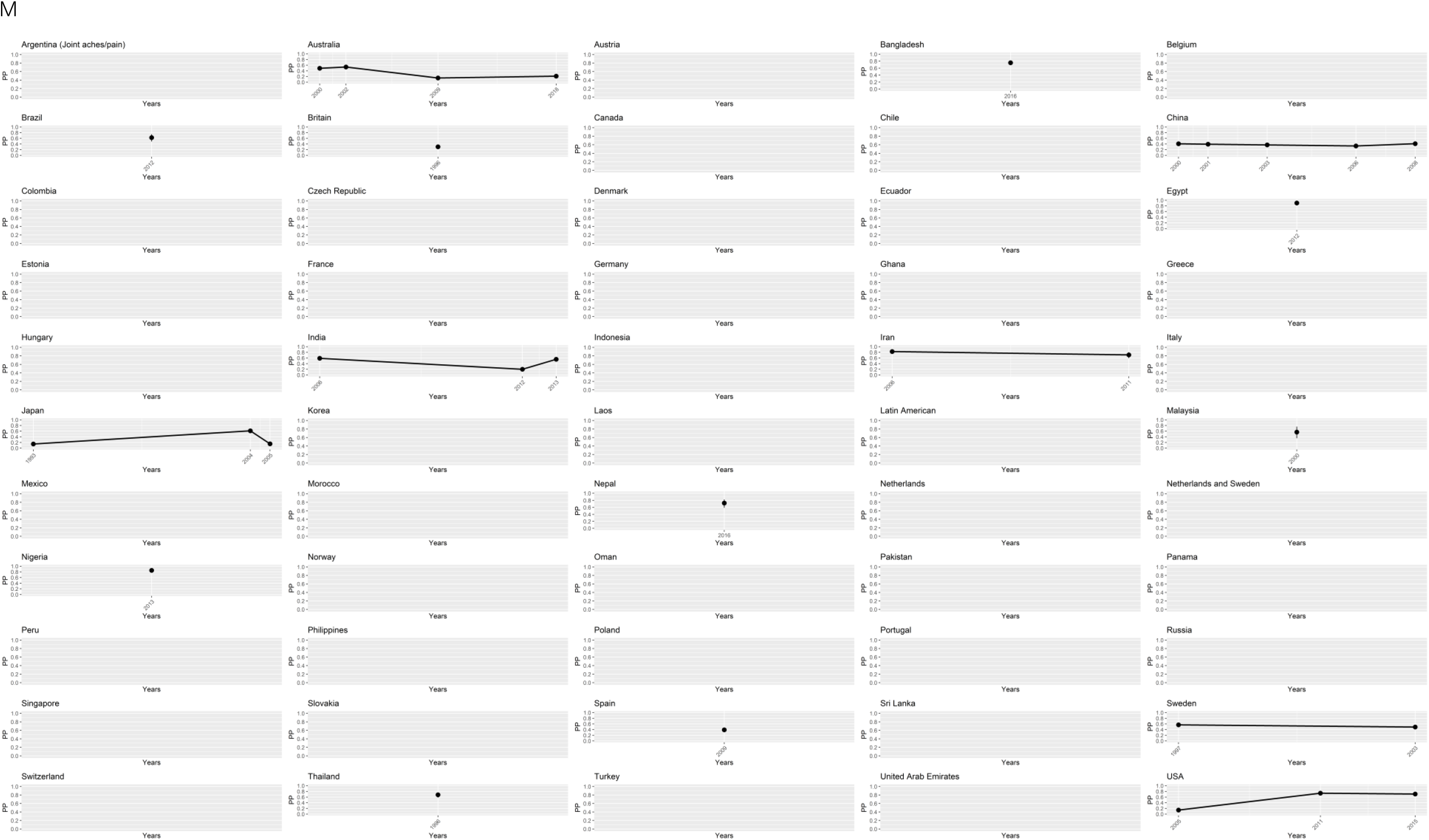
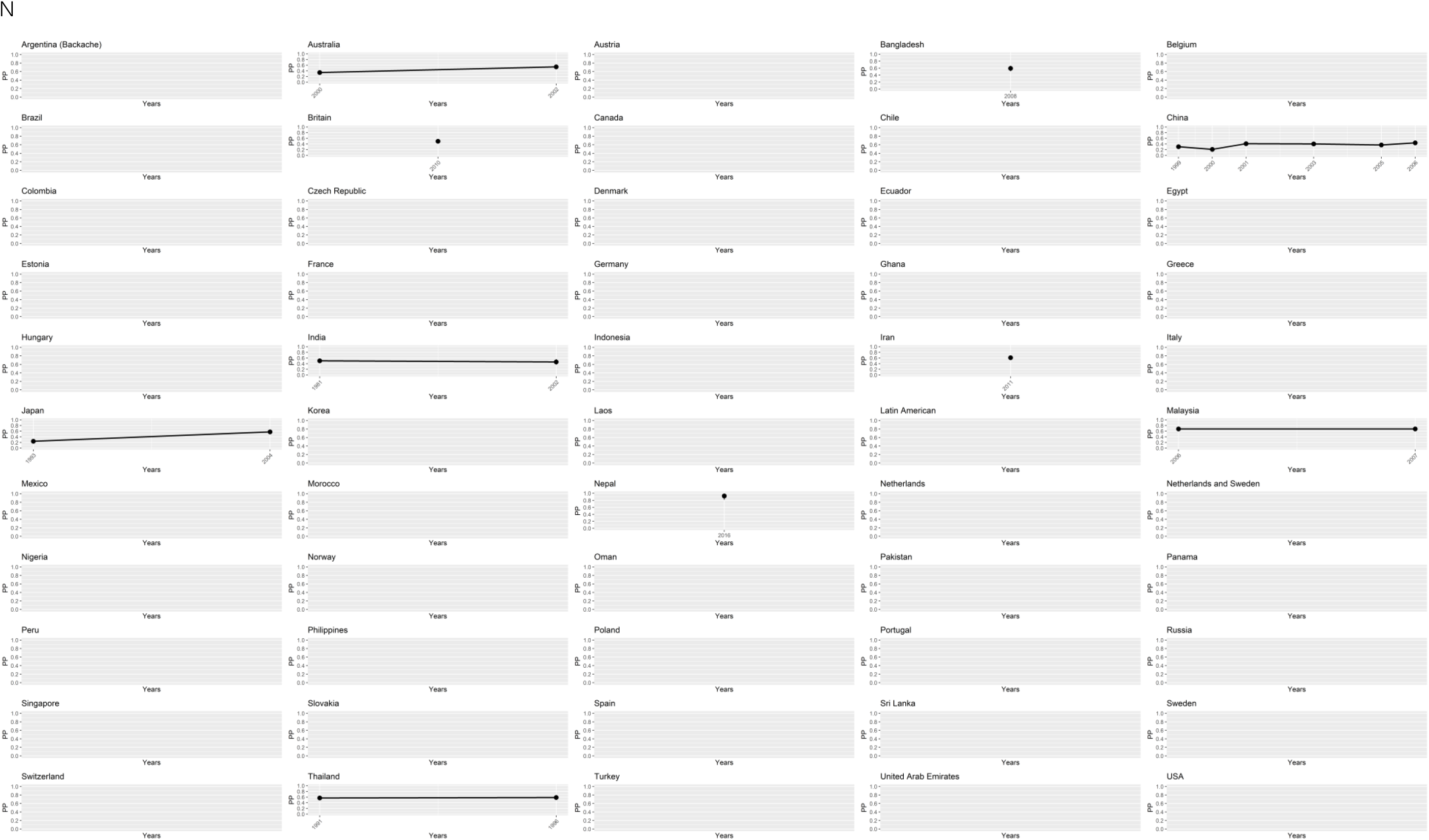
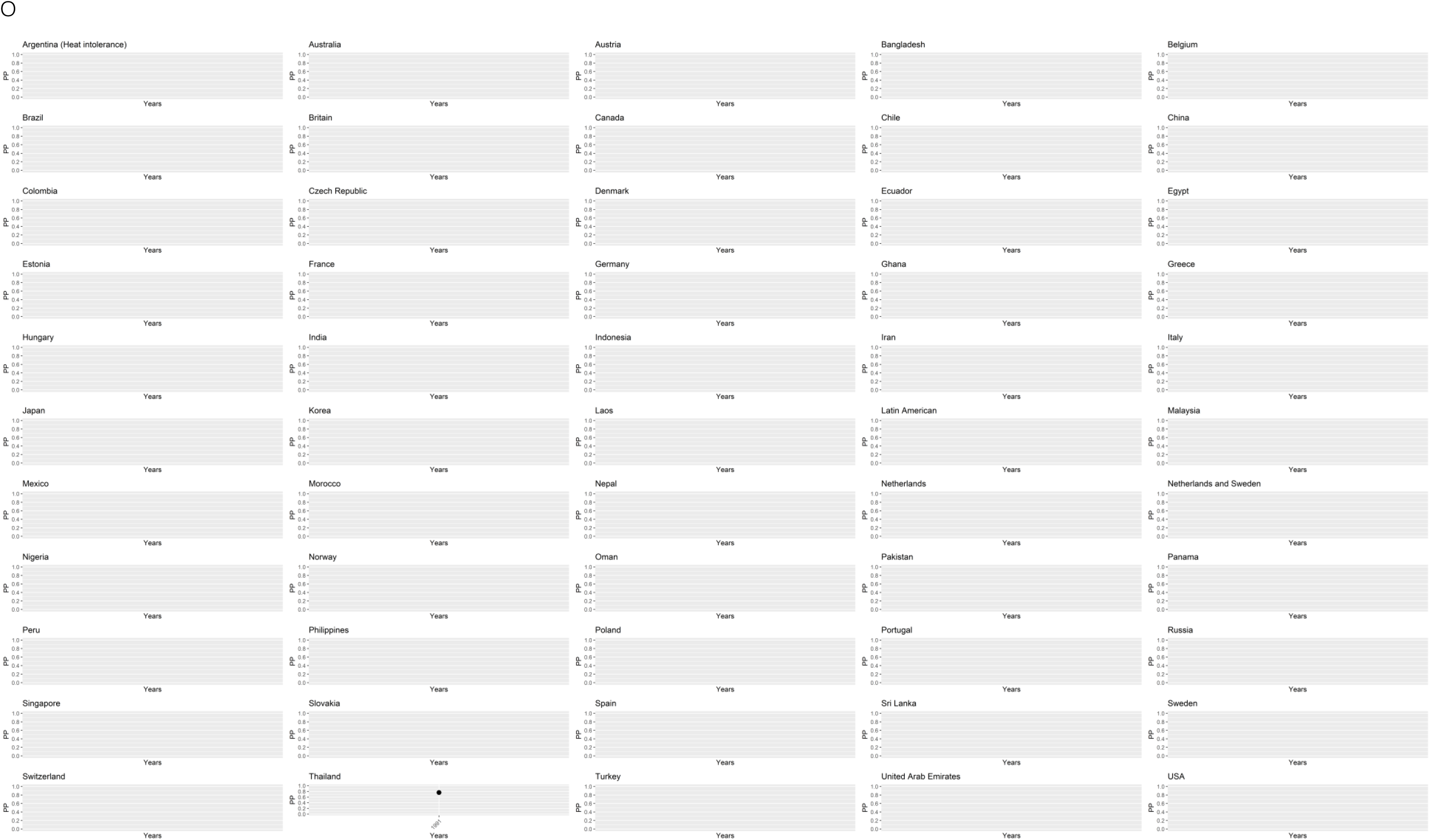
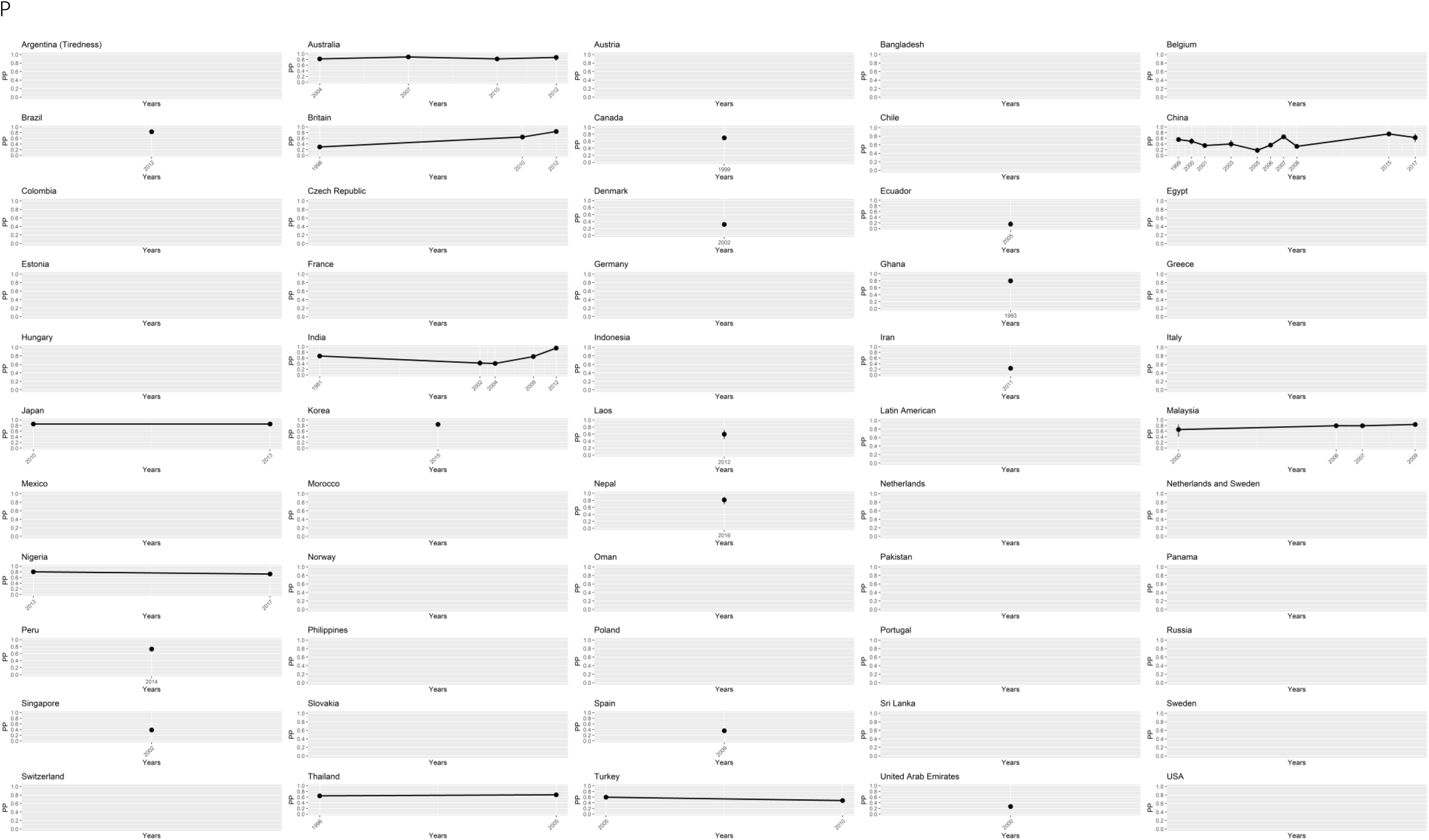
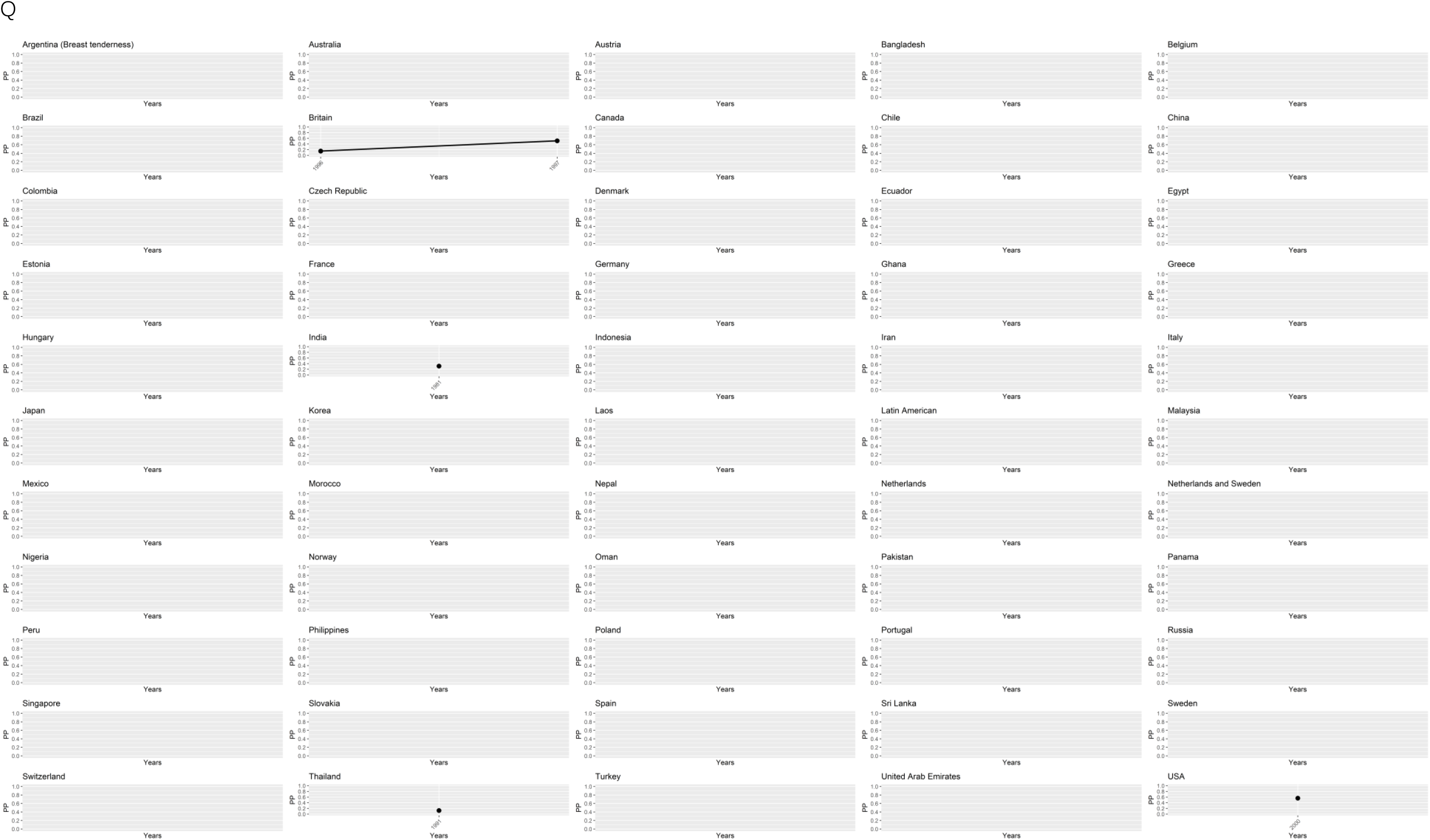
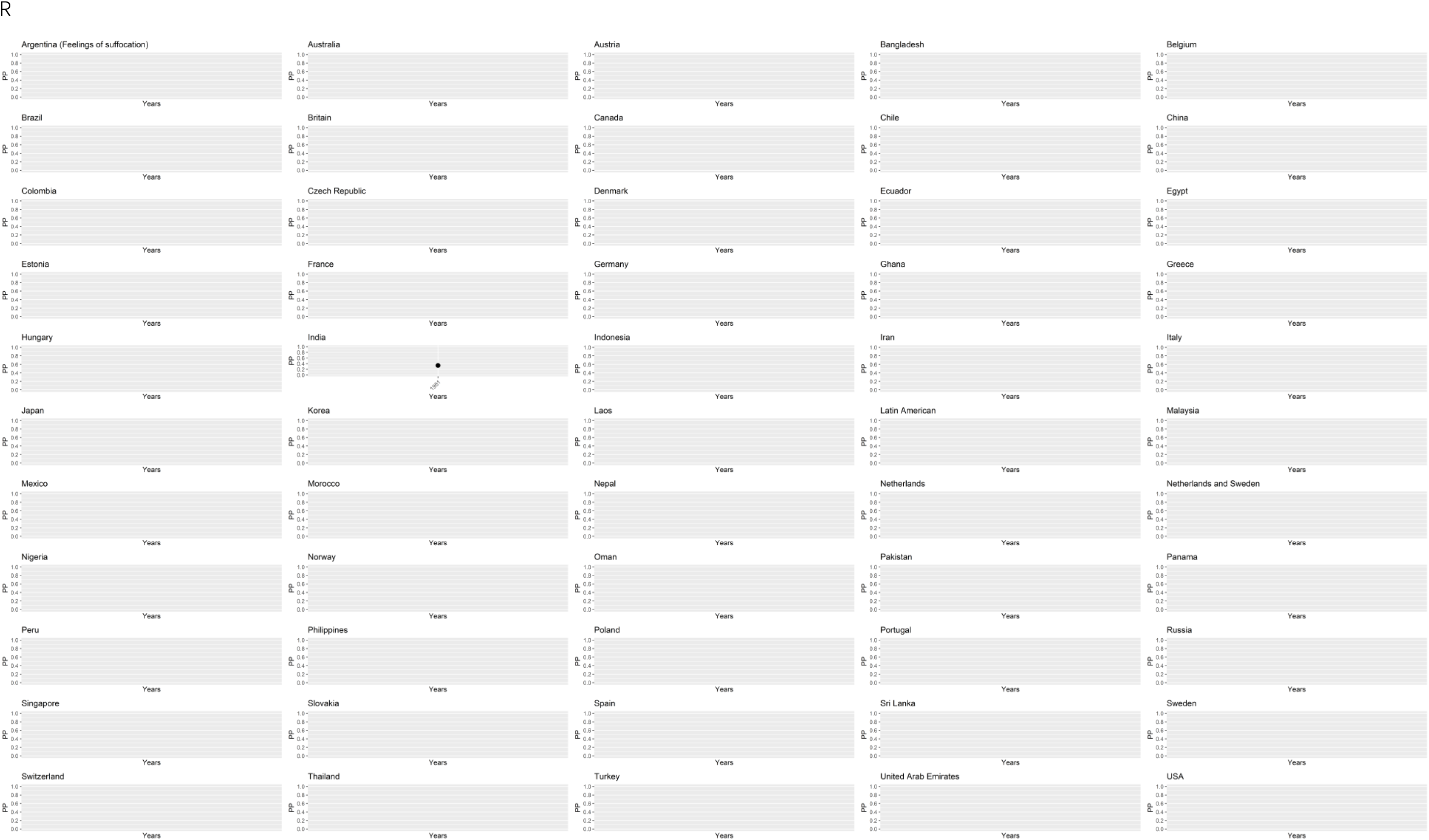
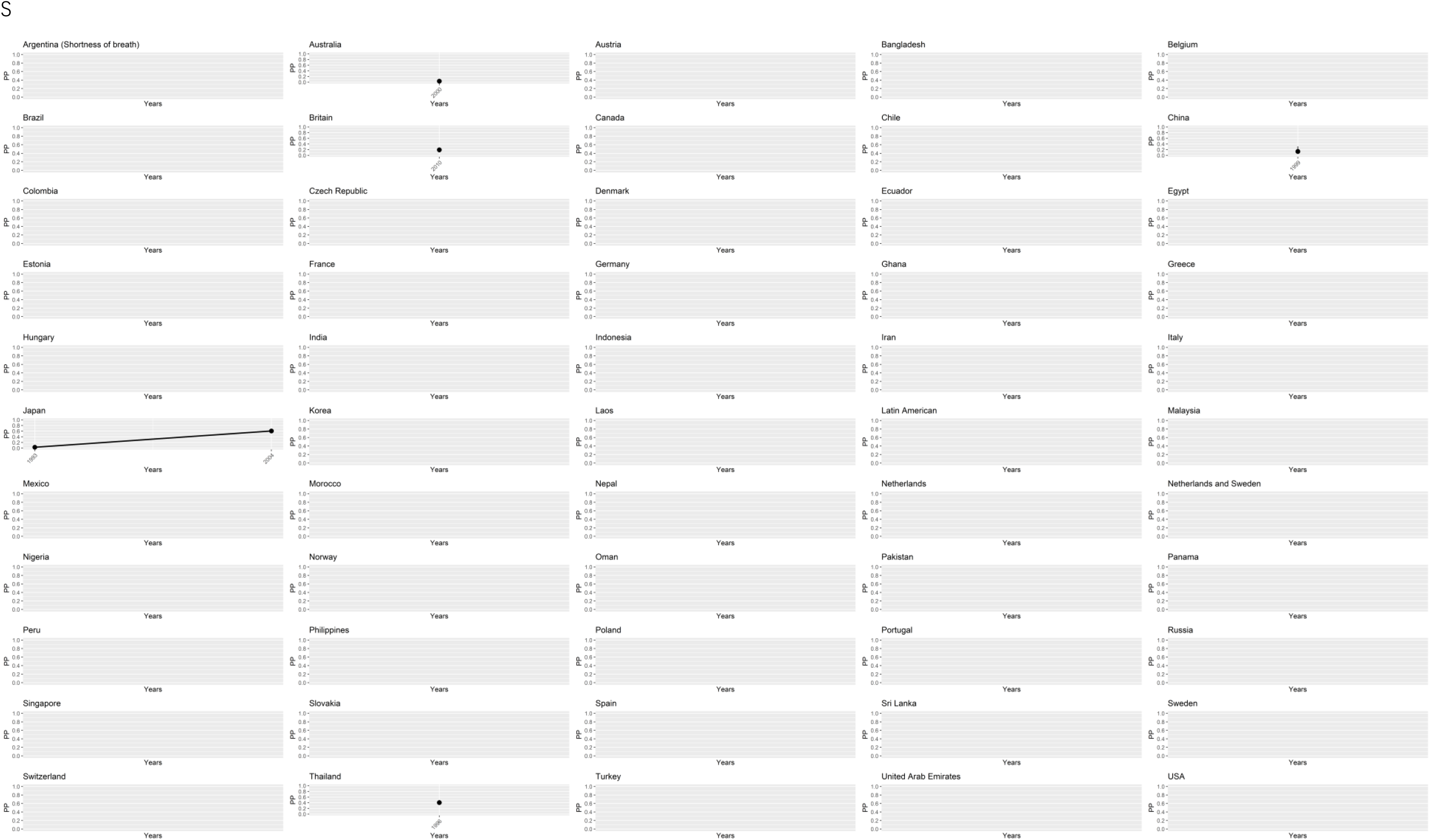
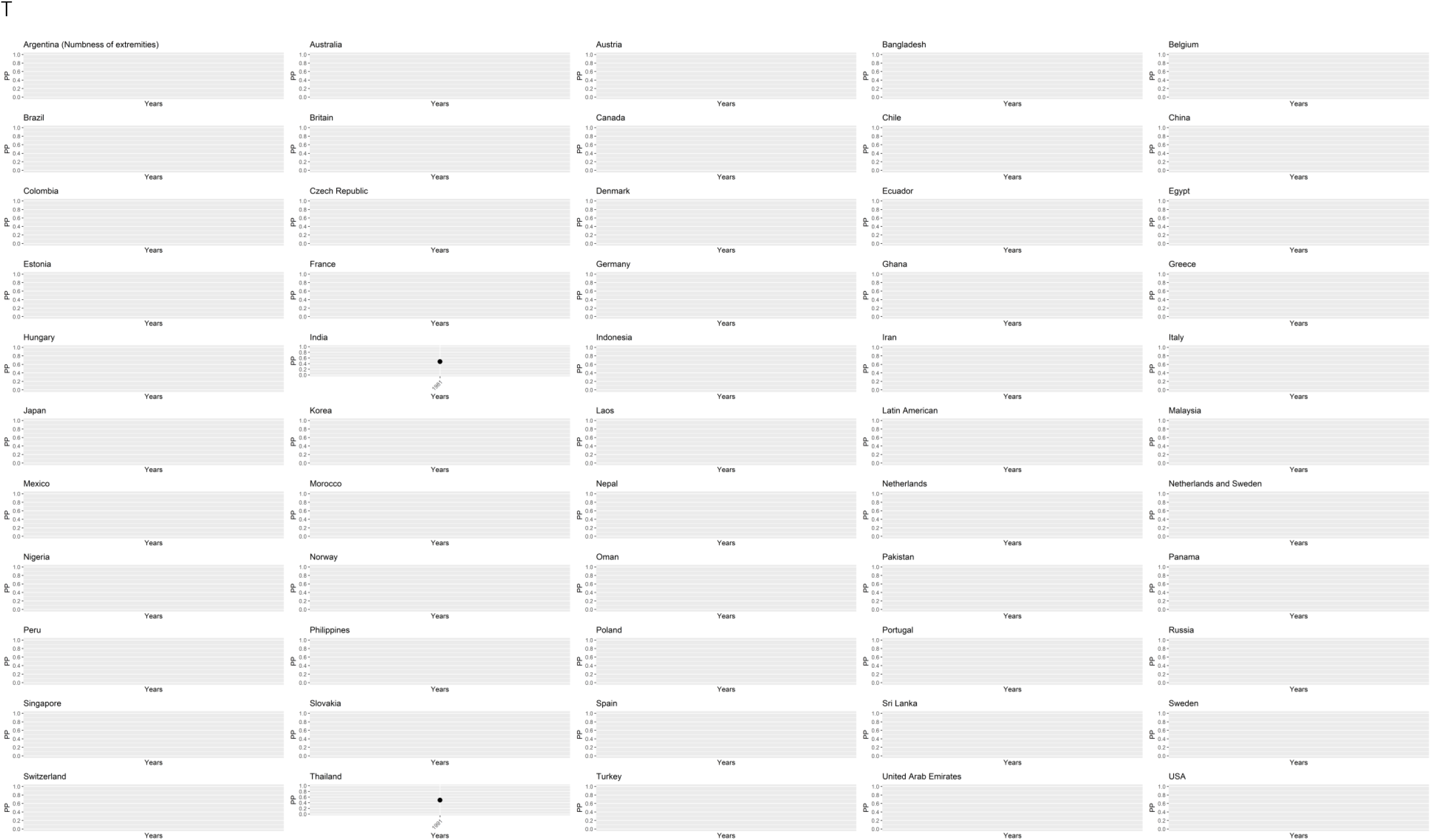
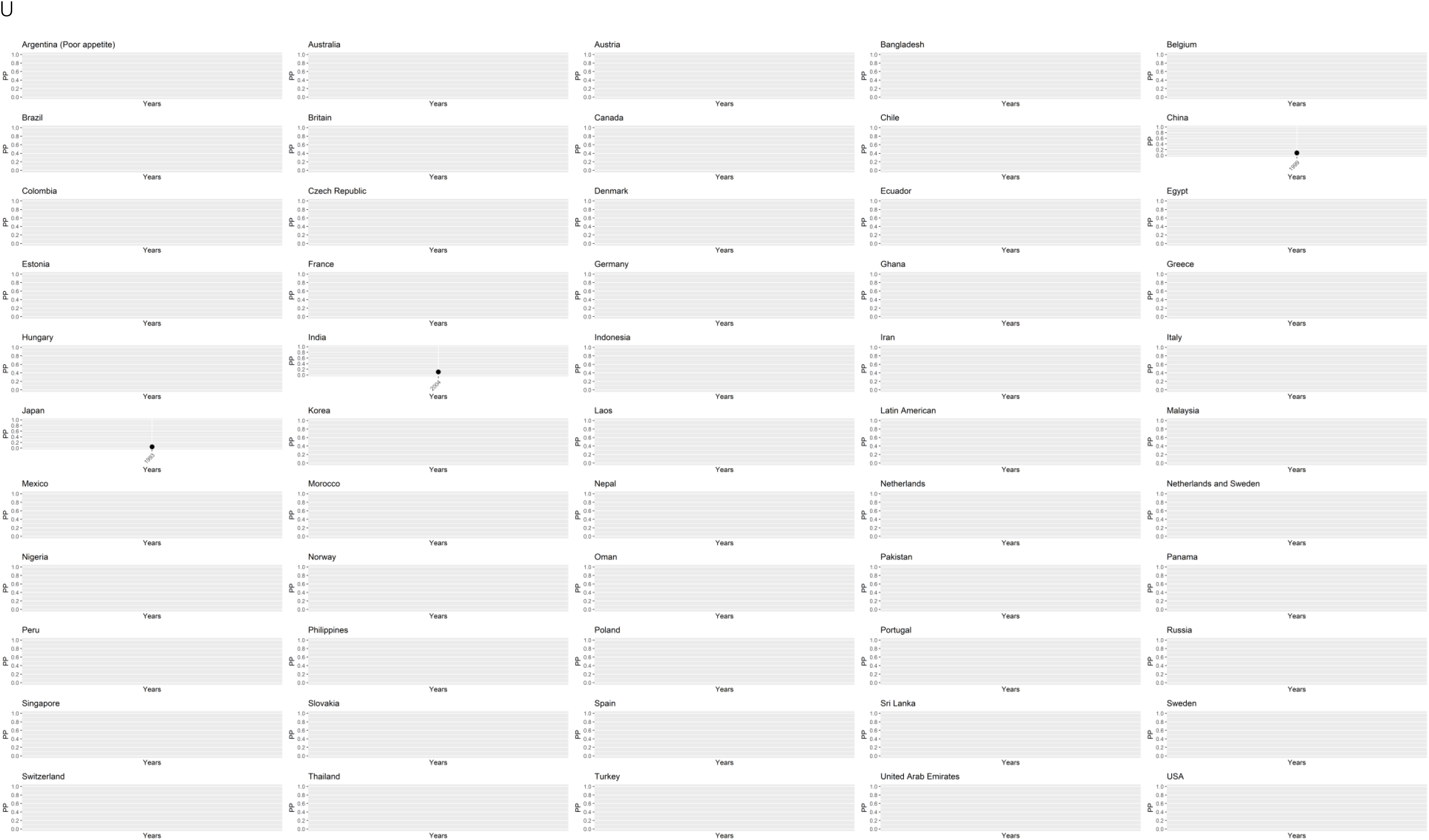
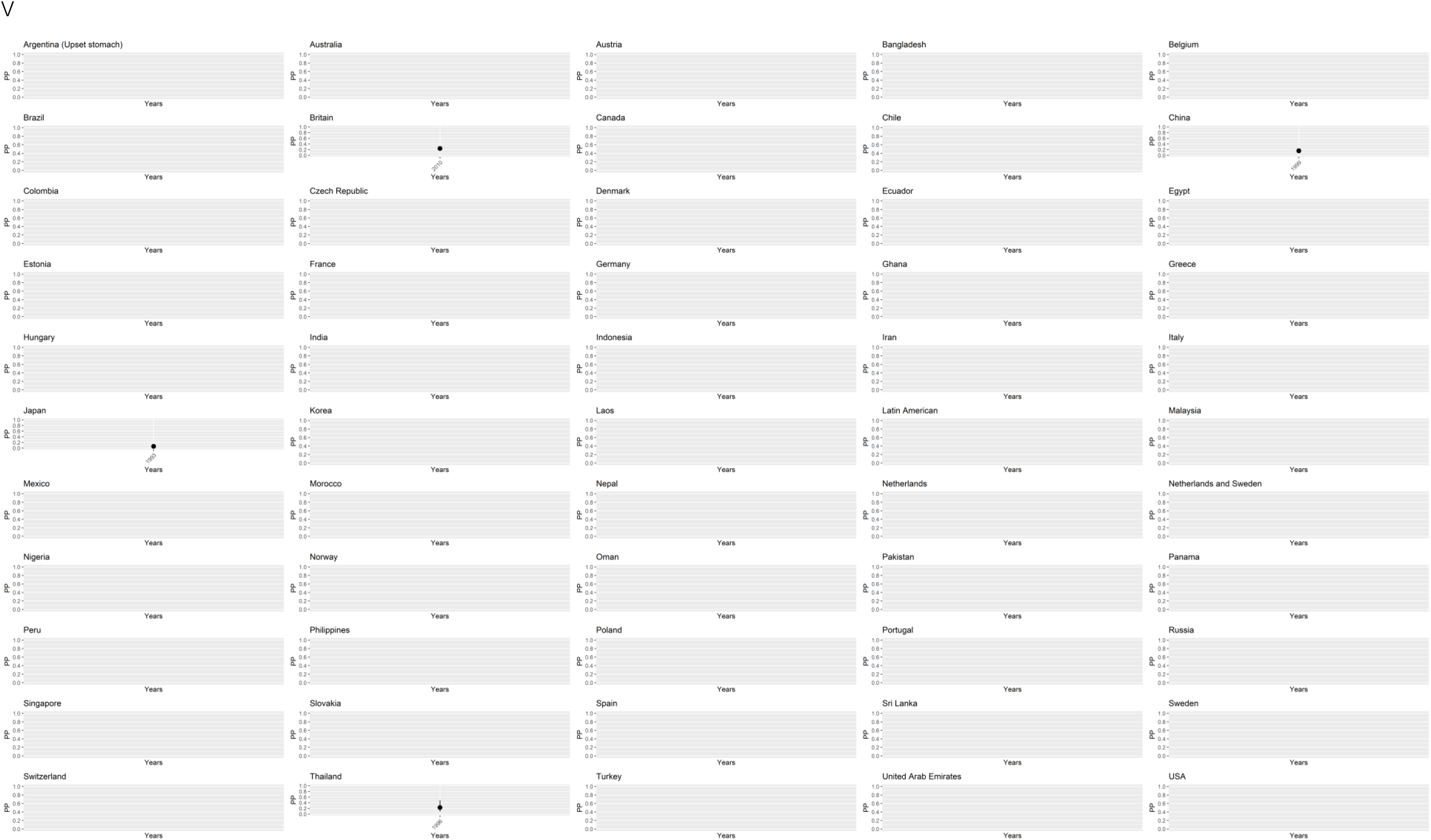
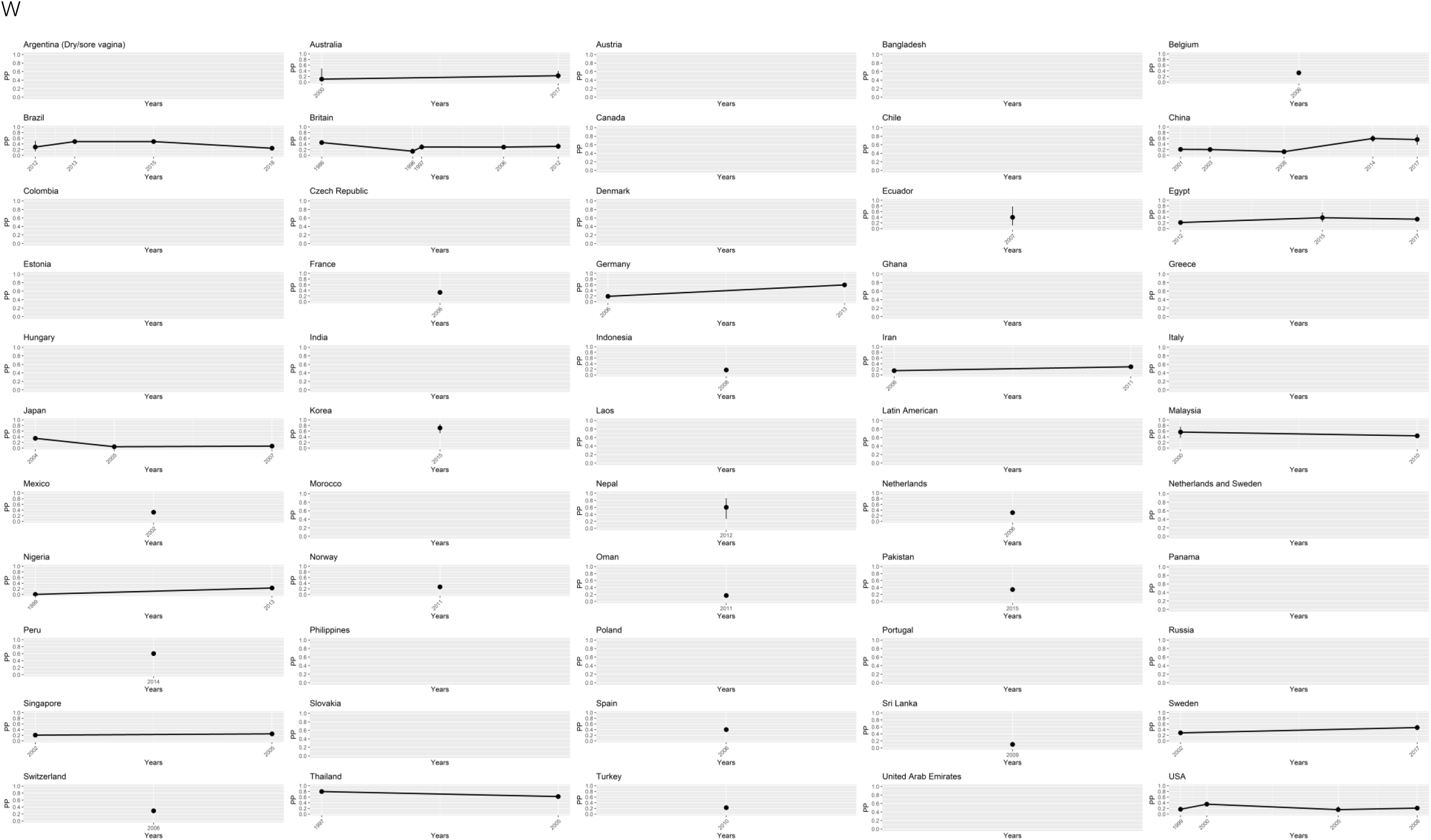
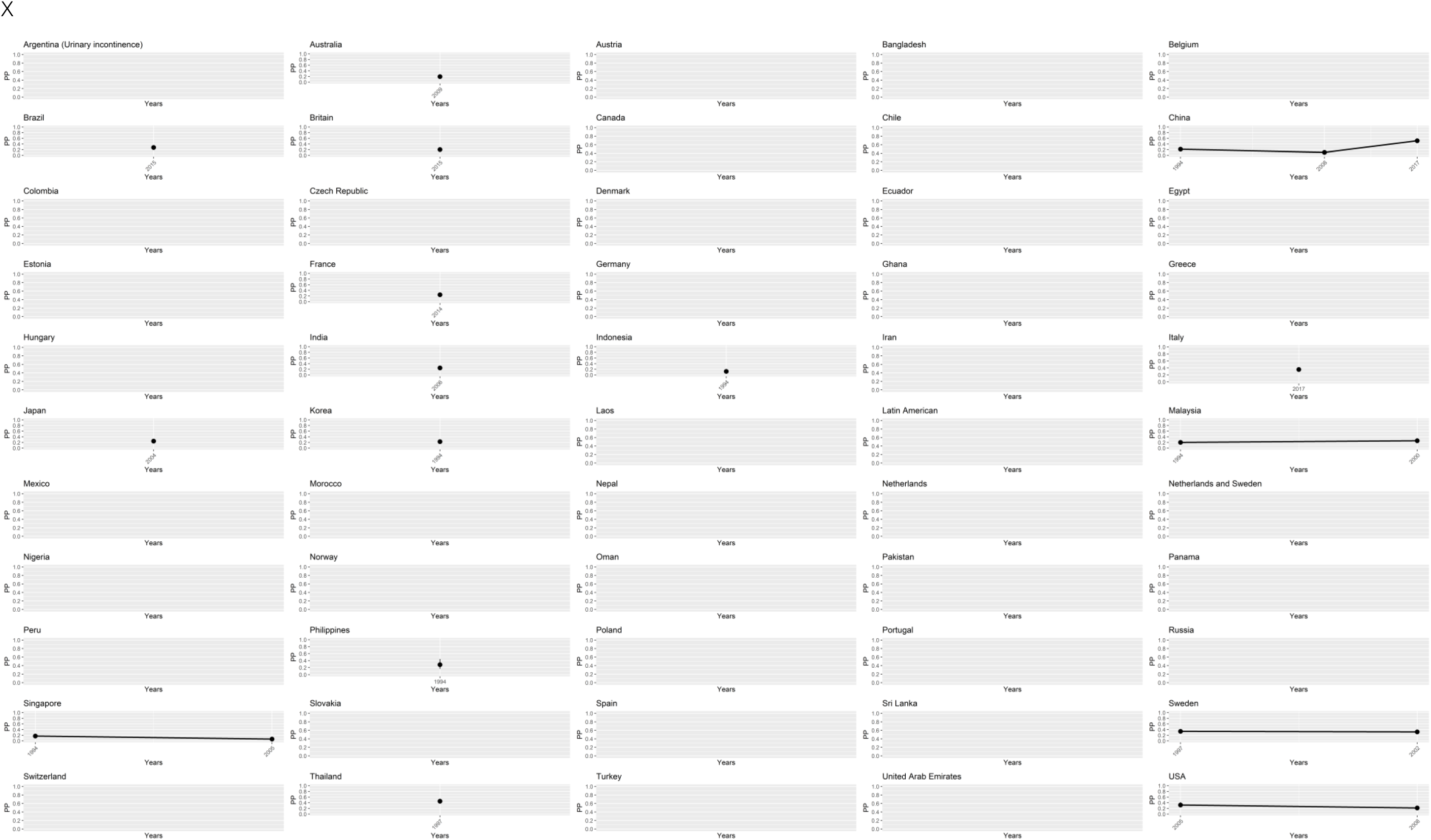
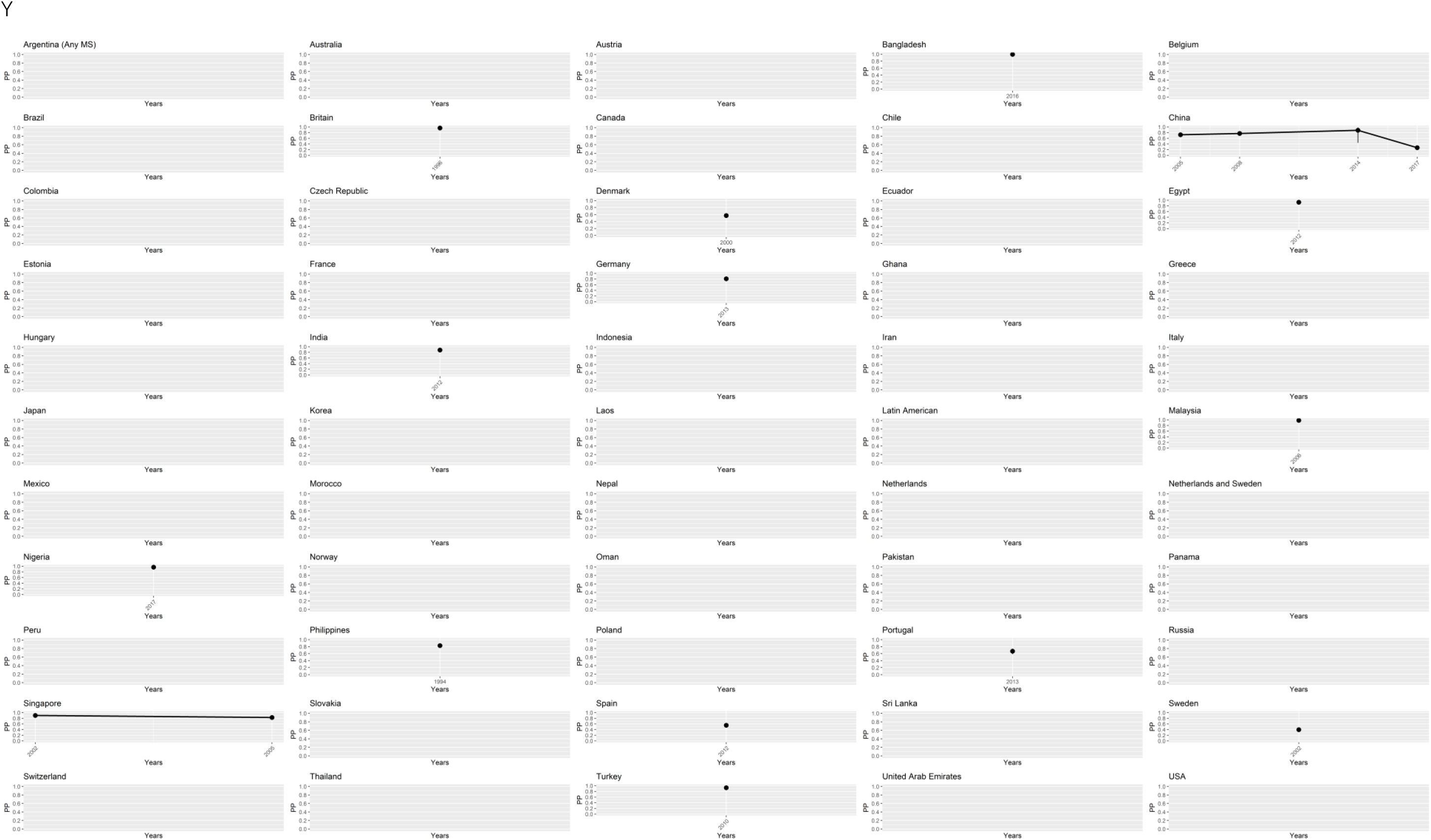

## Appendix 3. Global distribution of prevalence rates of other menopausal symptoms across different countries using world maps

**Figure 3.1.**
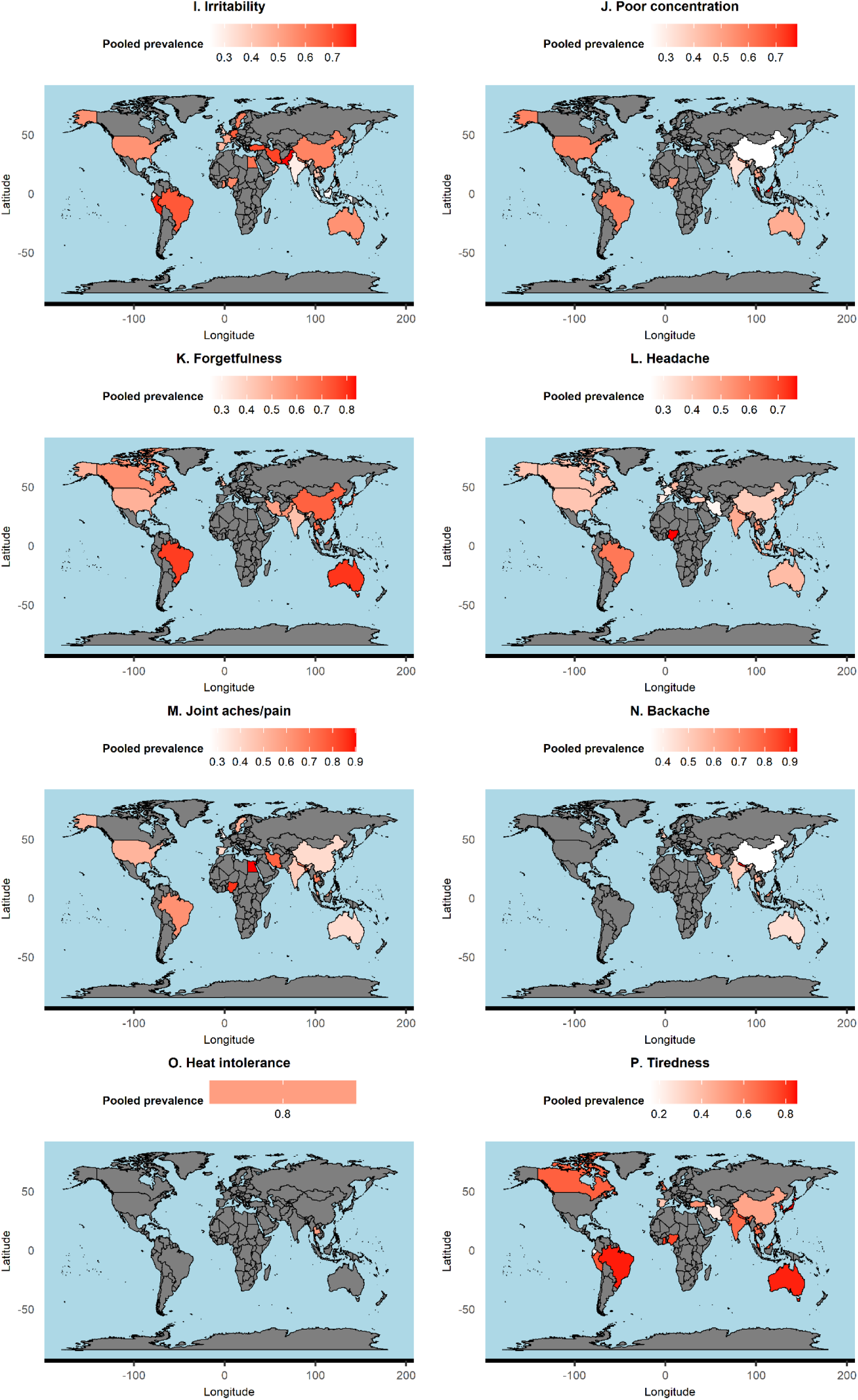
Global distribution of other menopausal symptoms by countries.

**Figure 3.2.**
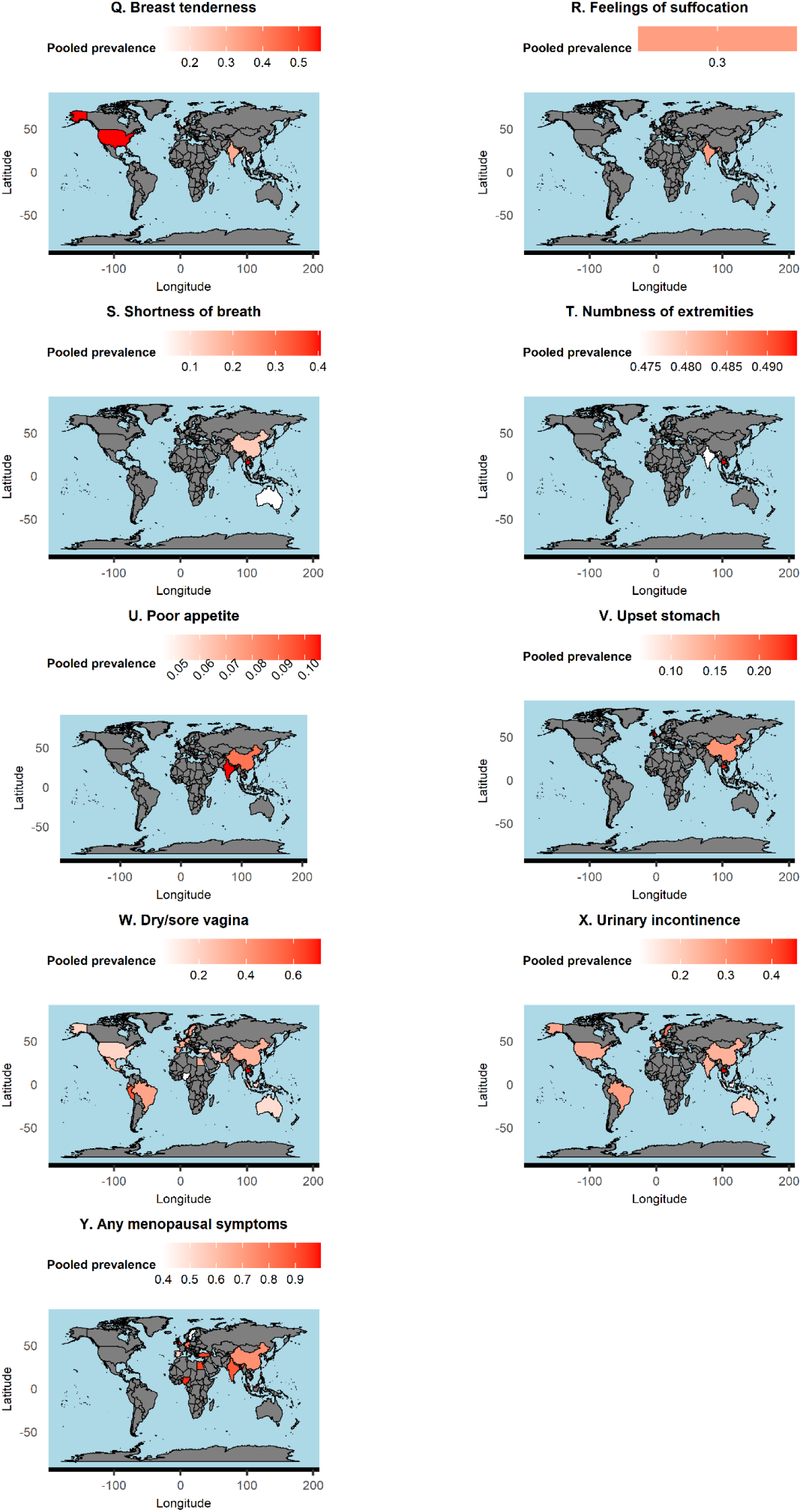
Global distribution of other menopausal symptoms by countries.

---

**Supplementary Table S1.**
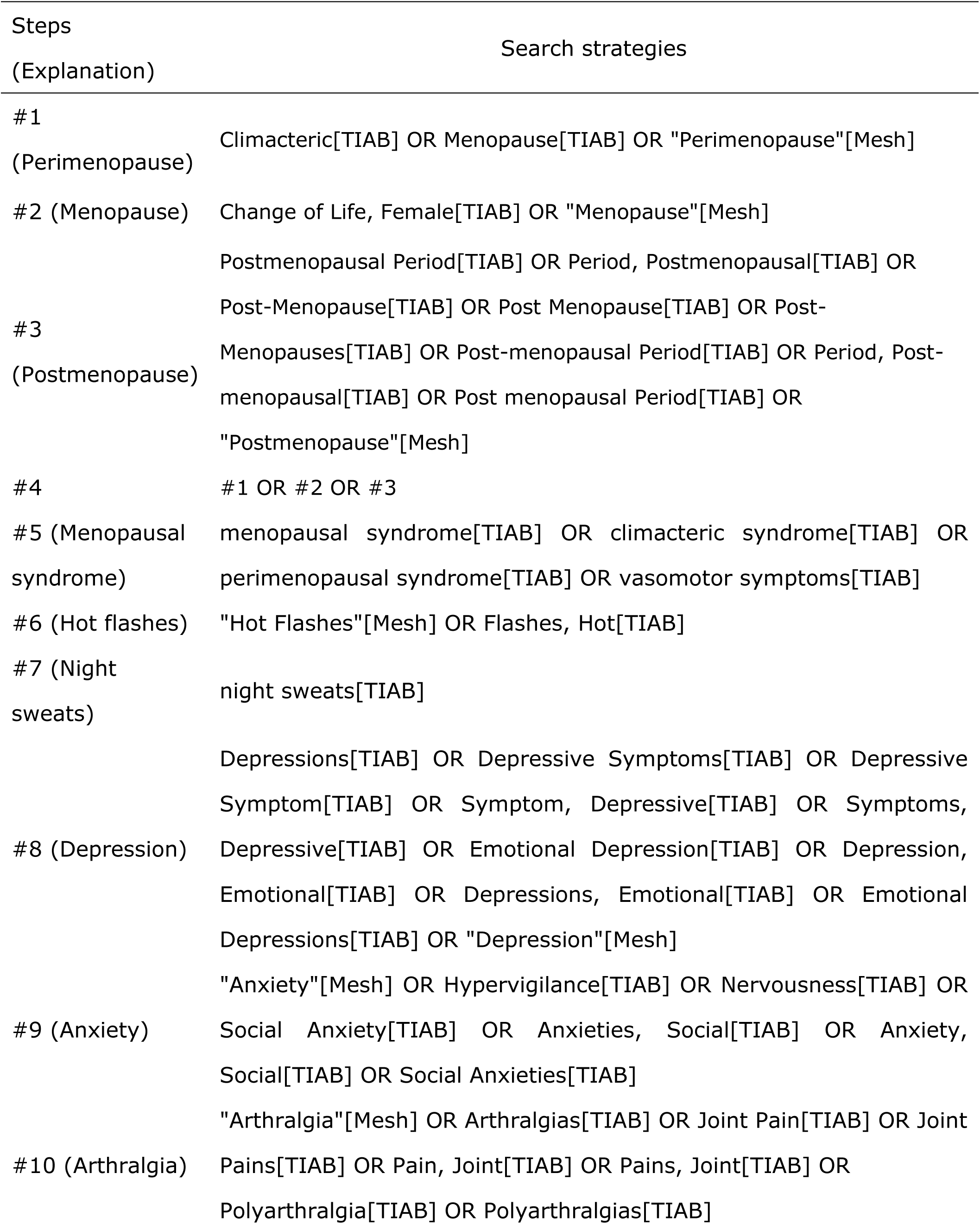
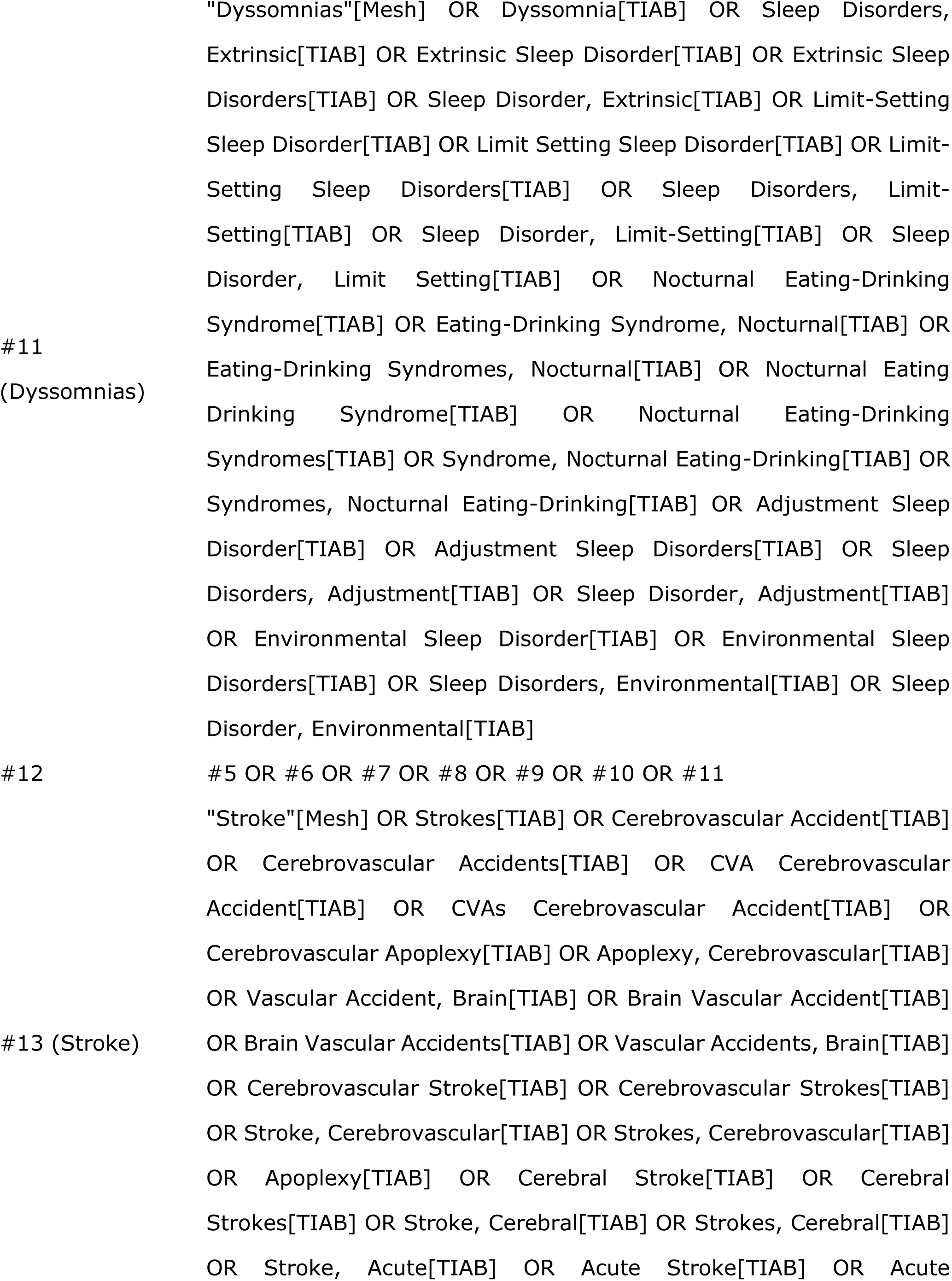
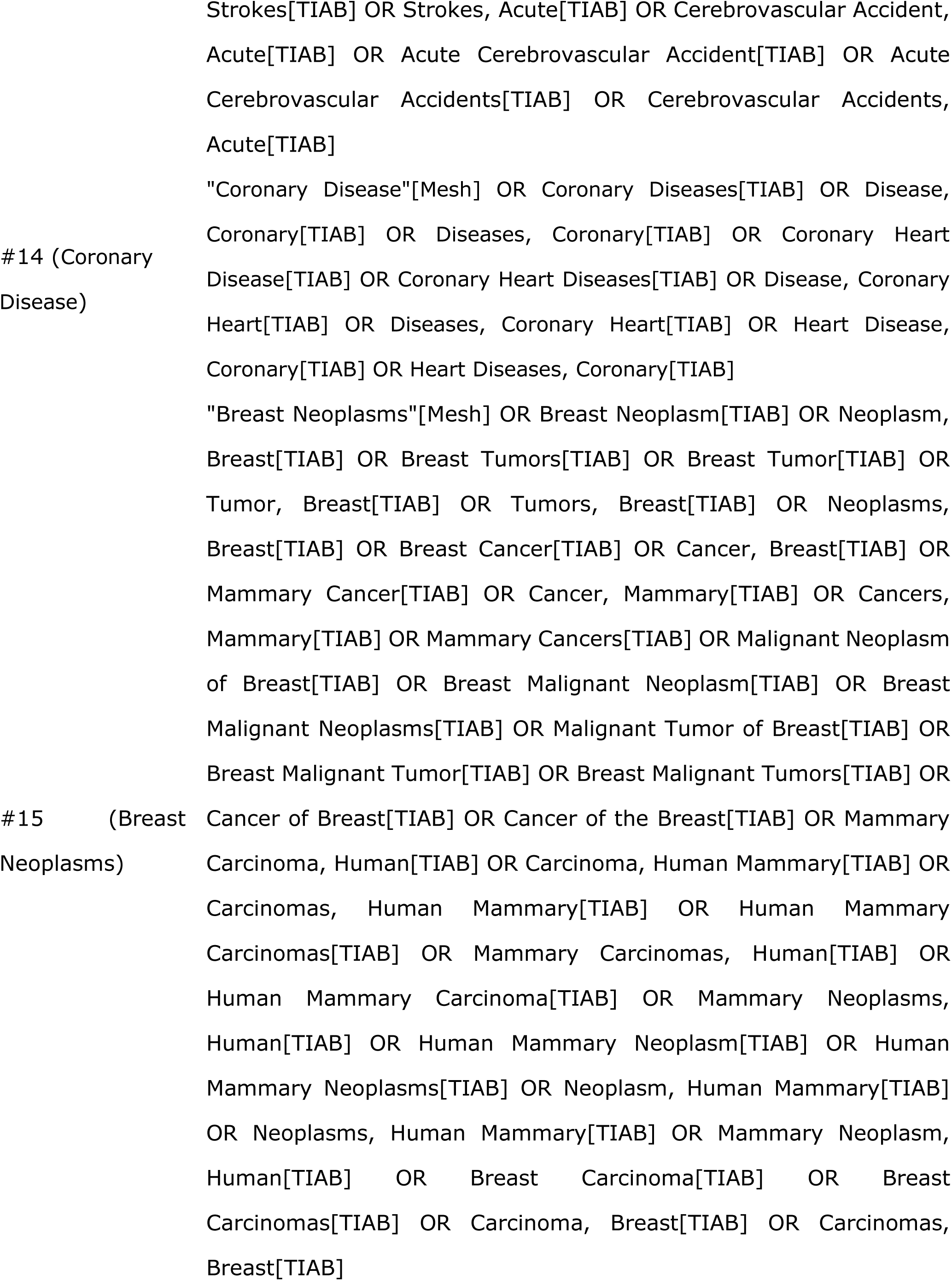
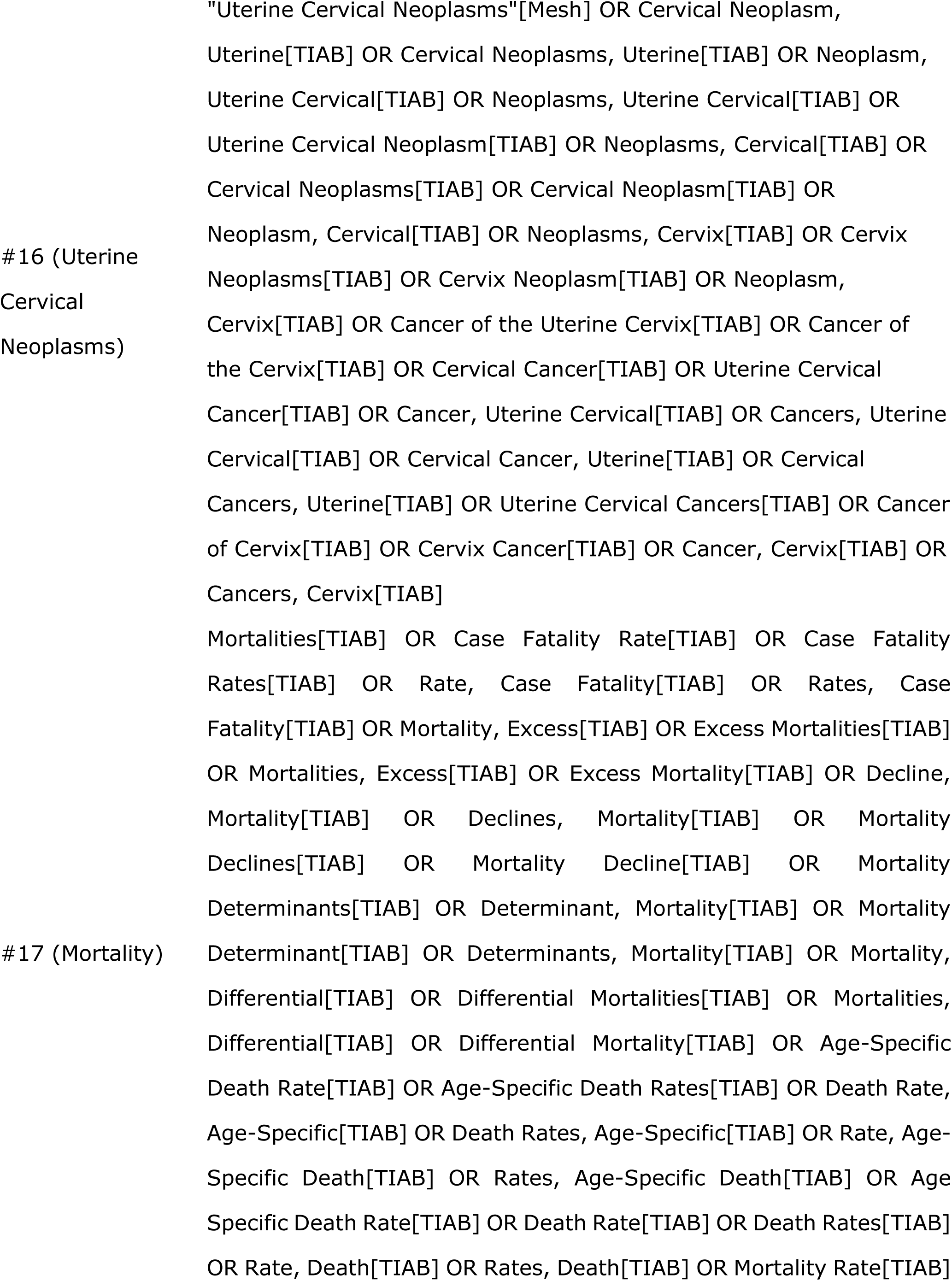
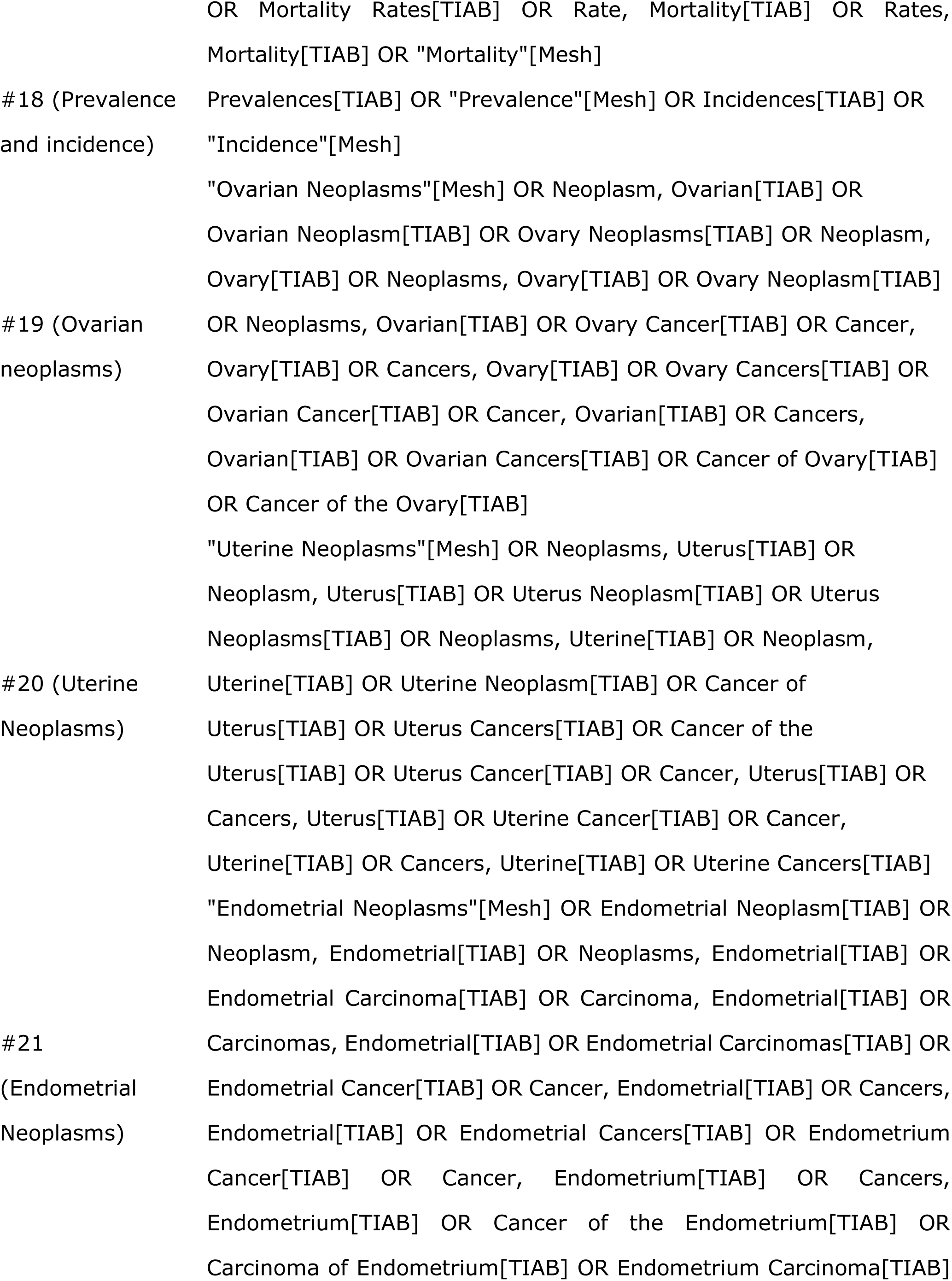
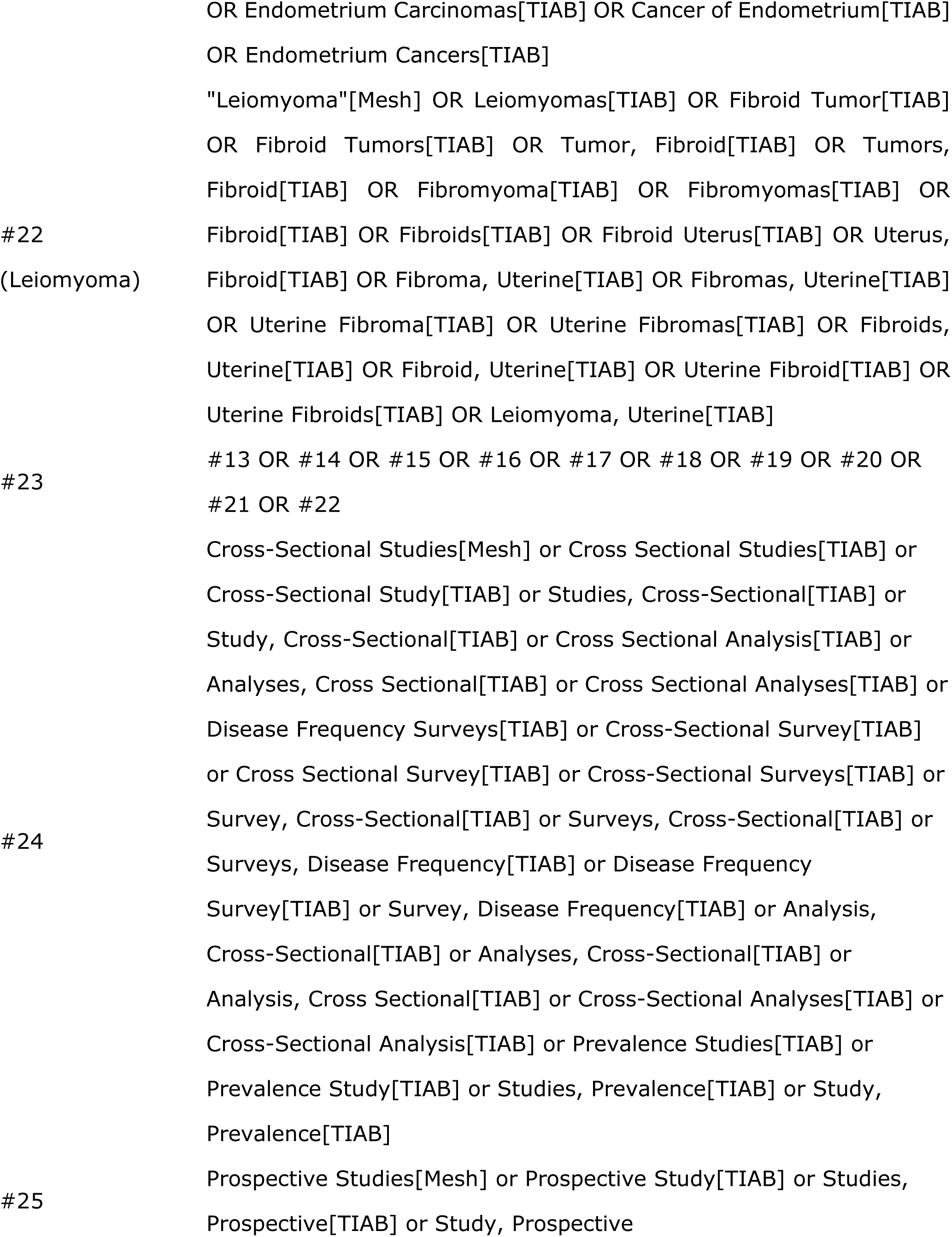
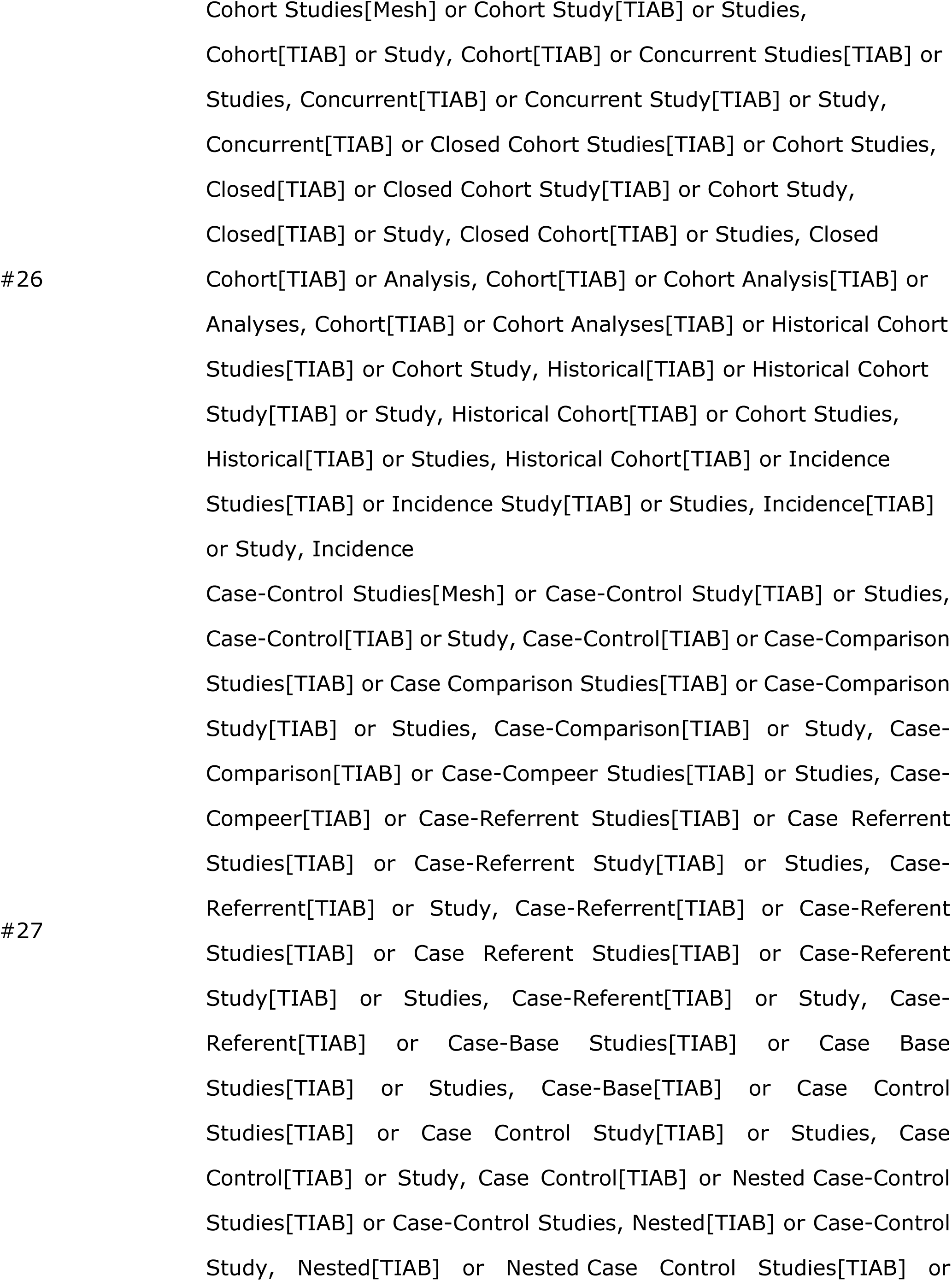
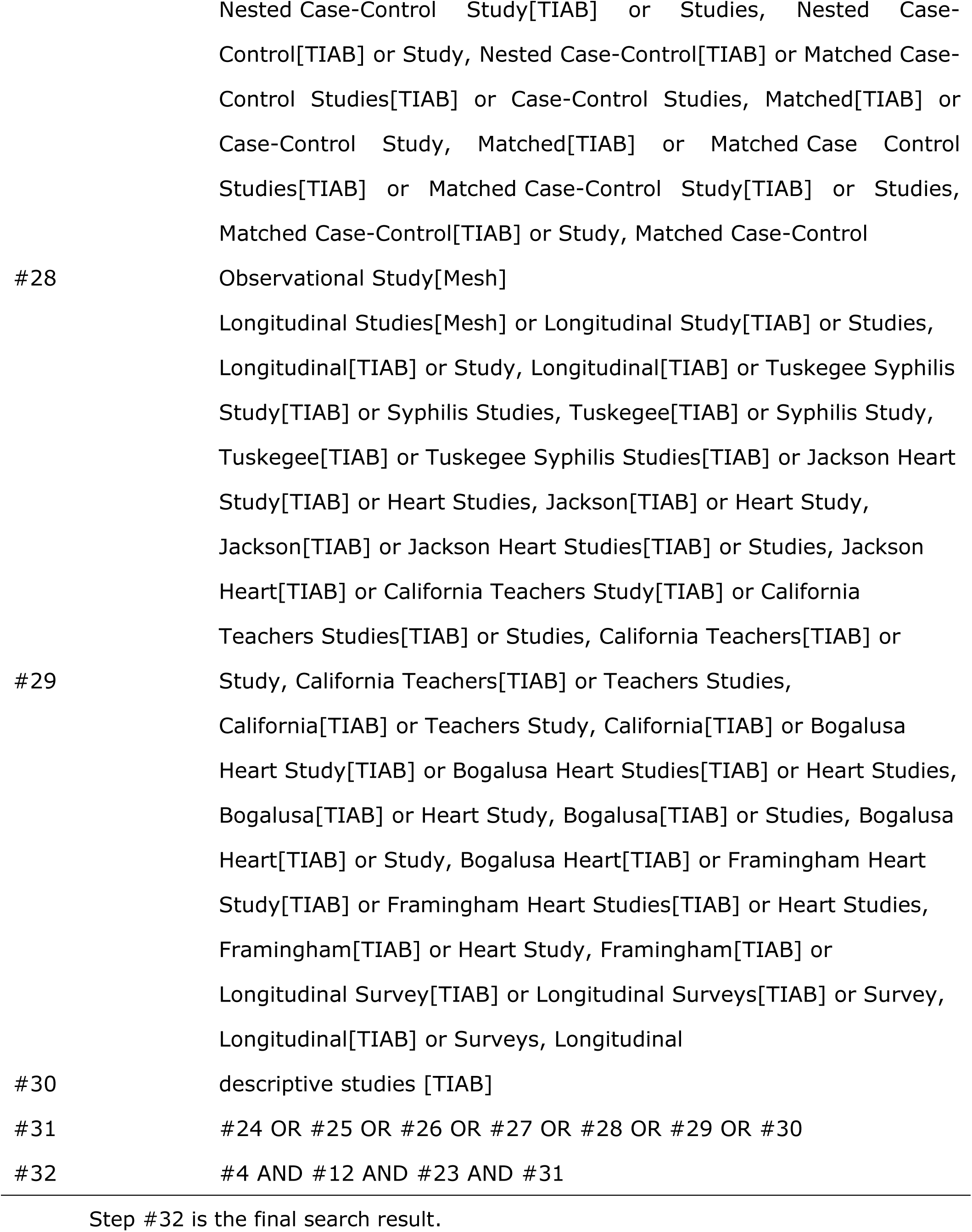
Search strategies of this meta-analysis.

